# Unravelling the genetic basis of stuttering: GWAS meta-analysis highlights link with rare speech disorders

**DOI:** 10.64898/2026.09.15.26363183

**Authors:** Victoria E. Jackson, Jenna J. Shin, Sarah Horton, Jessica O. Boyce, Else Eising, Olivia van Reyk, Richard Parker, Daisy G.Y. Thompson-Lake, Meghan Evans, Janet Beilby, Jennifer E. Below, Dorret I. Boomsma, Elinor Bridges, Elizabeth C. Corfield, Marie-Christine J. Franken, Scott D. Gordon, Alexandra Havdahl, Simone P.C. Koenraads, Shelly Jo Kraft, Michelle Luciano, Gunn-Helen Moen, Hayley S. Mountford, Agnieszka Musial, Craig E. Pennell, Hannah G. Polikowsky, René Pool, Valerie A. Rebattu, Kaili Rimfeld, Alyssa C. Scartozzi, Beate St Pourcain, Ingrid A. Szilagyi, Stian B. Valand, Kathryn Z. Viljoen, Carol A. Wang, Andrew J.O. Whitehouse, Yvonne E. Wren, Elsje van Bergen, Nathan A. Gillespie, Adam P. Vogel, Ingrid E. Scheffer, Michael S. Hildebrand, Lynette G. Sadleir, Frédérique Liégeois, Sheena Reilly, Simon E. Fisher, Nicholas G. Martin, Angela T. Morgan, Melanie Bahlo

## Abstract

Developmental stuttering affects up to 11% of children, with around one-fifth developing a persistent stutter. Twin and family studies indicate a strong genetic contribution and comorbidity with other heritable traits, but the common genetic architecture of clinically ascertained stuttering, persistence and recovery remains poorly characterised. We performed a genome-wide association study (GWAS) meta-analysis of 6,096 cases and 81,629 controls of European ancestries from 18 cohorts, including 2,953 cases from stuttering-specific collections with clinically confirmed or validated self-reported phenotypes. We additionally analysed persistence, severity and frequency, and integrated our results with an independent GWAS of self-reported stuttering. Our meta-analysis and the self-report GWAS showed high genetic correlation (r_g_ = 0.66). A polygenic score from the self-report GWAS was associated with stuttering in our ascertained cases, with modest predictive value (ΔAUC = 0.067, p = 1.8 × 10⁻²³), and with greater severity and frequency among people who stutter. We replicated two self-report loci (*CAMTA1*, *CTNND2*), and combined analysis identified a genome-wide significant association at *MPPED2*. Our 24 suggestive loci implicated genes with established roles in childhood apraxia of speech, including *SETBP1*, *CAMTA1*, *PTBP2*, *GRIN2A* and *KIRREL3*, with strong enrichment for apraxia genes (OR = 40.1, p = 1 × 10⁻⁴). Within our meta-analysis, persistent stuttering was genetically near-identical to stuttering overall (r_g_ = 0.97). Genetically predicted brain imaging nominally implicated basal ganglia and cerebellar motor circuitry. Common-variant liability to stuttering is shared across ascertainment approaches, relates to severity, and converges with rare-variant genes for speech disorders on circuits underlying speech motor control.

## Introduction

Stuttering is a complex communication disorder characterised by dysfluent speech, with features such as blocks, repetitions, and prolongations. Incidence estimates for stuttering vary; typically ranging from 5–8%,^1^ though a prospective children’s cohort found up to 11% of children experienced stuttering by 4 years of age.^2^ Whilst stuttering resolves for many, approximately one-fifth of children will go on to develop a persistent lifelong stutter.^3^ Sex differences in stuttering incidence and prevalence have also been consistently observed: males are more frequently affected than females, with the male-to-female ratio tending to increase with age, likely reflecting higher rates of recovery in females.^1^

Stuttering interventions to increase fluency, for those who want them, are effective for some children during pre-school years.^4^ However, few effective speech treatments are available for older children, adolescents or adults.^5^ While several clinical characteristics are associated with an increased likelihood of stuttering persistence,^6^ reliably predicting which individuals will continue to stutter remains difficult. Developmental stuttering can shape an individual’s experiences beyond speech, influencing social interactions, and daily life.^7^ People who stutter often report anxiety related to communication situations, and some perceive their speech differences as affecting educational and employment opportunities.^8^ These perspectives highlight the broader social and functional impacts that can accompany stuttering across the lifespan. Some individuals who experience stuttering feel it should be recognised as neurodivergence and would prefer that support is more psychosocial in nature, rather than focused on speech fluency.^9,10^

Identification of genetic factors relevant to stuttering susceptibility and persistence will provide insights into the underlying biology, including the neurological pathways and physiological mechanisms responsible for stuttering. Increased understanding of the causes of stuttering could also allow stratification of stuttering into clinically relevant subtypes, leading to the possibility of more targeted behavioural, psychosocial or pharmacological treatments where indicated.

Estimates for the heritability of stuttering from twin and family studies range from 0.42-0.85, indicating a strong genetic contribution.^11,12^ Family-based studies of persistent stuttering have implicated rare variants in a number of genes, including *GNPTAB, GNPTG, NAGPA*,^13^ *AP4E1,*^14^ *ZBTB20,*^15^ *ARMC3,*^16^ *INFAR1*,^17^ and *PPID.*^18^ A study which utilised a trio-based approach to identify *de novo* variants in 85 children with transient or persistent stuttering, but with no significant family history, found pathogenic, or likely pathogenic, variants in four genes (*SPTBN1, PRPF8, TRIO* and *ZBTB7A*) and identified two further genes with variants of interest (*FLT3* and *IREB2*).^19^ Collectively, these genes are heterogeneous in terms of function, their roles in other traits and disorders, and gene expression patterns in the brain, ^19^ although four *(GNPTAB, GNPTG, NAGPA, AP4E1*) are in lysosomal enzyme-targeting pathways associated with rare storage disorders.^11^ It is also not clear whether rare or common genetic variation in these genes may be relevant to stuttering in the general population.

Genome-wide association studies (GWAS) have begun to explore this question, and multiple common variants associated with stuttering susceptibility have been identified. The International Stuttering Project (ISP)^20^ combined 1,345 clinically ascertained cases, 785 self-reported cases from the Add Health Study^21^ and 14,331 controls in a GWAS which identified a genome-wide significant variant near *SSUH2* and 15 loci at suggestive significance.^20^ Another study investigating predicted stuttering within Vanderbilt’s Electronic Health Records, identified one locus surpassing genome-wide significance near *CYRIA.*^22^ More recently, 57 genome-wide significant loci were identified in a GWAS of 99,776 self-reported stuttering cases (who responded ‘yes’ to the question ‘Have you ever had a stammer or stutter?’), and over 1 million controls (who answered ‘no’) from 23andMe Research Institute data stratified for sex and ancestry.^23^ While the loci identified in these studies did not overlap, polygenic risk scores derived from the larger GWAS of self-reported stuttering significantly predicted stuttering status in both the ISP^20^ and AddHealth^21^ cohorts, indicating that these studies capture a shared underlying genetic architecture despite differences in power and case ascertainment.^23^ A recent review of the field highlights that stuttering is likely influenced by a complex, multifactorial genetic architecture involving both rare and common variation, with phenotypic definition and ascertainment playing a key role in shaping genetic findings and their interpretation.^24^

Here, we undertake a GWAS meta-analysis primarily comprising cases recruited specifically based on stuttering (hereafter ‘ascertained cases’), in addition to cases identified from multiple well-established population-based cohorts. Many of our ascertained cases include measures of persistence into adulthood and quantitative stuttering severity, allowing examination of: (i) associations with lifetime stuttering, (ii) whether loci associated with stuttering onset are also associated with persistence, and (iii) associations with severity. We hypothesised that the more comprehensive phenotyping and targeted ascertainment of our cohort would increase sensitivity to detect variants linked to persistent or severe stuttering.

## Methods

### Ethics

The study was approved by the Human Research Ethics Committee at the Royal Children’s Hospital, Melbourne (approval number 37353). Ethics approvals and participant consent procedures for each contributing cohort are described in the Supplementary Methods.

### Contributing cohorts and phenotype definitions

This study included data from 18 cohorts: the Genetics of Stuttering study (GenStutt),^8^ QSkin Sun and Health Study (QSkin),^25^ MPI Erasmus Genetics of Stuttering (MEGS) Study,^26^ Dutch Individual differences in language skills cohort (IDLaS-NL),^27^ International Stuttering Project (ISP),^20^ Early Language in Victoria Study (ELVS),^28^ the Raine Study,^29^ Australian Twins (AusTwins),^30^ Netherlands Twin Register (NTR),^31^ Twins Early Development Study (TEDS),^32^ TwinsUK,^33^ UK Biobank,^34^ Norwegian Mother, Father and Child Cohort Study (MoBa),^35,36^ Generation R,^37^ 1958 National Child Development Study (NCDS1958),^38^ Avon Longitudinal Study of Parents and Children (ALSPAC),^39,40^ Lifelines,^41^ and Generation Scotland: Scottish Family Health Study (GS:SFHS).^42^

Case-control definitions varied, with full phenotype definitions described in the Supplementary Methods. In brief, the cohorts can be broadly categorised into five groups based on their case identification methods and study designs:

1. Recruited specifically for stuttering (validated self-report): GenStutt and MEGS recruited individuals who self-reported stuttering. In GenStutt, a subset (∼32%) of participants had their self-reports validated through face-to-face assessments by speech-language pathologists.
2. Recruited via speech pathology clinics: The ISP recruited participants through stuttering clinics, ensuring clinical confirmation of stuttering by speech pathologists for all cases.
3. General population cohorts with self or parent report: Several studies, including the Raine Study, AusTwins, NTR, TEDS, TwinsUK, MoBa, Generation R, Lifelines, and GS:SFHS, identified cases through self-reports (in adult cohorts) or parental reports (in child cohorts) of stuttering within their general population samples.
4. General population cohorts with health records: In UK Biobank, cases were identified through stuttering-related codes in primary care records.
5. General population cohorts with mixed definitions: ALSPAC, NCDS1958, ELVs employed mixed methods for case identification, including parental reports, self-reports, and in some instances, expert assessments or quantitative measures of speech.

Control definitions included individuals with no reported history of stuttering, whether by self-report, parental report, or absence in clinical records, depending on the cohort.

Several cohorts, including GenStutt, MEGS, NTR, TwinsUK, NCDS1958, and Lifelines, additionally defined a subset of cases with persistent stuttering, generally characterised by reports of stuttering continuing into adulthood or at the time of assessment.

Contributing general population cohorts were identified via the international GenLang network (https://www.genlang.org/), through personal contacts, or through applications for individual level-data.

### Cohort-level genetic analyses

Cohort-level genotyping platforms, Quality Control (QC) and analyses are fully described in the Supplementary Methods.

Briefly, within each cohort, stringent QC procedures were implemented, typically including exclusion of variants with low call rates, significant deviation from Hardy-Weinberg equilibrium, and low minor allele frequency. Sample-level quality control excluded individuals with high missingness, sex mismatches, non-European ancestries (determined by principal component analysis), and extreme heterozygosity. Genotype imputation was performed using either the Michigan Imputation Server or the Sanger Imputation Service, with the Haplotype Reference Consortium (HRC) r1.1 panel as the primary reference for European populations.

Genome-wide association analyses were conducted separately for each cohort using either SAIGE,^43^ rvtests,^44^ RareMetalWorker,^45^ or Regenie.^46^ All analyses assumed an additive genetic model, and included adjustment for sex and ancestry principal components, with further cohort-specific adjustments, as appropriate. All studies undertook GWAS with stuttering (current or previous) as the primary phenotype. Where sample sizes permitted, secondary GWAS were conducted, including sex-specific analyses and persistent stuttering analyses, in cohorts where this phenotype could be determined.

Sex was defined as genetically-determined sex (inferred from genotype data, e.g. X-chromosome heterozygosity), rather than self-reported gender, across all cohorts and all sex-specific and sex-adjusted analyses in this study.

### Centralised quality control and meta-analysis of genetic associations

Each study’s raw GWAS summary statistics were first reformatted into a standardised form, and derived fields estimated. For each variant, the effective sample size (N_eff_) was calculated as N_eff_ = INFO * 4 / ((1/n_Cases_) + (1/n_Controls_)), and the effective minor allele count (effMAC) was computed as effMAC = MAF * N_eff_ * 2.

Study-level QC was then performed using the EasyX R package. For each study we applied variant level filters, including removal of variants with incomplete information, variants with invalid P (P≤0 or P>1), negative or extreme standard errors (SE ≤0, or ≥10), and large effect estimates (|BETA|≥10). Variants that were monomorphic, had effMAC <10, or INFO < 0.5 were also removed. Each study was then merged with the HRC reference panel; allele flips and mismatches were corrected or removed, and variants with allele-frequency discrepancies >0.1 versus the reference were excluded as outliers.

Harmonised summary statistics were then combined in a fixed-effect meta-analysis using METAL, with study effective sample sizes (N_eff_) used as the weightings. Within each study, variants were restricted to those with effMAC > 20, and genomic control was applied. The meta-analysed results were filtered to include only variants where the combined N_eff_ was at least 70% of the maximum N_eff_, with maximum N_eff_ calculated separately for autosomes and chromosome X. A final round of genomic control was then applied to the meta-analysed test-statistics and P-values. Additionally, we generated combined effect estimates and standard errors using an inverse-variance weighted scheme.

Independent signals were determined using LD-clumping in Plink2, using a p-value cut-off of 1E-5, and r^2^ of 0.1.

Gene-based analyses were performed on the meta-analysed test statistics using MAGMA^47^ v1.10. SNPs were mapped to protein-coding genes using a window extending 20 kb upstream of the transcription start site and 5 kb downstream of the transcription end site. Gene location files (NCBI 37.3 gene definitions) were converted from Entrez to Ensembl gene identifiers using the biomaRt^48^ R package, with missing or duplicate Ensembl IDs removed. Gene-based association tests were conducted using the mean SNP association model, with linkage disequilibrium (LD) estimated from individual-level genotype data from the GenStutt and QSkin cohorts. Gene-level associations were considered statistically significant if they exceeded the Bonferroni-corrected significance threshold, adjusted for the total number of genes tested.

### Mapping loci to genes

We primarily used the FLAMES^49^ annotation and scoring framework to prioritise effector genes at associated loci by integrating gene-based association and tissue enrichment (MAGMA^47^), functional prediction (Polygenic Priority Score; PoPS^50^), and locus fine-mapping (Sum of Single Effects; SuSiE^51,52^).

First, we used MAGMA v1.10 to map SNPs to genes and perform gene-based association analyses as described above, to obtain gene-level z-scores. Tissue enrichment analyses were then conducted using the gene-level Z-scores, and normalised gene expression data from 54 tissues in GTEx v8.^53^

Second, we applied PoPS^50^ v0.2 to compute gene-level polygenic priority scores, which quantify functional convergence across genome-wide association signals. PoPS was run using pre-computed feature matrices provided by the FLAMES authors (https://doi.org/10.5281/zenodo.12635505), together with the MAGMA gene-based association results.

Third, we undertook locus fine-mapping using the susieR^52^ package (v.0.14.2). For each lead SNP, a ±100 kb region was defined, and LD matrices were estimated using the ld_matrix function in the ieugwasr^54^ R package, based on GenStutt and QSkin individual-level genotype data. Fine-mapping was performed using the SuSiE regression with summary statistics^51^ (susie_rss) function, with z-scores as input, the sample size set to the maximum effective sample size (N_eff_), and the maximum number of non-zero effects (L) set to 3. Credible sets were extracted using susie_get_cs, initially applying a purity threshold (min_abs_corr) of 0.1, with a fallback to 0 if no credible sets were returned.

Fourth, FLAMES^49^ annotate was applied to each credible set, integrating PoPS predictions, MAGMA gene-based association and tissue enrichment results, and additional annotation data provided by the FLAMES authors (https://doi.org/10.5281/zenodo.12635505). FLAMES scoring was then used to compute gene-level scores and prioritise the most likely effector gene for each credible set.

In addition, positional mapping was performed using Variant Effect Predictor^55^ (VEP) via the Ensembl GRCh37 REST API to identify the nearest gene, predicted functional consequence, and most damaging variant annotation for each lead SNP.

For each locus, we report the FLAMES-prioritised effector gene where available. Where FLAMES did not prioritise a gene (defined as cumulative precision < 0.75), we report the gene(s) with the highest FLAMES score. In loci where no genes were annotated by FLAMES—either due to complex LD preventing fine-mapping or the absence of genes within 750 kb—the nearest gene based on positional mapping is reported.

### Transcriptome-wide Association Studies

S-PrediXcan^56^ and S-MultiXcan^57^ were used to assess associations between genetically predicted gene expression level and stuttering. Tissue-specific transcriptome-wide association studies (TWAS) were first conducted using S-PrediXcan (v0.80) across 13 brain tissues from GTEx (version 8)^53^, using pre-trained elastic net prediction models, corrected for inflation with variance control^58^. GWAS summary statistics were reformatted into a standardised format and summary statistics were imputed for variants missing in the GWAS, but present in the GTEx reference data. The prediction models were then applied to the harmonised and imputed GWAS summary statistics, with the effective GWAS sample size (N_eff_) and SNP-based heritability specified as parameters. Results from the 13 brain tissues were subsequently combined using S-MultiXcan, which applies multivariate regression to test the joint effects of genetically predicted gene expression across tissues on stuttering. A Bonferroni corrected p-value threshold (p < 4.69 × 10^-6^) was applied.

### GWAS-by-subtraction

The GWAS-by-subtraction framework, implemented in the GenomicSEM R package, was used to decompose genetic liability into two latent factors: a stuttering factor capturing shared genetic liability to stuttering onset, and a “persistent” stuttering factor capturing genetic influences specific to persistence beyond onset. Summary statistics for stuttering and persistent stuttering were harmonised to HapMap3 SNPs and filtered to retain variants with minor allele frequency (MAF) ≥ 0.01 and effective sample size (Neff) ≥ 70% of the maximum Neff. Multivariable LD Score Regression (LDSC) was conducted using European ancestry LD scores and weights to estimate the genetic covariance structure, assuming sample prevalences of 0.07 (stuttering) and 0.04 (persistent stuttering) and population prevalences of 0.11 and 0.02, respectively.

We specified a two-factor genomic structural equation model with unit-variance, orthogonal latent factors. An “onset” factor loaded on both stuttering and persistent stuttering, capturing shared genetic liability to stuttering onset, while a “persistence” factor loaded exclusively on persistent stuttering, capturing genetic liability specific to persistence beyond initial onset. The orthogonality constraint was imposed to ensure that SNP effects on the persistence factor represent genetic influences on persistence independent of liability to stuttering onset, consistent with the logic of GWAS-by-subtraction. Residual variances and residual covariances of the observed traits were fixed to zero. Model parameters were estimated using diagonally weighted least squares (DWLS).

For GWAS-by-subtraction, SNP effects on both latent factors were estimated using GenomicSEM’s userGWAS function applied to harmonised summary statistics aligned to a 1000 Genomes European reference panel (MAF ≥ 0.005). Independent signals for each latent factor were determined using LD-clumping in Plink2, using a p-value cut-off of 1E-5, and r^2^ of 0.1.

### Associations with Stuttering Severity Measures

To examine whether loci associated with stuttering were also associated with stuttering severity and frequency, the 24 lead SNPs from the primary meta-analysis of stuttering were each tested individually in the GenStutt cohort, restricted to individuals with persistent stuttering (n = 951 for severity; n = 480 for frequency).

Stuttering severity (rated 1–10) was analysed as a continuous trait using SAIGE^43^ (docker, version 1.3.0), with adjustment for sex and the first ten ancestry PCs. Stuttering frequency (rated 1–5) was analysed as an ordinal trait using POLMM (Proportional Odds Logistic Mixed Model)^59^, implemented in the R package GRAB (version 0.2.4), with a sparse genetic relatedness matrix constructed using GCTA (version 1.95.0), and adjustment for sex and the first ten ancestry PCs. A Bonferroni-corrected threshold of p < 2.08 × 10⁻³ (0.05/24) was applied.

### Heritability Estimations and Genetic Correlations

LDSC^60^ (v 1.0.1) was used to estimate SNP heritability for all stuttering phenotypes. All LDSC analyses were performed using precomputed linkage disequilibrium scores, calculated using individuals of European ancestries from the 1000 Genomes Project, and were restricted to SNPs included in the HapMap 3 reference panel. Total SNP heritability was estimated on the liability scale using population prevalence estimates based on the Early Language in Victoria (ELVS) prospective cohort study (0.11 for stuttering, 0.09 for female, 0.13 for male and 0.02 for persistent stuttering).^2,61^

Heritability enrichment in genomic annotations was estimated using the S-LDSC framework^62^. We used genomic annotations from the baselineLD model (v2.2) presented in Finucane et al.^63^ which includes 64 main genomic annotations (54 binary; 10 continuous) related to allele frequency, allelic age, linkage disequilibrium, nucleotide diversity, GERP scores, evolutionary conservation, and the age of epigenetic marks, along with 33 flanking regions. For reporting, we restricted to 54 main binary annotations, and significant enrichments were determined at FDR < 5%, using the Benjamini-Hochberg procedure.

We then used S-LDSC to identify trait-relevant cell types or tissues by estimating per-SNP contributions to SNP-heritability, in regions surrounding genes with high tissue-, or cell-specific expression^64^. We used publicly available gene expression datasets, including the Cahoy and GTEx brain gene sets which are specific to the brain, as well as a multi-tissue gene set which contains gene expression profiles for a wide range of tissues. The Cahoy gene set includes data from three brain cell types from mouse, while the GTEx brain gene set comprises 13 brain regions (human samples) derived from the GTEx dataset. The multi-tissue gene set integrates data from the GTEx and Franke laboratory (human, mouse and rat)^65,66^ datasets and includes gene expression profiles from 205 tissues.^64^ Annotation effects were then estimated, using the original LDSC baseline model (v1.2) to estimate regression coefficients (τ), to provide interpretable estimates of tissue-specific contributions to SNP-heritability.

We then used chromatin-based epigenetic data from the Roadmap Epigenomics project to conduct further tissue-type specific analyses. We selected brain-specific chromatin data for 10 brain regions from both adult and fetal samples, which included data on 6 epigenetic markers that annotate active regulatory elements and transcriptional activity. The chromatin data were also jointly modelled with baseline model (v1.2) to estimate regression coefficients (τ). All enrichment analyses underwent false discovery rate (FDR) adjustment using Benjamin-Hochberg approach to account for multiple significance testing.

Genetic correlations between stuttering and related traits were estimated using LDSC.^60^ Traits examined included epilepsies,^67^ migraine-related traits,^68^ reading and language traits,^69^ autism spectrum disorder,^70^ ADHD,^71^ Tourette’s Syndrome,^72^ orofacial cleft^73^, and neurodevelopmental conditions.^74^ Summary statistics for each trait were harmonised to the HapMap 3 reference panel using the LDSC munge function, and the same pre-computed European ancestry LD scores were used as described above. A Bonferroni-corrected threshold of p < 2.63 × 10⁻³ (0.05/19) was applied to account for multiple testing across the 19 traits examined.

### Replication look-ups, and polygenic risk score generation in an independent GWAS of self-reported stuttering

Sex-stratified genome-wide association summary statistics for self-reported ever stuttering were obtained from 23andMe Research Institute for European-ancestry participants. Male and female summary statistics were combined using inverse-variance weighted meta-analysis implemented in METAL, using the inverse-variance weighted scheme with genomic control correction applied prior to meta-analysis (λ_male = 1.017; λ_female = 1.013).^23^

A polygenic risk score was generated using the sex-combined summary statistics using SBayesRC (v0.2.6), which jointly models LD structure and functional genomic annotations.^75^ The European UK Biobank imputed common SNP LD reference panel (∼7 M SNPs)^75^ was used together with the baselineLD model v2.2 (96 functional annotations)^63^. The resulting polygenic scores were then applied to the GenStutt and Qskin cohorts, using individual-level HRC-imputed genotypes. Scores were then standardised to a z-score using the mean and standard deviation estimated in QSkin.

The PRS z-scores were tested for association with stuttering in GenStutt versus QSkin, using logistic regression. The incremental discriminative value of the PRS was quantified as the difference in area under the receiver operating characteristic curve (ΔAUC) between the full model (PRS + covariates) and a covariates-only model (sex and principal components), with a DeLong test used to assess whether this difference was statistically significant.

Within the GenStutt cohort, association between the PRS and stuttering severity was analysed using linear regression, and association with stuttering frequency, coded as an ordered categorical variable, was analysed by ordinal logistic regression, assuming proportional-odds. All models were adjusted for sex and the first 10 ancestry principal components, and restricted to a set of unrelated individuals.

### BrainXcan Analyses

BrainXcan^76^ was used to assess associations between genetically predicted brain image derived phenotypes (IDPs) and stuttering, using both structural (T1-weighted) and diffusion magnetic resonance imaging (dMRIs) measures. The S-BrainXcan framework integrates GWAS summary statistics with pre-computed reference LD data from 1000 Genomes and prediction weights derived from UK biobank’s brain MRI data to impute genetically determined IDPs. Prediction weights were derived using ridge regression models trained on brain IDPs in UK Biobank. Associations between genetically determined IDPs to stuttering were then tested to identify relevant brain-wide and region-specific features. Z-scores were adjusted using variance control, incorporating the effective GWAS sample size (N_eff_) and SNP-heritability^58^.

Structural and diffusion MRI IDPs were further categorised into subtypes pre-defined by the UK Biobank brain image processing pipeline^77^. Structural IDPs from T1 MRIs captured volumetric measures, including cortical grey matter volume, subcortical grey matter volume, subcortical total volume, cerebellum grey matter volume, and brainstem volume. Diffusion MRI features were categorised into four subtypes which describe diffusion properties of water molecules within the local tissue, including fractional anisotropy (FA), intracellular volume fraction (ICVF), isotropic volume fraction (ISOVF) and orientation diffusion index (OD). These diffusion properties were further stratified according to the orientation of estimation, including a voxel-wise approach using tract based spatial statistics (TBSS) and a long-range approach using probabilistic tractography.

### Evolutionary analyses

We estimated heritability enrichment, and undertook gene-set analyses, based on eleven human- and primate-specific evolutionary annotations, capturing accelerated evolution, positive selection, and introgression in humans and primates. We additionally examined gene sets derived from songbird models of learned vocalisation.

We used the S-LDSC framework to estimate heritability enrichment for 6 human and primate-specific annotations that approximately met the common SNP coverage threshold of 0.5%,^78^ including fetal brain human-gained enhancers and promoters at 7, 8.5 and 12 post conception weeks (pcw)^79^, adult brain human-gained enhancers and promoters^80^, ancient selective sweeps^81^ and Neanderthal introgressed alleles^82^. LD scores for these evolutionary annotations were estimated using LDSC (v1.0.1) and jointly modelled with baselineLD model (v2.2)^63^ to control for effect of other genomic annotations MAGMA gene set analysis^47^ was used to examine whether genes located in, or near to each evolutionary annotation set, were more strongly associated with the trait than expected, under a null model of no enrichment. Gene sets were derived for the six annotations above, as well as five additional evolutionary annotations including human accelerated regions (HARs), lineage-specific accelerated regions (linARs)^83^, high confidence HARs (zooHARs)^84^, and human ancestor quickly evolved regions (HAQERs)^85,86^. Evolutionary annotations not originally in GrCh37 coordinates were converted to GrCh37 using the rtracklayer R package (v1.68.0). Each annotation was then mapped to nearby genes using the biomaRt R package (v2.64.0) to identify genes located within 1,000 bases of each annotation. Genes mapped from each annotation were compiled into a single gene-set definition file, which was then used in conjunction with MAGMA gene-level analysis results to estimate regression coefficients for each gene set. An additional gene set analysis was performed for ten songbird vocal gene sets associated with vocal learning phenotypes, as presented in Gordon et al.^87^ Gene sets included: four sets comprised of differentially expressed genes in singing versus silent birds, in four song circuit nuclei (HVC, Area X, the robust nucleus of the arcopallium (RA), and the lateral magnocellular nucleus of the anterior nidopallium (LMAN));^88^ two set containing genes differentially expressed in Area X associated with singing versus silence and motif count, controlling for expression in the ventral striato-pallidum (VSP);^89^ two sets containing genes differentially expressed in auditory brain regions in response to song playback, in females and males separately;^90^ a locomotory behaviour gene set from the Molecular Signatures Database^91^, and a final set comprising all other sets combined.

## Results

Our primary GWAS focused on developmental stuttering (current or previous stuttering), encompassing 6,096 cases and 81,629 controls of European ancestries, from 18 cohorts. 2,953 cases were recruited specifically based on their stuttering (ascertained cases), either via online recruitment (The Genetics of Stuttering Study [GenStutt] and the MPI Erasmus Genetics of Stuttering [MEGS] Study), or via stuttering clinics (ISP). These ascertained case collections were enriched for individuals with persistent stuttering. The remainder of the cases (n=3,143) were identified in population-based studies where case definitions generally relied on self-reported stuttering in adult cohorts or parental reports in child cohorts, with some studies employing more rigorous criteria such as speech assessments or specific stuttering-related questionnaire items. Detailed phenotype definitions and study descriptions are provided in the Supplementary Methods with cohort sample sizes summarised in Table 1.

**Table 1:**
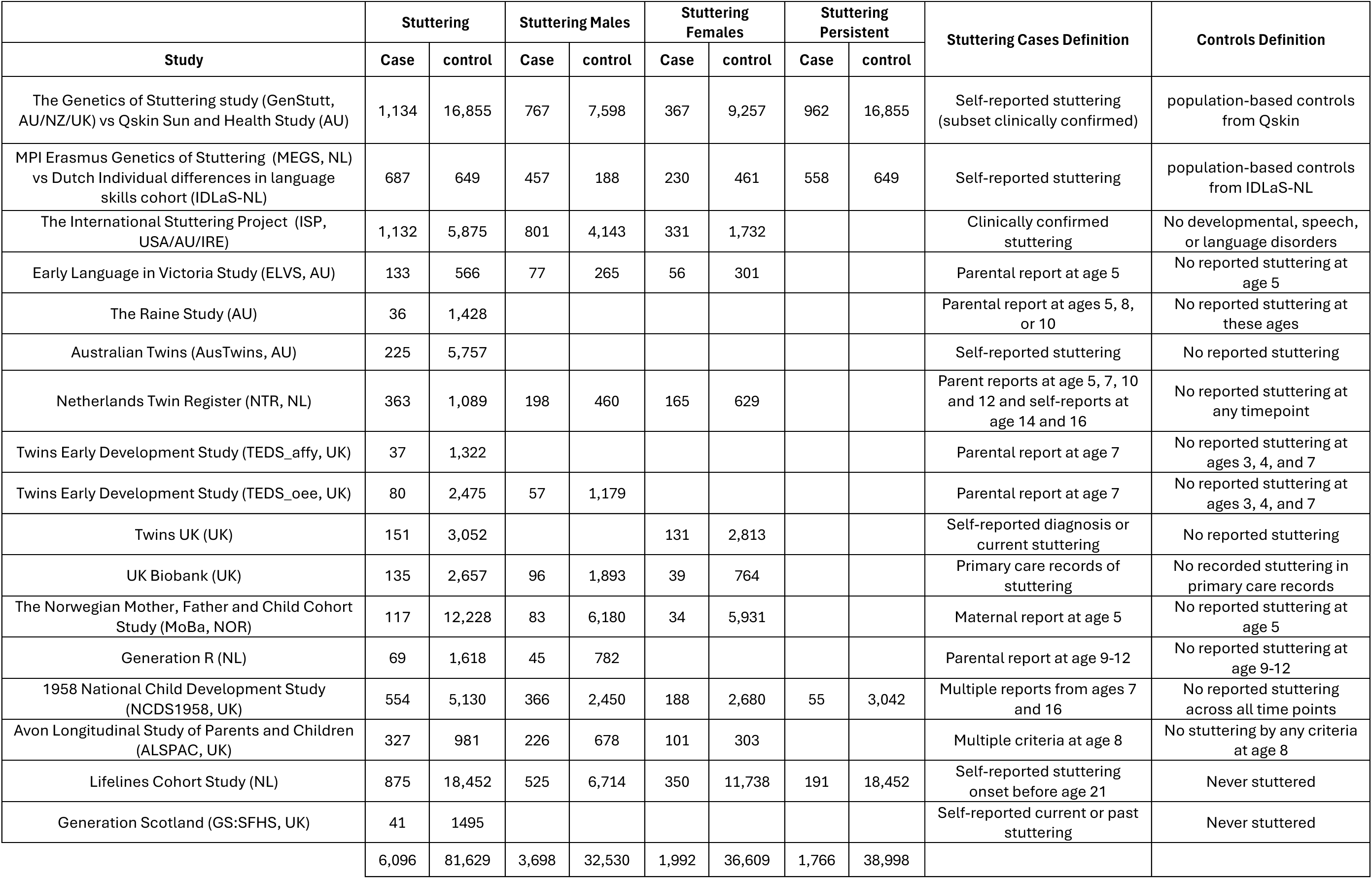
Cohort descriptions for the stuttering GWAS meta-analysis. . Sample sizes and case/control definitions for each cohort contributing to the stuttering GWAS. Columns report case and control counts for stuttering (primary analysis), stutteringMales, stutteringFemales, and stutteringPersistent analyses, alongside the phenotype ascertainment criteria and control definition used in each study.

We also conducted secondary analyses on persistent stuttering (1,766 cases vs 38,998 controls) in a subset of studies, defined as stuttering continuing into adolescence, adulthood or present at the time of assessment (GenStutt). Additionally, we performed sex-stratified analyses for stuttering where sample sizes of contributing cohorts allowed (males: 3,698 cases vs 32,530 controls; females: 1,992 cases vs 36,609 controls).

### Primary GWAS of stuttering

In our primary GWAS meta-analysis of stuttering, no single nucleotide polymorphisms (SNPs) reached genome-wide significance (p < 5 × 10^-8^). However, 24 loci achieved suggestive significance (p < 1 × 10^-5^, Table 2, Figure 1, region plots and forest plots in Figure S1). The top SNPs of these 24 suggestive loci showed consistent direction of effect across sexes, and their effect directions were concordant in the persistent stuttering analysis (Table S1, Figure S2). We mapped the 24 suggestive loci to candidate effector genes using FLAMES (fine-mapped locus assessment model of effector genes)^49^, a machine learning framework that integrates fine-mapped SNP-to-gene annotations with convergence-based evidence from functional gene networks. Where FLAMES did not prioritise a gene, we report the gene with the highest available FLAMES score, or the nearest gene(s).

**Figure 1.**
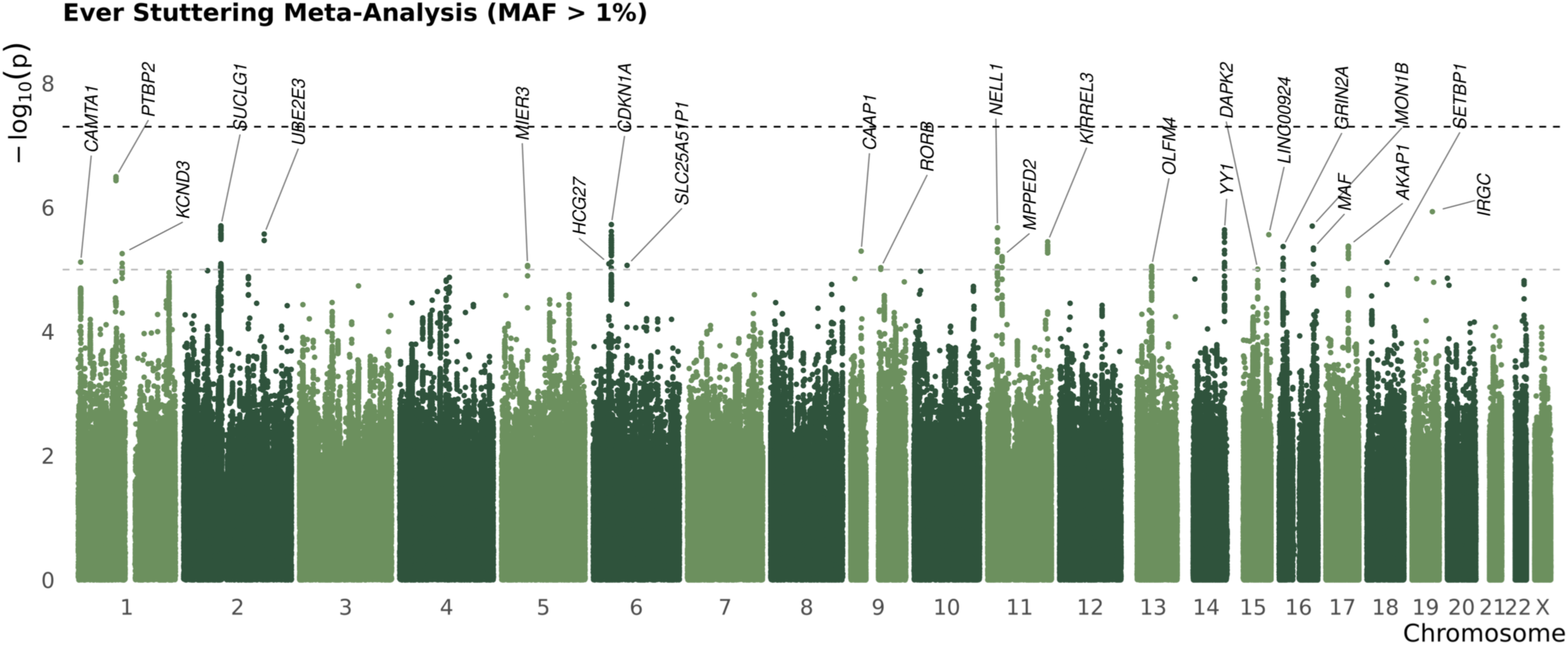
Manhattan plot of genome-wide association results from the primary stuttering GWAS. Each point represents a single nucleotide polymorphism (SNP), plotted by chromosomal position (x-axis) and −log₁₀ transformed p-value (y-axis). The dark dashed line indicates the genome-wide significance threshold (p < 5 × 10⁻⁸) and the light dashed line indicates the suggestive significance threshold (p < 1 × 10⁻⁵). Results are from the primary GWAS meta-analysis comprising up to 6,096 stuttering cases and 81,629 controls.

**Table 2:** 24 SNPs meeting p < 1 x 10^-5^ in the primary GWAS meta-analysis of stuttering (up to 6,096 cases and 81,629 controls). * Gene with the highest FLAMES score reported. Genes in bold were prioritised as an effector gene by FLAMES (defined as cumulative precision ≥ 0.75).

| rsID | Chr | Position (b37) | Effect allele | Other allele | Effect allele frequency | Beta | SE | p-value | Nearest gene | Variant consequence | FLAMES gene* |
| --- | --- | --- | --- | --- | --- | --- | --- | --- | --- | --- | --- |
| rs34204447 | 1 | 7014869 | G | C | 0.048 | 0.234 | 0.052 | 7.57E-06 | CAMTA1 | intron_variant | CAMTA1 |
| rs1359554 | 1 | 97110214 | A | G | 0.687 | -0.113 | 0.022 | 3.18E-07 | PTBP2 | intergenic | PTBP2 |
| rs3005776 | 1 | 112880499 | C | T | 0.396 | -0.095 | 0.022 | 5.53E-06 | CTTNBP2NL | intergenic | KCND3 |
| rs7582221 | 2 | 83924859 | T | C | 0.689 | 0.102 | 0.023 | 1.97E-06 | FUNDC2P2 | intergenic | SUCLG1 |
| rs147710719 | 2 | 181470191 | A | C | 0.039 | -0.282 | 0.061 | 2.68E-06 | UBE2E3 | intergenic | UBE2E3 |
| rs1806672 | 5 | 56275344 | A | G | 0.048 | 0.238 | 0.053 | 8.59E-06 | MIER3 | intergenic | MIER3 |
| rs1793893 | 6 | 31218416 | C | T | 0.224 | 0.116 | 0.027 | 8.07E-06 | HLA-DRB5 | intergenic | HCG27 |
| rs16884857 | 6 | 36070227 | T | G | 0.100 | 0.161 | 0.035 | 1.88E-06 | MAPK14 | intron_variant | CDKN1A |
| rs139878267 | 6 | 67237313 | G | A | 0.022 | 0.356 | 0.082 | 8.55E-06 | SLC25A51P1 | intergenic |  |
| rs12555602 | 9 | 26591043 | G | A | 0.615 | -0.099 | 0.022 | 5.05E-06 | CAAP1 | intergenic | CAAP1 |
| rs78493328 | 9 | 76975416 | T | C | 0.032 | -0.310 | 0.068 | 9.31E-06 | RORB | intergenic | RORB |
| rs34330747 | 11 | 20699315 | G | T | 0.122 | 0.156 | 0.033 | 2.11E-06 | NELL1 | intron_variant | NELL1 |
| rs532395 | 11 | 30450613 | T | C | 0.300 | 0.101 | 0.022 | 6.17E-06 | MPPED2 | non_coding_transcript<br>exon_variant | MPPED2 |
| rs676846 | 11 | 126877714 | G | T | 0.234 | -0.118 | 0.025 | 3.56E-06 | KIRREL3-AS3 | intergenic | KIRREL3 |
| rs7981317 | 13 | 53936098 | A | T | 0.497 | -0.091 | 0.021 | 8.83E-06 | OLFM4 | intergenic | OLFM4 |
| rs3117663 | 14 | 100686402 | G | A | 0.456 | -0.093 | 0.021 | 2.29E-06 | YY1 | intergenic | YY1 |
| rs148952809 | 15 | 64307616 | G | C | 0.031 | -0.308 | 0.069 | 9.83E-06 | DAPK2 | intron_variant | DAPK2 |
| rs1946310 | 15 | 96050882 | A | G | 0.594 | -0.104 | 0.022 | 2.74E-06 | LINC00924 | intergenic |  |
| rs10852372 | 16 | 9306136 | A | C | 0.403 | 0.099 | 0.021 | 4.25E-06 | MIR548X | intergenic | GRIN2A |
| rs62046982 | 16 | 77125221 | C | T | 0.459 | -0.100 | 0.021 | 1.99E-06 | MON1B | intergenic | MON1B |
| rs56243837 | 16 | 79833661 | A | G | 0.045 | 0.240 | 0.052 | 4.50E-06 | MAF | intergenic | MAF |
| rs4383210 | 17 | 55135619 | G | A | 0.773 | -0.118 | 0.026 | 4.19E-06 | RNF126P1 | intergenic | AKAP1 |
| rs6507587 | 18 | 42415449 | G | A | 0.867 | 0.139 | 0.033 | 7.61E-06 | SETBP1 | intron_variant | SETBP1 |
| rs12972416 | 19 | 44220733 | G | A | 0.222 | -0.127 | 0.027 | 1.17E-06 | IRGC | intron_variant | IRGC |

The most statistically significant SNP association was with rs1359554 (effect allele frequency [EAF] = 0.69, beta = -0.11, p = 3.18 × 10^-7^), an intergenic SNP, upstream of *PTBP2*. Prioritised effector genes for other top associations included *IRGC*, *CDKN1A*, *SUCLG1*, and *MON1B*. Notably, 6 of the 24 genes highlighted by these analyses have previously been reported in the severe childhood speech disorder, childhood apraxia of speech (CAS). Three of these genes (*SETBP1,*^92–95^ *CAMTA1,*^92^ and *KCND3*^92^ ) had high-confidence pathogenic variants identified in cohorts of individuals with CAS. A further two (*PTBP2* and *KIRREL3*^94^) had variants initially reported as of uncertain significance or likely pathogenic, with the *PTBP2* variant subsequently reclassified as pathogenic.^96^ Links between CAS and *GRIN2A* are well established.^97–99^ High-confidence CAS genes (pathogenic variants only, n = 67, table S2) were significantly enriched among the 24 GWAS-implicated genes relative to all mapped coding genes (mapped gene count based on the MAGMA analysis; Fisher’s exact test, OR = 40.1, 95% CI 11.7–137.8, p = 1× 10^-4^). Beyond this CAS gene set, *RORB,*^100^ *YY1*^101^ have been implicated in epilepsy and intellectual disability syndromes, respectively, both with concomitant speech and language involvement. Additionally, one gene (*MON1B*) had previously been identified as a candidate gene in a GWAS of individuals with speech sound disorder^102^.

Gene-based analyses using MAGMA^47^ yielded no significant genes, after correction for multiple testing (p < 2.92 × 10^-6^, Table S3). Similarly, transcription-wide association studies undertaken for 13 brain tissues using S-PrediXcan, then combined using multiXcan yielded no genes meeting the Bonferroni corrected p-value threshold (p < 4.69 × 10^-6^, Table S4).

The SNP-based heritability of stuttering on the liability scale was estimated by LD-score regression (LDSC) as h^2^ = 0.26 (95% CI: 0.18-0.34), assuming a population prevalence of 11%. However, we note that heritability estimates were sensitive to prevalence estimates for stuttering, for which there is some uncertainty, and may vary across study populations (Figure S3); given lower prevalences of 2% and 5%, the corresponding heritability estimates would be h^2^ = 0.15 (95% CI: 0.11-0.18), and h^2^ = 0.19 (95% CI: 0.14-0.23), respectively. We undertook partitioned heritability analysis using stratified LDSC (S-LDSC) with the baselineLD model^63^. While no functional category was statistically significant after multiple testing (Figure S4A, Table S5), the highest enrichment estimates were consistently observed in evolutionarily conserved regions (mammalian, primate, and vertebrate *phastCons* elements, and mammalian conserved sequences from Lindblad-Toh et al.^103^). Enrichment was also observed in human promoter-related annotations, and enhancer-histone quantitative trait loci. Although not statistically significant, this pattern suggests that stuttering heritability may be concentrated in highly conserved and regulatory regions of the genome.

### Sex-stratified analyses

Given the well-established sex differences in stuttering prevalence, we additionally undertook GWAS of stuttering in males and females separately to test for sex-specific genetic effects. SNP-based heritability estimates on the liability scale were higher in females (h^2^=0.36, 95% CI: 0.23-0.48, 9% population prevalence) compared to males (h^2^=0.29, 95% CI: 0.20-0.39, 13% population prevalence); however, we note that these estimates have relatively large standard errors, primarily due to limited sample size, particularly for females and again, were sensitive to prevalence estimates (Figure S3). There was a significant positive genetic correlation of r_g_= 0.67 (95% CI: 0.42-0.91), indicating substantial shared, but not identical, genetic architecture between sexes.

The male-specific analysis identified 18 loci reaching p < 1 × 10^-5^ (Table S6, Figure S5A), while the female-specific analysis found 13 such loci (Table S7, Figure S5B). Five loci from the male GWAS (*IRGC, CDKN1A, UBE2E3, LINC00S24, POU5F1*) were also suggestive (p < 1 × 10^-5^) in the sex-combined analysis. The *SUCLG1* locus was also suggestive in both male-specific and sex-combined analyses, but with different lead SNPs (males: rs142012273; EAF = 0.065; beta = 0.29; p = 2.80 × 10^-6^, sex-combined: rs7582221; EAF = 0.69; beta = 0.10; p = 1.98 × 10^-6^). The remaining 12 male-specific and all 13 female-specific loci were not suggestive in the sex-combined analysis.

### Robustness to variation in study design and ascertainment

To assess whether variation in study design and case ascertainment strategies across contributing cohorts influenced our GWAS results, we stratified cohorts into two groups: 1) Case collections specifically ascertained for stuttering, and 2) stuttering cases identified in population-based cohorts.

We meta-analysed these two groups of cohorts separately and compared the effect estimates, finding them to be highly consistent (Table S8, Figure S6). Genetic correlation analysis using LDSC revealed a genetic correlation estimate slightly exceeding 1 (r_g_ = 1.026, 95% CI: 0.615, 1.437), which can occur in LDSC because of sampling variation when the underlying correlation approaches unity. These findings suggest that ascertainment heterogeneity did not substantially bias our GWAS results. Indeed, in the overall stuttering GWAS including all cohorts, none of the lead SNPs of the suggestive loci showed significant evidence (p < 0.05 / 24) of heterogeneity, with nominal (p < 0.05) evidence of heterogeneity for only rs7582221 (*SUCLG1*, I^2 =^ 40.8, p = 0.041, Table S1). Estimated effects were also largely consistent across contributing studies (Figure S1).

### Stuttering persistence

We conducted an additional GWAS of persistent stuttering in 1,766 cases vs 38,998 controls. Although this was a smaller sample size, we hypothesised that focusing on individuals with persistent stuttering may reduce phenotypic heterogeneity from resolved stuttering cases. Our GWAS of persistent stuttering identified 17 loci with p < 1 × 10^-5^ (Table S9, Figure S7), none of which reached this threshold in the primary stuttering GWAS. The top SNP in this analysis was rs10832597 (EAF = 0.027; beta = 0.58; p = 4.51 × 10^-7^), an intronic variant in *SOXC*. The SNP-based heritability of persistent stuttering was estimated using LDSC as h^2^ = 0.27 (se = 0.044), assuming a population prevalence of 2%, with partitioned heritability again highlighting evolutionarily conserved regions (Table S10 and Figure S4B). The genetic correlation between all stuttering and persistent stuttering, estimated via LDSC was very high (r_g_ = 0.97, 95% CI: 0.85, 1.08).

We next applied genomicSEM to perform a GWAS-by-subtraction, which aimed to deconvolve genetic liability into two latent factors: one capturing liability to stuttering onset and the other capturing genetic contributions specific to persistence (Figure S8). In the genome-wide model without individual SNP effects, the stuttering onset factor loaded strongly on both stuttering (standardised loading = 1.00, SE = 0.06, *p* = 1.79 × 10⁻⁵⁹) and persistent stuttering (standardised loading = 0.97, SE = 0.08, *p* = 7.21 × 10⁻³⁶). In contrast, the persistence-specific factor showed a smaller, non-significant loading on persistent stuttering (standardised loading = 0.24, SE = 0.26, *p* = 0.35) and its SNP heritability was not significantly different from zero (h² = 0.19, SE = 0.21). Together these findings provide limited evidence for a distinct common-variant genetic factor specific to stuttering persistence, beyond liability to stuttering onset. A GWAS of the persistent stuttering factor was conducted for completeness, which identified suggestive associations at nine loci with p < 1 × 10^-5^ (Table S11).

Given these findings, the remainder of our analyses focus on results from the primary GWAS of stuttering, which captures the majority of common genetic variation underlying stuttering liability.

### Associations with stuttering severity

We next examined whether the 24 lead SNPs associated with stuttering were also associated with stuttering severity and frequency among individuals with persistent stuttering in the GenStutt cohort. Within this cohort, participants (n=951) provided self-reported stuttering severity ratings, using a scale from 1 (no stuttering in the past week) to 10 (extremely severe stuttering in the past week). A subset (n=480) of participants also reported stuttering frequency on an ordinal scale from 1 (sometimes, not every day) to 5 (several times per sentence). These self-reported measures have been shown to accurately reflect stuttering severity, when compared to speech pathologist ratings.^104^

No SNP was significantly associated with stuttering severity or frequency, after correction for multiple testing (p < 0.05/24). Three SNPs (loci: *NELL1, KIRREL3, YY1*) showed nominal (p<0.05) associations with stuttering severity where the allele associated with increased stuttering susceptibility was associated with increased severity (Table S12, Figure S9). Two further SNPs (*CAMTA1* and *KIRREL3*) showed nominal association (p<0.05) with stuttering frequency; again, the allele associated with increased risk was associated with increased stuttering frequency (Table S13, Figure S9).

### Overlap with an independent GWAS of self-reported stuttering

We performed a look-up within our GWAS results, for SNPs at 23 loci identified in the recent large-scale GWAS of self-reported stuttering in 23andMe (hereafter the self-report GWAS),^23^ which was not a contributing study to our meta-analysis (Table S14). These SNPs were genome-wide significant in either male-only, female-only, or sex-combined analyses in individuals of European ancestries within the self-report GWAS; where a locus was significant in both the sex-combined and sex-stratified analyses but with different top SNPs, the sex-combined top SNP was taken as representative for that locus. Two SNP associations replicated in our GWAS (same direction of effect, with p < 0.05/23 = 2.17 × 10^-3^). The first, near *CAMTA1*, was originally identified in male-specific and sex-combined self-report GWAS (representative top SNP rs12035477). In our study, this SNP met the significance threshold in both female-only (EAF = 0.16; beta = 0.17; p = 2.10 × 10^-4^) and sex-combined analyses (beta = 0.11; p = 2.34 × 10^-5^). The second SNP, mapping to *CTNND2*, was also originally identified in the male-specific and the sex-combined self-report GWAS (rs4331897). This SNP was significant in our female-only analysis (EAF = 0.32; beta = 0.13, p = 4.75 × 10^-4^). Both replicating SNPs showed consistent direction of effect across males and females in our study.

We also tested whether meta-analysing our GWAS results with the sex-combined self-report GWAS^23^ strengthened evidence for any of the 24 suggestive SNPs from the primary meta-analysis of stuttering. We combined summary statistics from our stuttering GWAS and the self-report GWAS for the 22 SNPs present in both datasets (Table S15), and found one SNP, rs532395, reaching genome-wide significance in this combined analysis (beta = 0.034; p = 6.81 × 10^-9^). This intronic SNP in *MPPED2* has previously been associated with reading ability.^105^ SNPs within *MPPED2* have also been associated with migraine with aura.^68,106^

Gene-prioritisation using FLAMES identified two genes, *CAMTA1* and *PTBP2*, that overlapped with mapped genes from the self-report GWAS, using the Open Targets Genetics ‘Variant-to-gene’ (V2G) pipeline. This comparison was performed at the gene level, cross-checking the FLAMES-prioritised gene list from our GWAS against the V2G-derived gene list from the self-report GWAS. For *CAMTA1*, the representative lead SNP from the self-report GWAS was significant in our stuttering GWAS, as noted above; however, our own lead SNP at this locus fell short of the genome-wide significance in the primary stuttering + self-report meta-analysis, though the direction of effect was consistent in the self-report GWAS (rs34204447; EAF = 0.047; beta 0.034, p = 6.03 × 10^-3^). Conditional analysis using GCTA-COJO^107^ suggested the two lead SNPs may tag the same underlying signal (r^2^ ∼ 0.2, D’ = 0.95), leaving it uncertain whether these represent the same or distinct associations. For *PTBP2*, the lead SNPs did not replicate between studies and rs12040559 and rs1359554 are in linkage equilibrium (r^2^ = 3 × 10^-4^, D’ = 0.03), indicating they represent two distinct signals. Despite the two studies identifying different lead SNPs at each locus, the convergence of evidence on *CAMTA1* and *PTBP2* is noteworthy: both genes have been implicated in rare speech disorders,^93,94,96^ and their identification here via an independent GWAS of self-reported stuttering provides additional support for their involvement in speech-related disorders more broadly.

Finally, we took a genome-wide approach to assess overlap between our meta-analysis and the self-report GWAS^23^. Using LDSC, we estimated a strong and statistically significant genetic correlation between the self-report GWAS^23^ and our meta-analysis (r_g_ = 0.66, SE = 0.049, p = 2.47×10⁻⁴⁰, Figure S10), supporting substantial shared genetic architecture between the two studies. We next generated a polygenic risk score (PRS) from the self-report GWAS summary statistics^23^ using SBayesRC^75^, and tested this for association with stuttering in GenStutt + QSkin. The PRS was strongly associated with stuttering (beta = 0.62, p = 3.31 × 10⁻^78^, Figure 2A), with an improvement in AUC of 0.067 (p = 1.78×10⁻^23^), compared to the baseline model of sex and ancestry PCs only. Individuals in the top PRS decile had 2.57-fold higher odds of stuttering compared to those in the 5th (reference) decile (OR=2.57, 95% CI 2.01–3.28, p=5.63×10⁻¹⁴, Figure 2B). Within GenStutt, the PRS also showed association with both stuttering severity (beta = 0.20, p = 4.92 x 10⁻³, Figure 2C) and stuttering frequency (beta = 0.23, p = 0.019, Figure 2D), with higher PRS associated with greater severity and frequency.

**Figure 2:**
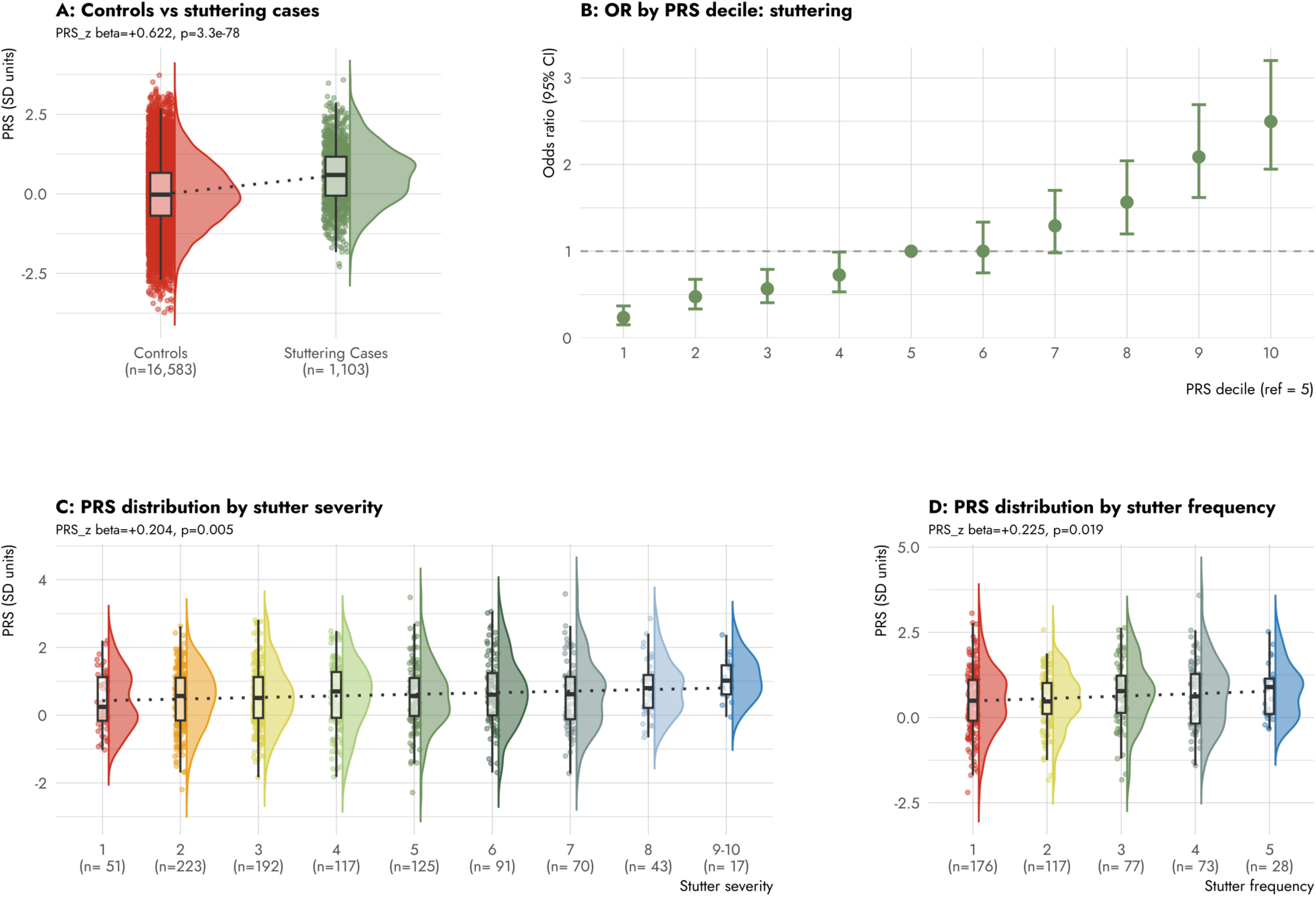
Associations between Polygenic Risk Scores derived from an independent GWAS of self-reported stuttering, with stuttering phenotypes in GenStutt. PRS were constructed from the summary statistics of the sex-combined GWAS of self-reported ever-stuttering and standardised to z-scores using the mean and SD of unrelated QSkin controls; analyses were restricted to unrelated individuals, and sex and 10 ancestry PCs were included as covariates throughout. (A) PRS in QSkin controls versus GenStutt stuttering cases as a raincloud plot, with the dotted line showing the linear trend across groups. Beta and p-value from a logistic regression model. (B) Odds ratio (95% CI) for ever stuttering across PRS deciles, adjusted for sex and PCs, relative to the fifth (middle) decile (plotted at OR = 1, no CI). (C) PRS by self-reported stuttering severity, rated on a scale from 1 (no stuttering in the past week) to 10 (extremely severe stuttering in the past week), with a dotted line showing the linear trend across categories. Beta and p-value from a linear regression model. (D) PRS by self-reported stuttering frequency, rated on an ordinal scale from 1 (sometimes, not every day) to 5 (several times per sentence), with a dotted line showing the linear trend across categories. Beta and p-value from an ordinal regression model.

### Genetic correlations

Several of the genes highlighted through our GWAS have been implicated in childhood apraxia of speech, neurodevelopmental conditions and epilepsies, largely based on rare pathogenic variants, while the *MPPED2* locus, which replicated in the independent self-reported stuttering GWAS, has also been associated with migraine (with aura), and reading ability.

We therefore sought to further examine whether there is a shared genetic basis between stuttering and these, as well as other related traits, by examining genome-wide genetic correlations using LDSC. After accounting for multiple testing across the 19 traits tested (p < 0.05/19 = 2.6 ×10⁻³), a significant negative genetic correlation was identified with genetic generalised epilepsy (r_g_ = -0.19, SE = 0.06, p = 0.002; Table S16, Figure S10). A nominal negative correlation was also observed with phoneme awareness (r_g_ = -0.20, SE = 0.10, p = 0.032), though this did not survive correction for multiple testing.

### Highlighting brain regions with relevance to stuttering

To identify brain regions with potential relevance to stuttering disposition, we integrated GWAS results with brain-specific gene expression and chromatin annotations using S-LDSC, and tested associations between genetically predicted brain imaging–derived phenotypes (IDPs) and stuttering using BrainXcan.

In our S-LDSC analyses, nominal enrichment of SNP-heritability (p < 0.05) was found in several brain-related annotations, although none was significant after correction for multiple testing (FDR < 5%, Table S17, Figure S11). Gene-expression–based partitioning using the multi-tissue gene expression datasets, showed the strongest enrichment across brain tissues, with the largest coefficients observed for frontal lobe (p = 1×10⁻³), parietal lobe (p = 2×10⁻³), and brain overall (p = 5×10⁻³). In the brain-only tissue specific analysis^64^ only the cerebellar hemisphere was nominally significant (p = 0.05), with the next highest, non-significant enrichments observed for the cortex (overall), amygdala and anterior cingulate cortex. Cell-type–specific analyses found no significant enrichment for any cell-type.

Partitioning based on chromatin features (Table S18, Figure S12) revealed nominally significant enrichments in fetal brain regulatory elements, particularly female fetal DNase hypersensitivity sites (p = 0.01), H3K4me1 enhancer marks (p = 0.02) and H3K4me3 promoter marks (p = 0.04), consistent with a role for open chromatin and active regulatory elements during early neurodevelopment. Additional nominal enrichment (p < 0.05) was detected in adult cortex, including H3K9ac marks in the dorsolateral prefrontal cortex and H3K27ac marks in the cingulate gyrus.

BrainXcan analysis identified multiple genetically predicted brain IDPs nominally associated with stuttering, spanning both structural MRI and diffusion MRI modalities (Table S19, Figure 3, Figures S13-14); however, none were significant after correction for multiple testing (FDR < 5%). The strongest nominal associations related to subcortical motor regions, most notably decreased grey matter volume in the pallidum. Diffusion MRI associations clustered within major white-matter projection pathways, particularly the external capsule, with decreased isotropic volume fraction (ISOVF), and the anterior and posterior limbs of the internal capsule (bilaterally), and the fornix (cres)/stria terminalis (left), where associations with increased intracellular volume fraction (ICVF) were observed. Nominal associations with increased fractional anisotropy (FA) were also observed for the anterior limb of the internal capsule, but not the posterior. Cerebellar involvement was also highlighted, with increased grey matter volume in lobule VI and increased orientation dispersion in the superior cerebellar peduncle, a principal cerebellar output tract.

**Figure 3:**
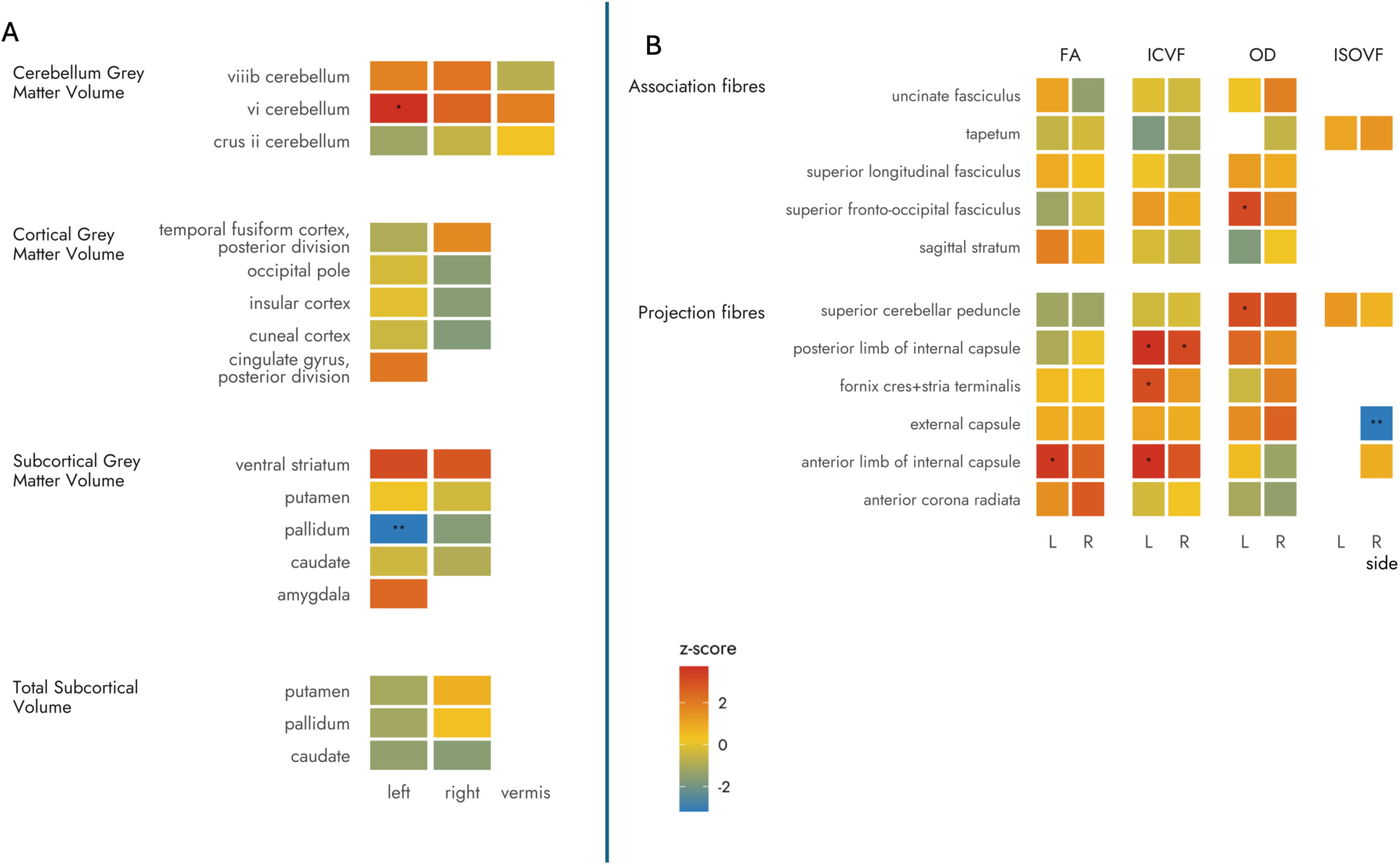
Partial heatmaps of S-BrainXcan association analyses for: A. structural (T1) brain magnetic resonance imaging (MRI) features in stuttering; B. diffusion magnetic resonance imaging (dMRI) features in stuttering by white matter fibre type. White matter tracts were categorised into association fibres, which connect brain regions within the same hemisphere, and projection fibres, which connect the cerebral cortex to subcortical structures. Z-scores were adjusted for variance control, with GWAS trait heritability of 0.24 and GWAS effective sample size of 11,348. * = p value < 0.05. ** = p value < 0.01.

### Evolutionary and Comparative Genomic Enrichment Analyses

Given that fluent speech is a recently evolved human trait and relies on neural circuits that are partially conserved across species, we next investigated whether genetic susceptibility to stuttering is enriched in annotations with relevance to hominin evolution, or in gene sets linked to vocal learning. We focused on annotations capturing accelerated evolution, positive selection, and introgression in humans and primates, and additionally examined gene sets derived from songbird models of learned vocalisation, which provide a well-established comparative framework for studying the genetic architecture of complex vocal behaviours^108^ (annotations summarised in Table S20).

We first used S-LDSC to test for enrichment of SNP-heritability in a set of human- and primate-focused evolutionary annotations that met the ≥0.5% common SNP coverage threshold. These included fetal brain human-gained enhancers and promoters at 7, 8.5 and 12 post-conception weeks,^79^ adult brain human-gained enhancers and promoters,^80^ ancient selective sweeps,^109^ and Neanderthal introgressed alleles.^82^ When jointly modelled with the baselineLD model, none of these annotations showed significant enrichment of SNP-heritability (Table S21, Figure 4A).

**Figure 4:**
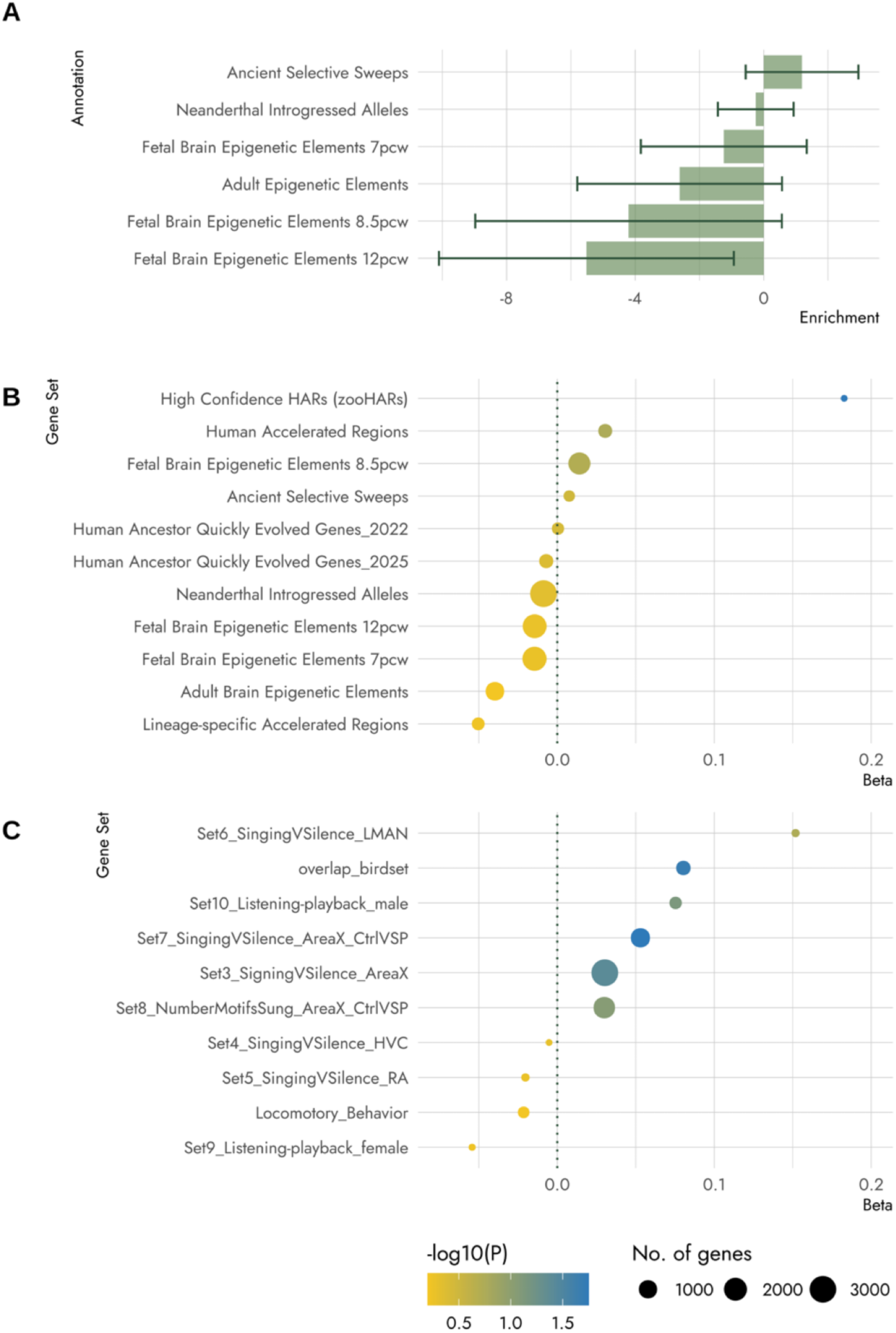
Heritability enrichment for stuttering with respect to evolutionary annotations and genes associated with songbird vocal behaviours. Heritability enrichment was estimated using **(A)** SLDSC for six evolutionary annotations approximately meeting the 0.5% SNP proportion threshold. Error bars indicate the standard error of the enrichment. MAGMA gene set analyses were used to estimate regression coefficients for **(B)** eleven evolutionary annotations and **(C)** ten songbird vocal gene sets. P-values from a one-sided test that the beta coefficient is greater than zero.

We next performed MAGMA gene-set enrichment analyses for genes mapped to these annotations, in addition to genes mapped to human accelerated regions (HARs)^110^, lineage-specific accelerated regions (linARs),^83^ high confidence HARs defined from the Zoonomia 241-mammal alignment (zooHARs),^84^ and two sets of human ancestor quickly evolved regions (HAQERs).^85,86^ No gene sets were significantly enriched after correction for multiple testing; however, a nominally positive enrichment was observed for zooHARs (beta = 0.18, p = 0.014, Table S22, Figure 4B).

Finally, we evaluated nine gene sets derived from songbird models of vocal learning, mapped to their human homologues. These gene sets capture differential gene expression patterns associated with vocal learning behaviours across core components of the songbird song system, including Area X, HVC (used as proper name), the lateral magnocellular nucleus of the anterior nidopallium (LMAN) and the robust nucleus of the arcopallium (RA)^87–90^. Nominal evidence of enrichment in MAGMA was observed for three gene-sets related to Area X, a basal ganglia nucleus crucial for vocal learning (Table S23, Figure 4C). Behavioural phenotypes related to these Area X gene-sets included singing versus silence (p = 0.040); singing versus silence, controlling for differential expression of genes in the ventral striato-pallidum (p = 0.018); and an overlapping gene set incorporating these sets together with number of motifs sung (p = 0.021). None of these songbird-derived gene sets remained significant after correction for multiple testing.

## Discussion

We performed a GWAS meta-analysis of stuttering, combining data from across 18 cohorts (6,096 cases, 81,629 controls), with complementary secondary analyses of persistent stuttering and sex-stratified GWAS. While no variants reached genome-wide significance in the primary stuttering GWAS, 24 loci showed suggestive association (p < 1 × 10^-5^), with the most significant single-variant association being rs1359554 (p = 3.18 × 10^-7^), an intergenic variant upstream of *PTBP2*. SNP-based heritability for stuttering was moderate (h^2^ = 0.26). Gene-prioritisation highlighted a number of genes with known roles in neurodevelopmental conditions and epilepsies, which include a speech-related phenotype (*RORB*^100^*, YY1*^101^), and childhood apraxia of speech (*SETBP1,*^92–95^ *CAMTA1,*^92^ *KCND3,*^92^ *PTBP2,*^94,96^ *GRIN2A,*^94,97,98^ and *KIRREL3*^94^). This supports the finding of a stuttering phenotype in an individual with SETBP1 haploinsufficiency.^111^ Of note, genes implicated in childhood apraxia of speech were significantly enriched among effector genes at suggestive loci overall (p = 1 × 10^−4^). Genome-wide genetic correlations with neurodevelopmental and language-related traits were not significant after correction for multiple testing, with the exception of genetic generalised epilepsy, which showed a significant negative correlation (p = 2× 10^−3^). A nominal negative correlation was also observed with phoneme awareness (p = 0.032).

Sex-stratified analyses indicate substantial shared, but non-identical, genetic architecture between males and females. Although we identified multiple suggestive loci in the sex-specific GWAS, effect estimates at primary analysis loci were broadly consistent across sexes, and the significant male-female genetic correlation (r*_g_* = 0.667, p = 1.45 × 10⁻⁷) suggests that most common variant risk is shared. These results are broadly consistent with the recent sex-stratified analysis of self-reported stuttering in 23andMe^23^; in that study, genetic correlation between sexes was estimated to be higher than in our study, yet there was no overlap in genome-wide significant loci between the European male and female analyses. Analyses targeting persistence failed to provide convincing evidence for a distinct common-variant genetic component beyond general susceptibility. Together, these results imply that much larger samples with deep phenotyping will be needed to resolve with GWAS whether true genetic differences exist between sexes or between resolved versus persistent stuttering.

We cross-checked the findings from our primary GWAS against an independent GWAS of self-reported stuttering, which provided corroboration for three loci (*MPPED2; PTBP2; CAMTA1)* at either the SNP or gene level, and additionally provided independent replication of the *CTNND2* association, first identified in the self-report GWAS. Corroboration at *MPPED2* was at the SNP level, with rs532395 reaching genome-wide significance in a combined meta-analysis with the self-report GWAS. *MPPED2* encodes a metallophosphoesterase that is predominantly expressed in fetal brain and may play a role in development of the nervous system.^112^ GWAS Catalog associations mapped to *MPPED2* are dominated by renal function traits,^113,114^ with additional associations involving blood pressure,^115^ anthropometric measures,^116^ and neurological and behavioural traits, including risk-taking behaviour,^117^ morningness,^118^ migraine with aura,^68^ reading ability,^105^ and cortical surface area.^119^

At the gene level, genes prioritised from our GWAS (*CAMTA1* and *PTBP2*) overlapped with genes mapped to loci from the independent self-report GWAS, providing convergent support for their involvement in stuttering. Notably, both these genes are amongst those previously implicated in severe speech disorders.^94,120^ *PTBP2* encodes a neuron-specific RNA-binding protein that regulates alternative splicing during neuronal differentiation and early brain development, with expression at highest levels prenatally and declining postnatally as neurons mature.^121^ Common variation at *PTBP2* has been linked to cognitive and psychiatric traits including schizophrenia,^122^ educational attainment,^123^ and reaction time^124^, in addition to body mass index^125^. *CAMTA1* encodes a calcium-responsive transcription factor, expressed in the brain, and thought to regulate neuronal development, maturation, and survival, particularly in cerebellar regions.^120^ Rare protein-altering variants in *CAMTA1* cause cerebellar dysfunction with variable cognitive and behavioural abnormalities (CECBA), a heterogeneous disorder, with most individuals showing global developmental delay, particularly of motor and language skills, evolving to mild intellectual disability, and movement disorder.^120^ Common variants mapped to *CAMTA1* have been associated with cognitive and neuropsychiatric phenotypes, consistent with roles in motor coordination and higher-order brain function.

Despite limited replication at the level of individual SNPs and genes between our primary GWAS and the independent GWAS of self-reported stuttering, the strong genome-wide genetic correlation (r_g_ = 0.66) and predictive utility of the self-report GWAS PRS (AUC = 0.73) indicate substantial shared genetic architecture between the two studies. Notably, the PRS was also associated with stuttering severity and frequency within GenStutt, suggesting that genetic liability identified through case-control GWAS may extend to variation in symptom severity among those who stutter.

We sought to identify brain regions most relevant to genetic susceptibility to stuttering via complementary approaches: testing for enrichment in genome-wide annotations (S-LDSC) and genetically predicted imaging analyses (BrainXcan). S-LDSC analyses suggested tissue-level enrichment in the frontal and parietal cortex and nominal enrichment in cerebellar annotations and fetal brain regulatory elements, while BrainXcan suggested decreased pallidum grey matter volume, increased grey matter in cerebellar lobule VI and altered microstructure of major white matter tracts. Anomalies of the pallidum, a major component of the subcortical basal ganglia, have been observed in a large family with inherited stuttering, associated with a *PPID* variant,^18,126^ though we found no evidence of association with *PPID* in our GWAS. Our BrainXcan analyses also highlighted similar brain regions to an analogous analysis using the GWAS of self-reported stuttering^127^: these two independent analyses showed consistent patterns for cerebellar lobule VI (increased grey matter volume in both analyses) and the external capsule (decreased ISOVF and increased FA, both consistent with reduced free water and greater tissue organisation), but a less readily reconciled pattern for the fornix (cres)/stria terminalis, where increased ICVF in our analysis and increased ISOVF in the analysis based on the self-report GWAS pointing to differing interpretations. Our findings are also broadly consistent with an imaging study which found functional and structural differences in cortical motor/premotor regions, the basal ganglia, cerebellum, and white matter tracts in a cohort of people who stutter.^128^

Our partitioned heritability analysis suggested that stuttering heritability was enriched in deeply conserved genomic and regulatory regions. This pattern suggests that genetic contributors to stuttering may lie in evolutionarily ancient biological pathways, potentially involving core neural or motor-control mechanisms shared across vertebrates. Further S-LDSC and MAGMA gene-set analyses leveraging human- and primate-specific evolutionary annotations did not show evidence of enrichment for more recently evolved regions, although a nominal positive signal was observed for zooHARs, a set of regions highly conserved across mammals, but with a rapid rate of substitutions specifically in the human lineage.^84^ Gene-set analyses related to songbird derived vocal-learning^87^ showed weak enrichment in several Area X (basal-ganglia/vocal-learning) gene sets, consistent with findings from a multivariate GWAS of rhythm impairment and dyslexia.^129^ Overall, our findings suggest that stuttering risk variants are preferentially located in conserved regulatory elements and may implicate genes related to vocal learning, across species.

Our study had several limitations. First, despite combining multiple case collections and large population-based cohorts, our sample size remained modest for a GWAS-based approach, limiting power to detect small-effect associations. This partly reflects the difficulty of identifying stuttering cases within population-based cohorts given this trait is rarely included. While as many as 11%^2^ of individuals experience stuttering at some time in their life, substantially lower proportions seem to report stuttering (current, or past) in population studies, potentially leading to misclassification and attenuating genetic associations. However, within the GenStutt cohort, self-reported stuttering showed strong concordance with speech pathologist assessment,^8,104^ lending confidence to the self-report measures used across our cohorts. Second, we were unable to replicate several findings in the independent, self-reported stuttering GWAS, potentially reflecting heterogeneity in ascertainment or phenotype definitions, but this may also be due to limited power in our analyses. Third, our secondary analyses examining sex-specific effects and persistent stuttering did not provide convincing evidence for differences between males and females or a distinct genetic architecture underlying persistence. These null findings may reflect insufficient power rather than true biological homogeneity.

More broadly, phenotypic information on stuttering, and other speech and language traits, remain limited in most population-based studies. We advocate greater focus on collecting speech and language phenotypes in large-scale cohort studies and biobanks. Detailed collection of these traits in emerging studies would enable more nuanced genetic investigations in the future and should be included in data collection from the outset.^130^ Finally, we restricted analyses to individuals of European ancestry due to insufficient power to include non-European populations. Future research with deeper phenotyping in ancestrally diverse populations is essential to ensure greater generalisability of genetic discoveries.

Our findings provide convergent evidence that genetic susceptibility to stuttering involves neurodevelopmental pathways relevant to speech and motor control. The overlap with genes implicated in childhood apraxia of speech, together with converging imaging analyses, highlights biologically plausible mechanisms underlying speech fluency.

## Data Availability

Genome-wide summary statistics for the primary GWAS of stuttering, and all secondary analyses (persistent stuttering, and sex-stratified) will be made available via the GWAS catalog, on publication.

## Contributions

Formal Analysis: V.E.J., J.J.S., E.E., J.E.B., D.I.B., E.B., S.D.G., K.M.K., G.H.M., H.S.M., A.M., H.G.P., R.Pool, V.A.R., A.C.S., C.A.W.; Phenotyping: A.T.M., S.H., J.O.B., O.v.R., J.B., J.E.B., E.v.B., D.I.B., M-C.J.F., A.H., S.P.C.K., S.J., M.L., A.M., C.E.P., V.A.R., K.Z.V., A.J.O.W., Y.W., S.R.; Supervision: J.E.B., E.v.B., D.I.B., A.H., M.L., C.E.P., R.Pool, K.R., A.J.O.W., L.S., F.L., S.R., S.E.F., N.G.M., I.E.S., A.T.M., M.B.; Project Administration/Recruitment: V.E.J., A.T.M., S.H., J.O.B., E.E., R.Parker, O.v.R., D.T-L., M.E., E.v.B., D.I.B., A.H., M.L., H.S.M., C.E.P., R.Pool, B.StP., S.B.V., Y.W., S.E.F.; Resources/Funding Acquisition: J.E.B., E.v.B., D.I.B., E.C.C., A.H., S.J., M.L., C.E.P., R.Pool, A.C.S., S.B.V., N.A.G., A.V., I.E.S., M.S.H., L.S., F.L., S.R., S.E.F., N.G.M., A.T.M., M.B. Writing: V.E.J., J.J.S., A.T.M. and M.B. drafted the manuscript. All authors provided comments and feedback.

## Declaration of Interests

The authors declare no competing interests relevant to this study.

## Acknowledgements

We are extremely grateful to all the families and participants who took part in this study across all contributing cohorts. Their continued engagement and willingness to contribute data made this research possible.

During the preparation of this manuscript, Claude (Anthropic) was used to improve the readability, clarity, and conciseness of the text. After using this tool, the authors reviewed and edited the content as needed and take full responsibility for the final content of the publication.

## Funding

This work was supported by the National Health and Medical Research Council (NHMRC) Centre of Research Excellence in Speech and Language (SLANG) grant no. 1116976 awarded to A.T.M., M.B., I.E.S., M.H., S.R., S.E.F., F.L. and project grant no. 1160893 awarded to A.T.M., M.B., S.R., S.E.F., V.E.J.. Australian National Health and Medical Research Council (NHMRC) Investigator Grants were awarded to M.B. (GNT1195236), A.T.M. (GNT 1195955, 2041068) and I.E.S. (GN1172897T, GNT2033247). This work was made possible through the Victorian State Government Operational Infrastructure Support Program and the Australian Government NHMRC IRIISS.

B.St.P., E.E., I.A.S and S.E.F were supported by the Max Planck Society. E.E. was also supported by a Veni grant from the Dutch Research Council (NWO; VI.Veni.202.072). I.A.S. was also supported by the Dutch Research Council (Language in Interaction consortium: Gravitation grant number 024.001.006). In addition, B.St.P. was supported by the R2D2-MH grant funded by Horizon Europe (grant agreement no. 101057385), by UK Research and Innovation (UKRI) under the UK government’s Horizon Europe funding guarantee (grant no. 10039383), by the Swiss State Secretariat for Education, Research and Innovation (SERI) under contract number 22.00277, and by the Radboud University Donders RSF 2026. Y.W. was supported by The Underwood Trust.

K.R. is supported by UK Medical Research Council Grant UKRI1503. H.S.M. was supported by the Biotechnology and Biological Sciences Research Council [BB/T000813/1]. E.B. was funded by a PhD studentship from the Economic and Social Research Council with the Scottish Graduate School of Social Sciences.

E.v.B. is funded by the European Union (ERC, Project "InterGen", PI: E. van Bergen, 101076726), the Research Council of Norway (Project "GenEd", PI: M. Melby-Lervåg, 335634), the Dutch Research Council NWO (VENI and VIDI Talent Grants, PI: E. van Bergen, 451-15-017; VI.Vidi.221G.007), and she is a Jacobs Foundation Research Fellow.

D.I.B. acknowledges a Royal Netherlands Academy of Science Professor Award (PAH/6635) and funding from the Dutch Research Council NWO: Biobanking and Biomolecular Research Infrastructure (BBMRI–NL, 184.033.111) and the BIOS Consortium (NWO 184.021.007 & 184.033.111); Aggression in Children: Unravelling gene-environment interplay to inform Treatment and InterventiON strategies project (ACTION) funded by the European Union Seventh Framework Program (FP7/2007-2013) under grant agreement no. 602768; European Research Council (ERC-230374, Genetics of mental illness); NWO/SPI 56-464-14192.

G.H.M. is funded by the Research Council of Norway (Project grant: 325640). A.H. is supported by the Research Council of Norway (#336085) and the South-Eastern Norway Regional Health Authority (#2020022; #2026069). E.C.C. was supported by the Research Council of Norway (#274611) and South-Eastern Norway Regional Health Authority (#2021045) and is a member of the MRC Integrative Epidemiology Unit at the University of Bristol, which is supported by the Medical Research Council and the University of Bristol (MC_UU_00032/1). S.B.V. was supported by the South-Eastern Norway Regional Health Authority (#2022029).

This research was supported by National Institutes of Health (NIH) grants from the National Institute on Deafness and Other Communication Disorders (NIDCD) to Vanderbilt University Medical Center and Wayne State University (1R03DC015329 and R01DC017175) and to Vanderbilt University Medical Center (5R21DC016723 and R01DC020311). A.C.S. received funding from the NIDCD under award number F31DC022482.

Cohort-specific acknowledgments:

Genetics of Stuttering Study: We thank the Genetics of Stuttering Study participants and their families for their involvement in this study.

ALSPAC: We are extremely grateful to all the families who took part in this study, the midwives for their help in recruiting them, and the whole ALSPAC team, which includes data collection staff, data and administration staff, technical managers, and the technical staff at the Bristol Bioresource Laboratory, based within the University of Bristol. We are particularly grateful to Dr Rosemarie Hayhow and Professor Sue Roulstone for their work on identifying children with a stutter in the cohort. The UK Medical Research Council and Wellcome (Grant ref: MR/Z505924/1) and the University of Bristol provide core support for ALSPAC. GWAS data was generated by Sample Logistics and Genotyping Facilities at Wellcome Sanger Institute and LabCorp (Laboratory Corporation of America) using support from 23andMe.

Generation Scotland: Generation Scotland received core support from the Chief Scientist Office of the Scottish Government Health Directorates [CZD/16/6] and the Scottish Funding Council [HR03006] and is currently supported by the Wellcome Trust [216767/Z/19/Z]. Genotyping of the GS:SFHS samples was carried out by the Genetics Core Laboratory at the Edinburgh Clinical Research Facility, University of Edinburgh, Scotland, and was funded by the Medical Research Council UK and the Wellcome Trust (Wellcome Trust Strategic Award "STratifying Resilience and Depression Longitudinally" (STRADL) Reference 104036/Z/14/Z). Phenotype collection was supported by the Biotechnology and Biological Sciences Research Council [BB/T000813/1]. We would like to thank all the Generation Scotland participants who gave up their time to take part in our study, the GS:SFHS staff, and the telephone interviewers for their hard work on this project.

Dutch Individual differences in language skills cohort: we would like to acknowledge our project partners Florian Hintz and Antje S. Meyer. The cohort was funded by the Dutch Research Council (NWO), Gravitation grant ‘Language in Interaction’ (grant number 024.001.006) and by the Max Planck Society.

International Stuttering Project: We thank participants from the International Stuttering Project.

Lifelines: The Lifelines initiative has been made possible by subsidy from the Dutch Ministry of Health, Welfare and Sport, the Dutch Ministry of Economic Affairs, the University Medical Center Groningen (UMCG), University of Groningen and the Provinces in the North of the Netherlands (Drenthe, Friesland, Groningen).

MEGS: We are very grateful to all people who stutter who participated in the Max Planck Institute Erasmus Genetics of Stuttering study.

MoBa: This study includes data from the Norwegian Mother, Father and Child Cohort Study (MoBa) conducted by the Norwegian Institute of Public Health. This work was performed on the Tjeneste for Sensitive Data (TSD) facilities, owned by the University of Oslo, operated and developed by the TSD service group at the University of Oslo, IT-Department (USIT), using resources provided by Sigma2—the National Infrastructure for High Performance Computing and Data Storage in Norway (UNINETT). For generating high-quality genomic data, we thank the Norwegian Institute of Public Health (NIPH), the HARVEST collaboration, the NORMENT Centre at the University of Oslo, the Center for Diabetes Research at the University of Bergen, deCODE Genetics, the Research Council of Norway, the SouthEastern and Western Norway Regional Health Authorities, the ERC AdG, Stiftelsen KG Jebsen, the Trond Mohn Foundation, and the Novo Nordisk Foundation. We are grateful to all the participating families in Norway who take part in the ongoing MoBa cohort study.

NCDS: This work made use of data and samples generated by the 1958 Birth Cohort (NCDS), which is managed by the Centre for Longitudinal Studies at the UCL Institute of Education, funded by the Economic and Social Research Council (grant number ES/M001660/1). Access to these resources was enabled via the Wellcome Trust and MRC: 58FORWARDS grant [108439/Z/15/Z] (The 1958 Birth Cohort: Fostering new Opportunities for Research via Wider Access to Reliable Data and Samples). Before 2015, biomedical resources were maintained under the Wellcome Trust and Medical Research Council 58READIE Project (grant numbers WT095219MA and G1001799). We are grateful to the Centre for Longitudinal Studies (CLS), UCL Social Research Institute, for the use of the NCDS data and to the UK Data Service for making them available. However, neither CLS nor the UK Data Service bear any responsibility for the analysis or interpretation of these data.

Netherlands Twin Register: The Netherlands Twin Register is supported by grant NWO 480-15-001/674: Netherlands Twin Registrer Repository: researching the interplay between genome and environment (PI DI Boomsma); Individual development: Why some children thrive, and others don’t. Gravitation program of the Dutch Ministry of Education, Culture and Science and the Dutch Research Council (NWO 0240-001-003); VCWE-2021-111 (PI C Kemner); Genotyping was made possible by grants from Genetic Association Information Network (GAIN) of the Foundation for the National Institutes of Health, Rutgers University Cell and DNA Repository (NIMH U24 MH 068457-06), the Avera Institute, Sioux Falls (USA) and the National Institutes of Health (NIH R01 HD042157-01A1, MH081802, Grand Opportunity grants 1RC2 MH089951 (PI P Sullivan) and 1RC2 MH089995 (PI J Hudziak). We gratefully acknowledge the participating families and teachers of the Netherlands Twin Register (NTR) for their valuable contributions to this research.

QSKIN Study: The QSKIN Study was supported by NHMRC Grants [APP1185416, APP1073898, APP1063061]. We thank Professor David Whiteman and the participants in the QSKIN Study for contributing genetic data to this study as population controls.

The Raine Study: The Raine Study was supported by the National Health and Medical Research Council of Australia [Grant Numbers 572613, 403981, 1059711], the Canadian Institutes of Health Research [Grant Number MOP-82893], and the Western Australian Future Health Research and Innovation Fund (Grant ID WACSOSP2023-2024, WACSOSP2025/7). The authors are grateful to the Raine Study participants and their families for their continued participation in the study, as well as the Raine Study team for study coordination and data collection. We also thank the NHMRC, the Canadian Institutes of Health Research, and the Raine Medical Research Foundation for their support. The core management of the Raine Study is funded by The University of Western Australia, Curtin University, The Kids Research Institute Australia, Women and Infants Research Foundation, Edith Cowan University, Murdoch University, The University of Notre Dame Australia, and the Western Australian Future Health Research and Innovation Fund. This work was supported by resources provided by the Pawsey Supercomputing Centre with funding from the Australian Government and Government of Western Australia.

TEDS: Twins Early Development Study (TEDS) is supported by the UK Medical Research Council (MR/V012878/1 and previously MR/M021475/1). We gratefully acknowledge the ongoing contributions of the participants in TEDS and their families.

TwinsUK: TwinsUK is funded by the Wellcome Trust, Medical Research Council, Arthritis UK, European Union Horizon, Chronic Disease Research Foundation (CDRF), Wellcome Leap Dynamic Resilience Programme (co-funded by Temasek Trust), ZOE Ltd, the National Institute for Health and Care Research (NIHR) Research Delivery Network (RDN), and Biomedical Research Centre based at Guy’s and St Thomas’ NHS Foundation Trust in partnership with King’s College London.

UK Biobank: This research has been conducted using the UK Biobank Resource under Application Number 36610.

23andMe: We would like to thank the research participants and employees of 23andMe Research Institute for making this work possible.

## Data Sharing Statement

23andMe GWAS summary statistics are available through the 23andMe website to qualified researchers under agreement with 23andMe that protects the privacy of the 23andMe participants. Interested investigators should visit the 23andMe Publication Dataset Access Program at https://research.23andme.com/dataset-access.

ISP GWAS data will be made available to qualified investigators in dbGaP. Lifelines data may be obtained from a third party and are not publicly available.

Researchers can apply to use the Lifelines data used in this study. More information about how to request Lifelines data and the conditions of use can be found on their website (https://www.lifelines-biobank.com/researchers/working-with-us).

The informed consent obtained from ALSPAC (Avon Longitudinal Study of Parents and Children) participants does not allow the data to be made available through any third party maintained public repository. Supporting data are available from ALSPAC on request under the approved proposal number, B3620. Full instructions for applying for data access can be found here: http://www.bristol.ac.uk/alspac/researchers/access/. The ALSPAC study website contains details of all available data (http://www.bristol.ac.uk/alspac/researchers/our-data/).

Data from the Norwegian Mother, Father and Child Cohort Study is managed by the Norwegian Institute of Public Health. Access requires approval from the Regional Committees for Medical and Health Research Ethics (REC), compliance with GDPR, and data owner approval. Participant consent does not allow individual-level data storage in repositories or journals. Researchers seeking access for replication must apply via www.helsedata.no.

## Notes

### Competing Interest Statement

The authors have declared no competing interest.

### Summary of Updates

Updated abstract. Correction of minor typographical errors in text.

