## Supplementary Methods and Figures for "Unravelling the genetic basis of stuttering: GWAS meta-analysis highlights link with rare speech disorders"

#### Descriptions of contributing studies and phenotype definitions

##### The Genetics of Stuttering study (GenStutt)

A total of 1,607 individuals with stuttering, or a history of stuttering, were recruited for the GenStutt study between April 2018 and January 2024 from Australia (AU), New Zealand (NZ) and the United Kingdom (UK).

Participants were sought via national media campaigns and promotion through support organizations, university departments, and stuttering clinics via mail-outs, e-mail, and social media. Participants self-enrolled through the study websites (www.geneticsofstutteringstudy.org.au for AU and NZ; www.geneticsofstutteringstudy.co.uk for UK), where they provided their consent and completed surveys covering basic demographics, information about their stuttering, and general health. Participants then provided saliva samples via postal spit-kits, to allow DNA extraction and genotyping. Individuals who were already participating in the Australian Genetics of Depression Study (AGDS) were also invited to participate in GenStutt; these individuals completed the online survey but were not sent postal spit-kits, as their genetic data were already available via the AGDS (Byrne et al. 2020).[1]

Inclusion criteria were: (1) participants aged 5 years or older (UK, NZ, and AU recruited after May 2020), or aged 7 years or older (AU April 2018 – May 2020), and (2) individuals who currently stutter or have stuttered in the past. People who stuttered were asked: Have you ever stuttered? The following description was provided for the specificity of our phenotype:

*"People who stutter have trouble getting their words out. Stuttering is when people repeat sounds over and over (e.g. C-c-can I go?); repeat words or syllables over and over (e.g. Can-can-can-I go?); make long prolonged sounds (e.g. Caaaaaaaaaaaaaan I go?); have speech 'stoppages' or 'blocks' where no sounds come out."*

Persistent stuttering was defined as individuals who reported stuttering during the past 12 months.

Speech-language pathologists conducted initial face-to-face speech assessments with 195 participants (12.1%) to confirm presence of stuttering (or recovery), and stuttering severity, on a 10-point scale. There was found to be 100% agreement between clinicians and participants when identifying presence or absence of stuttering, with 81.2% of participant-reported severity ratings being within 1 point of the speech pathologists' rating, thus validating the self-report phenotype of participants as reliable.[2] A further follow-up study of 327 participants also found high agreement between self-report and speech pathologist assessment. Of the 255 participants who reported persistent stuttering, 91% displayed overt stuttering during speech assessment.[3]

Participants were excluded if they experienced any acquired neurological disorders, such as traumatic brain injury, before the onset of their stuttering.

The study was approved by the Human Research Ethics Committee at the Royal Children's Hospital, Melbourne (approval number 37353), New Zealand Health and Disability Ethics Committee (approval number 20/CEN/63) and UCL Research Ethics Committee (approval 18643/001).

#### QSkin Sun and Health Study (QSkin)

QSkin is a cohort of 43,794 adults aged 40-69 years, randomly sampled from the population of Queensland, Australia in 2011. At baseline, participants completed comprehensive questionnaires covering basic demographics, general medical history, and factors specifically related to skin cancer risk. Genetic data are available for a subset of approximately 19,000 participants[4]. In the present study, QSkin serves as a population-based control cohort for genetic analyses, after being combined with the GenStutt cases. The QSkin Sun and Health Study was approved by QIMR Berghofer Human Research Ethics Committee (P1309, P2034), including amendments to share data with this study.

#### MPI Erasmus Genetics of Stuttering (MEGS) Study

621 individuals with stuttering, or a history of stuttering, were recruited to the Max Planck Institute Erasmus Genetics of Stuttering (MEGS) Study, between December 2019 and December 2022.[5] Participants were recruited through several channels, including national media campaigns, articles in newspapers, television broadcasts, invitation through speech therapists, and via promotion through support organizations, and social media. Participant recruitment for this study followed a similar protocol to the GenStutt study. Individuals self-enrolled or were enrolled by a parent or caretaker through the dedicated study website (https://www.mpi.nl/genetica-van-stotteren), where they provided informed consent and completed adapted versions of the GenStutt surveys. Participants then submitted saliva samples for DNA analysis using postal spit-kits. Inclusion criteria for the study were (a) participants aged 7 years or older and (b) with self-reported or parent-reported stuttering. Participants or their parents were asked whether they had stuttered in the last 12 months

to determine whether they experienced persistent stuttering. The medical ethics committee of the Erasmus Medical Center in Rotterdam approved this study (registration number: MEC-2019-0491).

In addition, MEGS includes 66 children recruited as part of the RESTART-randomized trial.[6] Children who stutter aged 3 to 6 years were included between 2007 and 2010 in 20 private practice speech clinics in The Netherlands. All children who were seen during a follow-up visit for the clinical trial were asked to provide a saliva sample. Persistent stuttering was determined based on parent, teacher and trained observer ratings. The medical ethics committee of the Erasmus Medical Center in Rotterdam approved this study (registration number: MEC-2006-349) and all parents provided informed consent for the participation of their children and themselves.

#### Dutch Individual differences in language skills cohort (IDLaS-NL)

The IDLaS-NL cohort consists of 622 individuals who were recruited to investigate individual differences in language skills at the Max Planck Institute for Psycholinguistics.[7] Participants were recruited via the participant database of the Max Planck Institute for Psycholinguistics, through social media, through articles and advertisements in local newspapers and by contacting local vocational colleges. After completion of the language tests, participants submitted saliva samples for DNA analysis using postal spit-kits. The study was approved by the Radboud ECSS/FSW (ECSW-2019-074R1) and all participants provided informed consent. In the present study, IDLaS-NL serves as a population-based control cohort for genetic analyses, after being genotyped jointly with the MEGS cases.

#### The International Stuttering Project (ISP)

The ISP comprises 1,345 clinically ascertained developmental stuttering cases.[8] All participants were recruited via: the Curtin Stuttering Treatment Clinic, Perth, Australia; the SpeechMatters Clinic and the Irish Stammering Association, Dublin, Ireland; the National Stuttering Association, USA; online recruitment on reddit.com; and Dr. Shelly Jo Kraft's research team at Wayne State University. Stuttering was confirmed in all recruited individuals by a speech pathologist with expertise in fluency disorders. Up to five ancestry- and sex-matched population-based control subjects per case individual were selected from BioVU, Vanderbilt University Medical Center's (VUMC's) electronic health record-linked biobank. Controls were screened for developmental, speech, or language disorders, via ICD9 and ICD10 codes, and using a phenome-risk classifier.[9]

ISP studies (protocol 0225119MP2E) were approved by Wayne State University's Committee for the Protection of Human Subjects

#### Early Language in Victoria Study (ELVS)

ELVS is a longitudinal study initiated in 2003, following a cohort of 1,910 children from infancy to adolescence in Victoria, Australia. It aims to investigate language development, literacy, and related outcomes.[10] When children were 2 years of age, parents were contacted about a sub-study in ELVS focused on stuttering.[11] Parents were also given a fridge magnet with examples of stuttering features. Parents were sent reminder letters every 4 months to contact the research team if they noticed their child stuttering or showing any of the behaviors on the fridge magnet. When a parent reported that their child was stuttering, a speech pathologist interviewed the parent over the phone to confirm the speech behaviors as stuttering. Where confirmation was not clear, a home visit was arranged. Home visits with a speech pathologist were arranged wherever possible, within 2 weeks of the initial telephone interview. The home visits included a 25-minute play session between parent and child where the child's speech was rated using the stuttering severity scale (10 point scale where 1 represents no stuttering, 2 extremely mild stuttering and 10 extremely severe stuttering), by both the parent and speech pathologist. Two speech pathologists then confirmed stuttering in the cases[12] using the Stuttering Severity Scale, which rates stuttering from 0 (no stuttering) to 9. In the present GWAS study, cases were individuals who had been rated 2 or above on the Stuttering Severity Scale. Controls were individuals who either did not have stuttering reported or who had been rated by the speech pathologists to have a score of 1 on the severity scale.

#### The Raine Study

The Raine Study, established in 1989, is one of the largest prospective cohorts of pregnancy, childhood, adolescence, and adulthood. It has followed nearly 3,000 participants from before birth through to adulthood, providing valuable insights into health and well-being.[13]

Parents were asked about stuttering when participants were 5, 8, and 10 years. At 5 and 8 years, ratings ("always"; "mostly"; "sometimes"; "never") were provided in response to the statement "My child stutters when talking". At 10 years, the statement was "My child stutters (e.g. repeats words unnecessarily; draws words/sounds out, or gets stuck on certain words or sounds while speaking)", with ratings: "no problem"; "minor problem"; "major problem". Participants were recorded as a case if they were reported as stuttering “always” or “mostly” at age 5 or 8, or if it was reported that stuttering was a “major problem” at age 10. Participants not meeting these definitions for stuttering were defined as controls.

#### Australian Twins (AusTwins)

Participants were recruited from twin studies conducted at the Queensland Institute of Medical Research (QIMR) using subjects enrolled on the Australian Twin Registry. Phenotyping for stuttering was performed via a one-word prompt “Stuttering” (or “Stuttering or stammering”) within a broader disease/medical checklist. Data came from two main assessments: an older cohort (Cohort 1): Initially assessed in the early 1980s (Canberra survey) and re-assessed in 1988 (White/Green Health & Lifestyle Survey); and a younger cohort (Cohort 2): Assessed in 1989 (Yellow/Green Health & Lifestyle Survey; primarily individuals aged 17–29). Participants answered “yes” (cases) or “no” (controls). Both cohorts are population-based and unselected.[14,15]

#### Netherlands Twin Register (NTR)

Founded in the mid-1980s, the NTR collects data on twins and their families throughout the lifespan, focusing on health, lifestyle, and behavior. It includes over 250,000 participants, including around 120,000 twins.[16]

Stuttering status was determined based on parental reports when the child was 5 years old. Mothers evaluated the frequency of six speech behaviors during typical conversational speech, using a 5-point scale (1 = never to 5 = very often). These behaviors included: (a) repeating part of a sentence (e.g., a phrase); (b) slowly repeating whole words in a sentence; (c) rapidly repeating whole words in a sentence; (d) repeating part of a word (e.g., a syllable); (e) experiencing blocks at the beginning or middle of a word; and (f ) prolonging a sound within a word.

Children were classified as having probable stuttering (PS) if they exhibited one or more core stuttering behaviours (items d-f) "often" or "very often". Those displaying at least two non-fluent behaviours (items a-c) "often" or "very often", while exhibiting core stuttering behaviours infrequently, were classified as highly non-fluent (HNF).[17] Children meeting PS criteria were designated as stuttering cases. Those not meeting criteria for either PS or HNF were classified as controls with typical fluency.

#### Twins Early Development Study (TEDS)

TEDS is a UK-based longitudinal study of over 12,000 twin pairs born between 1994 and 1996, investigating how genes and environments influence language, cognition and behavioral development.[18] Stuttering status was determined based on parental reports at multiple time points. Cases were defined as children reported to have a stammer or stutter at age 7, as indicated by a "yes, stammer/stutter" response to the "Talking difficulties" question in the 7-year parent questionnaire. Controls were defined as children who showed no symptoms of stuttering at ages 3, 4, and 7, based on parental reports at each of these time points.

#### Twins UK

Established in 1992, TwinsUK is the UK's largest adult twin registry, with over 14,000 twins enrolled. Detailed longitudinal data are available on participants, collected via numerous questionnaires, clinical visits and biological measures. [19]

Stuttering phenotypes were determined based on three questions: a) Have you been diagnosed with a stammering/stuttering problem at any time in your life?; b) Do you currently stammer/stutter?; c) Do you think that others perceive you have a stammering/stuttering problem now? Individuals were classified as cases if they responded "yes" to either question a or b. Controls were defined as those who answered "no" to all three questions.

TwinsUK is set up and governed as a Programme of Research (IRAS: 342169, REC reference: 24/NW/0107), with broad ethics and consent covering all future studies, sample collections and sharing of data for health research.

#### UK Biobank

UK Biobank is a large-scale biomedical database and research resource containing genetic, lifestyle, and health information from half a million UK participants, aged 40-69 at recruitment between 2006-2010. All analyses with UK Biobank data were undertaken under Application 36610.

Stuttering cases were identified using primary care records (GP clinical events) accessed via UK Biobank's "Data Portal: Record Repository" on 25/08/2023. Relevant Read codes for stuttering were ascertained by identifying codings whose descriptions string-matched "stutter" or "stammer" from UK Biobank's primary care codings file (http://biobank.ndph.ox.ac.uk/showcase/showcase/auxdata/primarycare_codings.zip).

For Read v2, relevant codes were "1B92.", "2B49.", "38QT.", "E270.", and "Eu9y5", while for Read v3, identified codes were "1B92.", "2B49.", "E270.", "Ub1S6", "Ub1S7", "Ub1S8", "Ub1SA", "Ub1SB", "Ub1SC", "Ub1SD", "Ub1SE", "Xab9Y", "XaCyi", "XM0h3", "XM0jR", "XM0jS", and "XM0jT". Individuals with any of these codes were initially identified. The descriptions of all codes for identified individuals were then inspected, and any codes related to acquired stuttering were excluded. Cases were defined as individuals with any other stuttering-related code (i.e., stuttering, but not acquired stuttering). For each identified case, 20 individuals without stuttering recorded in their primary care records were randomly selected as controls, matched for sex with the cases. This methodology aimed to identify individuals with developmental stuttering while excluding cases of acquired stuttering from UK Biobank primary care records.

#### The Norwegian Mother and Child Cohort Study (MoBa)

The Norwegian Mother, Father and Child Cohort Study (MoBa) is a population-based pregnancy cohort study conducted by the Norwegian Institute of Public Health. Participants were recruited from all over Norway from 1999-2008. The women consented to participation in 41% of the pregnancies. The cohort includes approximately 114,500 children, 95,200 mothers, and 75,200 fathers. [20,21]

The stuttering phenotype in children was based on response to the maternal questionnaire, undertaken when the child was age 5 years. If Yes was answered to the question: "Has your child been assessed for language delay or other difficulties with language/speech or communication?", and "What was the conclusion after the assessment?" was answered with "5-Stammer or stutters when talking", then the child was defined as a case. All other children whose mothers answered the maternal questionnaire at child age 5 were defined as controls.

#### Generation R

Generation R is a population-based prospective cohort study from fetal life until adulthood in Rotterdam, The Netherlands. It includes 9,778 mothers and their children, focusing on early environmental and genetic causes of growth, development, and health.[22]

At the age of 9–12 years, parents completed a speech and language developmental questionnaire. Four questions from this questionnaire were utilized to obtain a measure of childhood stuttering:

1. In the past, has your child had any (more than one answer possible): language problems (using word and sentences), speech problems (speaking clearly), stuttering (not speaking fluently), none of the problems

2. Does your child currently have any (more than one answer possible): language problems (using word and sentences), speech problems (speaking clearly), stuttering (not speaking fluently), none of the problems

3. Has your child any speech therapy in the past?:  No, Yes. If yes, in the past my child has had speech therapy for (more than one answer possible): language problems (using word and sentences), speech problems (speaking clearly), stuttering (not speaking fluently), none of the problems

4. Does your child currently have speech therapy?:  No, Yes. If yes, my child currently receiving speech therapy for (more than one answer possible): language problems (using word and sentences), speech problems (speaking clearly), stuttering (not speaking fluently), none of the problems

Participants were classified as stuttering cases if a parent answered "yes" and indicated stuttering to one of the above-mentioned questions. All other individuals were classified as controls.

#### 1958 National Child Development Study (NCDS1958)

The NCDS follows the lives of 17,000 people born in England, Scotland, and Wales in a single week of 1958.[23]

Stuttering phenotypes were defined using data from multiple time points and sources. The 'stuttering phenotype' included cases based on positive responses to stuttering questions at age 7 or 16 years, reported by parents, or teachers, or recorded when participants attended medical exams. At age 7, parental reports (N225) indicated whether stuttering had ever occurred (Yes/No/Don't know), while medical exam reports (N385) categorised stuttering severity (Severe/Moderate/Slight/No/Don't know/not tested). At age 16, medical exam reports (N1947) classified stuttering as Yes-severe/Yes-moderate/Yes-slightly/No/Don't know, teacher assessments (N2319) used a scale of Certainly applies/Applies somewhat/Does not apply, and parental reports (N2507) categorised stuttering as Yes-severe/Yes-mild/No. Controls for this phenotype were individuals with consistent negative responses across all time points and sources.

The 'Persistent stuttering' phenotype defined cases as those reporting stuttering at both age 7 (parental report) and age 16 years (any source), while controls were individuals consistently reporting no stuttering at both time points.

For both phenotypes, individuals not meeting either case or control criteria were excluded from the analysis.

#### The Avon Longitudinal Study of Parents and Children (ALSPAC)

ALSPAC is trans-generational prospective observational study investigating influences on health and development across the life course. Pregnant women resident in Avon, UK with expected delivery dates between April 1991 and December 1992 were recruited, with an initial enrolment of 14,541 pregnancies. Following additional recruitment phases, the total sample available for analyses using data collected after age 7 years comprises 15,447 pregnancies (14,901 children alive at 1 year of age).[24]

Cases and controls for the stuttering GWAS were defined using multiple criteria, using data collected via questionnaires, and speech and language assessments at age 8 years. Cases were identified as children meeting at least one of the following: (1) parent-reported stuttering or stumbling with expressed concern, (2) confirmed stuttering by expert rating, or (3) ≥3% of syllables stuttered. The parent-report criterion was based on a questionnaire asking whether the child stutters or stumbles when talking, with an additional question about parental concern. Expert rating involved a trained assessor listening to audio recordings of the child's speech and confirming the presence of stuttering. The syllable stuttering percentage was calculated from these recordings. Controls were defined as children not meeting any of the case criteria, with additional exclusions applied. Children were excluded from the control group if they had a diagnosed phonological disorder, parent-reported stuttering or stumbling without parental concern, or if they were assessed for expert rating.[25]

Ethical approval for the study was obtained from the ALSPAC Ethics and Law Committee and the Local Research Ethics Committees. Informed consent for the use of all data collected was obtained from participants following the recommendations of the ALSPAC Ethics and Law Committee at the time. Participants can contact the study team at any time to retrospectively withdraw consent for their data to be used. Study participation is voluntary and during all data collection sweeps, information was provided on the intended use of data. Please note that the ALSPAC study website contains details of all the data that are available through a fully searchable data dictionary and variable search tool (http://www.bristol.ac.uk/alspac/researchers/our-data/).

#### Lifelines

Lifelines is a multi-disciplinary, prospective, population-based cohort study examining in a unique three-generation design the health and health-related behaviors of 167,729 persons living in the North of the Netherlands. It employs a broad range of investigative procedures in assessing the biomedical, socio-demographic, behavioural, physical and psychological factors which contribute to the health and disease of the general population, with a special focus on multi-morbidity and complex genetics.[26] The general Lifelines protocol has been approved by the UMCG Medical ethical committee under number 2007/152. All participants gave informed consent for participating in Lifelines.

Stuttering cases and controls were defined based on self-report questionnaire responses. Participants were asked, "Have you ever stuttered or do you currently stutter?" with five possible responses: (1) "Yes, I stuttered as a child, but I do not stutter anymore", (2) "Yes, I started stuttering as a child and I still have a stutter", (3) "Yes, as a young person/adult I stuttered for a while, but I do not stutter anymore", (4) "Yes, I started stuttering as a young person/adult and I still stutter", and (5) "No, I have never stuttered". Two follow-up questions were posed to those who reported stuttering: "At what age did you begin to stutter?" and "At what age did the stuttering stop?", with participants asked to provide their best numerical estimates. Cases were defined as individuals who reported stuttering onset before the age of 21 years. Among these cases, those who indicated they "still have a stutter" were classified as having persistent stuttering. Controls were defined as individuals who responded "No, I have never stuttered".

#### Generation Scotland : Scottish Family Health Study (GS:SFHS)

GS:SFHS is a family- and population-based study of genetic and environmental determinants of health and disease in Scotland, involving over 24,000 participants in approximately 7,000 family groups.[27]

Stuttering was assessed using a self-report questionnaire in adulthood. Participants were asked the question, "Have you ever had a stutter/stammer?" with three possible responses: "Yes", "Not now but in the past", and "No”. Cases were defined as individuals who responded either "Yes" or "Not now but in the past". Controls were those who responded "No", reporting no history of stuttering.

### Genotyping, quality control and study-level analyses

#### GenStutt and QSkin

Genotyping was performed using the Illumina Global Screening array for both GenStutt and QSkin samples. Quality control procedures were implemented using PLINKv.1.90, PLINK2, KING v2.3.2, and the 1KG_HRC alignment tool (https://www.chg.ox.ac.uk/~wrayner/tools/HRC-1000G-check-bim-v4.3.0.zip).

Initial QC on QSkin data excluded SNPs with BLAST issues, GenTrain score < 0.6, Hardy-Weinberg equilibrium p-value < $10^{-6}$, X chromosome SNPs with > 1% heterozygosity in males, and SNPs with call rate < 0.95. Samples with call rate < 98% were also excluded.

GenStutt samples were genotyped in multiple batches (AUNZ batches 1-3, UK batch, and AGDS-recruited samples). A number of QSkin samples were included within each GenStutt batch for quality control purposes. Within each batch, SNPs and samples with call rates < 98% were excluded. AGDS-recruited GenStutt samples underwent additional QC mirroring the QSkin criteria.

Following batch-specific QC, all batches were merged, retaining SNPs present in > 90% GenStutt cases and > 95% of individuals overall. Further QC steps included exclusion of samples with indeterminable or discordant sex and X chromosome SNPs with > 5% heterozygous calls in males. Consistency across batches for the QSkin samples included in each GenStutt batch was examined, with any SNP not showing 100% concordance being excluded.

Ancestry principal component analysis was performed using KING. Individuals who were genetically similar to the 1000 Genomes European reference population were retained. Batch effects were examined among GenStutt samples, with SNPs showing association with batch ($P < 10^{-5}$, either PC-adjusted or unadjusted) being excluded.

Finally, A/T & G/C SNPs with MAF > 0.4, SNPs with differing alleles, SNPs with > 0.2 allele frequency difference from the Haplotype Reference Consortium (HRC) reference panel, and SNPs not in the reference panel were removed.

Genotype imputation was performed using the Michigan Imputation Server. Phasing was conducted using Eagle v2, followed by imputation with Minimac v4. The HRC r1.1 panel was used as the reference for the European population.

Association analyses were carried out using SAIGE (docker, version 1.3.0), with adjustment for sex and the first ten ancestry PCs.

#### MEGS and IDLaS-NL

Genotyping was performed using the Illumina Infinium Omni Exome2.5 array. Quality control procedures were implemented using PLINK 1.9b6, eigensoft 7.21 and the 1KG_HRC alignment tool (https://www.chg.ox.ac.uk/~wrayner/tools/HRC-1000G-check-bim-v4.2.11.zip).

SNP-based QC excluded variants with call rates < 98%, those deviating from Hardy-Weinberg equilibrium ($p < 1 \times 10^{-6}$), and SNPs with MAF < 0.01. Additionally, SNPs with MAF differing by more than 0.2 from the HRC reference were removed.

Sample-based QC excluded samples with call rates < 98%. Sex mismatches were identified by comparing reported sex with genetic sex based on X and Y-chromosomal variants. Ancestry was estimated using PCA, with 1000 Genomes data, excluding individuals whose PC1 and/or PC2 values fell outside the range of European subsets. Heterozygosity outliers beyond ±4 standard deviations were removed.

Genotype imputation was performed using a local copy of the Michigan Imputation Server. Phasing was conducted using eagle-2.4, followed by imputation with minimac4-1.0.2. The 1000 Genomes Phase 3 v5 panel was used as the reference for the European population.

Association analyses were carried out using rvtests (version 2.1.0), with adjustment for sex and the first four ancestry PCs.

#### ISP

Genotyping was performed using the Illumina Expanded Multi-Ethnic Genotyping Array. Quality control procedures were implemented using PLINKv.1.90.

SNP-based QC excluded variants with call rates < 98% for controls and < 97% for cases, those deviating from Hardy-Weinberg equilibrium ($p < 1 \times 10^{-15}$), genotyped duplicate variants, indels, and variants with MAF < 0.001.

Sample-based QC excluded samples with call rates < 97% for controls and < 95% for cases. Sex mismatches were identified by removing samples with ambiguous sex, defined as minimum F on X chromosome < 0.8 for males or maximum F on X chromosome > 0.4 for females. Samples were also excluded if their expected and genetic sex did not match without a determinable cause. Ancestry was assigned using PCA with HapMap3 reference data, calculating likelihoods for each ancestry group. Heterozygosity outliers with F > 0.2 were removed. Additional exclusions were made for variants with MAF < 0.01 and $r^2 < 0.4$.

Genotype imputation was performed using the TopMed Server. Phasing was conducted using EAGLE v.2.4, followed by imputation with Minimac4. The TopMed panel was used as the reference for the European population.

Association analyses were carried out using SAIGE (docker, version 0.43.2), with adjustment for sex and the first six ancestry PCs.

#### ELVS

Genotyping was performed using the Illumina Global Screening array. Quality control procedures were implemented using PLINKv.1.90, PLINK2, and the 1KG_HRC alignment tool (https://www.chg.ox.ac.uk/~wrayner/tools/HRC-1000G-check-bim-v4.2.11.zip).

SNP-based QC excluded variants with call rates < 98% and those deviating from Hardy-Weinberg equilibrium ($p < 1 \times 10^{-10}$).

Sample-based QC excluded samples with call rates < 98%. Individuals with discrepant or ambiguous sex data were removed. Ancestry principal component analysis was performed using PLINK2. Individuals who were genetically similar to the 1000 Genomes European reference population were retained. Heterozygosity outliers beyond ±4 standard deviations from the mean were excluded.

Finally, A/T & G/C SNPs with MAF > 0.4, SNPs with differing alleles, SNPs with > 0.2 allele frequency difference from the Haplotype Reference Consortium (HRC) reference panel, and SNPs not in the reference panel were removed.

Genotype imputation was performed using the Michigan Imputation Server. Phasing was conducted using Eagle v2, followed by imputation with Minimac v4. The HRC r1.1 panel was used as the reference for the European population. Post-imputation QC applied tiered filtering criteria based on minor allele frequency (MAF) and imputation quality (Rsq). Genotyped SNPs were retained if MAF > 5e-4. Imputed SNPs were filtered using Rsq ≥ 0.5 for MAF ≥ 0.01 and Rsq ≥ 0.8 for MAF < 0.01.

Association analyses were carried out using SAIGE (docker, version 1.3.0), with adjustment for sex and the first ten ancestry PCs.

#### The Raine Study

Genotyping was performed using the Illumina Human660W-Quad BeadChip. Quality control (QC) procedures were implemented using PLINK v1.9.

SNP-based QC excluded variants with call rates < 0.95, those deviating from Hardy-Weinberg equilibrium ($p < 1 \times 10^{-6}$), SNPs with MAF < 0.01, and AT/CG ambiguous SNPs.

Sample-based QC involved a two-stage call rate filtering: exclusion if call rate < 0.95 for SNPs with MAF ≥ 0.05, or < 0.99 for SNPs with MAF < 0.05. Sex mismatches were identified by cross-checking genotyped data against core data. Participants were excluded if both parents were not of European ancestry. Heterozygosity outliers (> 0.32) were removed based on histogram inspection. Samples showing cryptic relatedness (identity-by-descent proportion > 0.1875) were excluded, retaining the individual with lower missing genotype rate in related pairs.

Genotype imputation was performed using the Michigan Imputation Server. Phasing was conducted using SHAPEIT, followed by imputation with Minimac v3. The HRC r1.1 2016 panel was used as the reference for the European population.

Association analyses were carried out using rvtests (version 20150104), with adjustment for sex and the first two ancestry PCs.

#### AusTwins

Genotyping was performed using multiple Illumina family arrays (see [28] for full description of genotyping and QC). After QC, genotyping data were imputed to the Haplotype Reference Consortium (HRC) reference panel v1.1. Association analyses were carried out using RareMetalWorker (version 4.13.7) with adjustment for sex, imputation run, and the first four ancestry PCs.

#### NTR

Genotyping was performed using three platforms: Affymetrix 6.0 (n=579 samples), Affymetrix Axiom (n=184), and Illumina GSA (n=689). Quality control procedures were implemented using Plink 1.07, Plink 1.96, and the 1KG_HRC alignment tool (https://www.chg.ox.ac.uk/~wrayner/tools/HRC-1000G-check-bim-v4.2.10.zip).

SNP-based QC excluded variants with call rates < 95%, those deviating from Hardy-Weinberg equilibrium ($p < 1 \times 10^{-6}$), SNPs with MAF < 0.0001, minor allele count (MAC) < 1, and allele frequency differences > 0.20 compared to the 1000 Genomes reference. Palindromic SNPs (A/T and C/G) with MAF 0.40-0.50 were also removed.

Sample-based QC excluded samples with call rates < 90%. Sex mismatches were identified by removing males homozygous for X-chromosome, females heterozygous for X-chromosome, and ambiguous samples. Non-CEU outliers were removed by PCA projection of 1000 Genomes on the NTR sample. Heterozygosity outliers with Plink $F_{het} > 0.10$ or $< -0.10$ were excluded. Additional exclusions were made for SNPs discordant between platforms and IBD inconsistencies in family data.

Genotype imputation was performed using the Michigan Imputation Server pipeline. Phasing was conducted using Eagle 2, followed by imputation with Minimac3. The 1000 Genomes phase 3 v5 panel was used as the reference for all ancestries. Post-imputation, VCFs across platforms were merged using Bcftools 1.9 and converted to BGEN format using Qctool 2.06.

Association analyses were carried out using SAIGE (version 0.44.5), with adjustment for genotyping platform, sex and the first ten ancestry PCs.

#### TEDS

Genotyping was performed using two platforms: AffymetrixGeneChip 6.0 (Affy, n=1,359 samples) and Illumina HumanOmniExpressExome-8v1.2 (OEE, n=2,555). Quality control procedures were implemented using PLINK, R, BCFtools, and EIGENSOFT.

SNP-based QC excluded variants with call rates < 75%, those deviating from Hardy-Weinberg equilibrium ($p < 1 \times 10^{-5}$), SNPs with MAF < 0.05, and data missingness > 2%. SNPs were also removed based on their association with the batch, plate, and/or well on which they were genotyped ($p < 1 \times 10^{-4}$).

Sample-based QC excluded samples with call rates < 98%. Sex mismatches were identified using PLINK's --check-sex flag, with all mismatches removed from the dataset. Ancestry outliers were identified by matching TEDS ancestry against 1000 Genomes Project data. Heterozygosity outliers beyond 3 standard deviations from the mean were excluded.

Genotype imputation was performed using the Sanger Imputation Service. Phasing was conducted using Eagle v2, followed by imputation using the PBWT (Positional Burrows-Wheeler Transform) algorithm. The HRC r1.1 panel was used as the reference for the European population.

Analyses were carried out separately for the two platforms. Association analyses were conducted using rvtests (version 2.1.0), with adjustment for sex, age and the first ten ancestry PCs. Genotyping batch was also included in the analyses of the OmniExpressExome samples (the Affymetrix samples were genotyped in a single batch).

#### Twins UK

Genotyping was performed using a combination of Illumina arrays (HumanHap300, HumanHap610Q, 1M-Duo and 1.2MDuo 1M). Quality control procedures were carried out for theHumanHap610Q, 1M-Duo and 1.2MDuo 1M samples combined, and for HumanHap300 separately. Quality control procedures were implemented using PLINK2, and the 1KG_HRC alignment tool (https://www.chg.ox.ac.uk/~wrayner/tools/HRC-1000G-check-bim-v4.2.5.zip).

SNP-based QC excluded variants with Hardy-Weinberg equilibrium $p < 10^{-6}$ and MAF < 1%. SNPs with call rates < 97% (for MAF ≥ 5%) or < 99% (for 1% ≤ MAF < 5%) were also removed.

Sample-based QC excluded samples with call rates < 98%. Heterozygosity outliers beyond ±2 standard deviations from the mean were excluded. Ancestry principal component analysis was performed, and individuals of non-European ancestry were removed based on comparison with HapMap3 populations. Samples with pairwise identity-by-descent probabilities suggestive of sample identity errors were also excluded.

Finally, A/T & G/C SNPs with MAF > 0.4, SNPs with differing alleles, SNPs with > 0.2 allele frequency difference to the HRC reference panel, and SNPs not in the reference panel were removed.

Genotype imputation was performed using the Michigan Imputation Server. Phasing was conducted using Eagle v2.3, followed by imputation with Minimac v4. The HRC r1.1 panel was used as the reference for the European population.

Association analyses were carried out using SAIGE (docker, version 1.2.0), with adjustment for sex and the first ten ancestry PCs.

#### UK Biobank

The genotyping procedure, quality control and imputation of the UK Biobank cohort is described in detail elsewhere [29]. Individuals with excess missingness, or outlying heterozygosity were excluded prior to imputation to a HRC and UK10K reference panel.

We further excluded individuals who had withdrawn consent, samples where the self-reported sex did not match the genetically inferred sex, samples with putative sex chromosome aneuploidy, and samples with an excess of relatives (>10 3rd degree). Samples were then restricted to the “White British” subset, as defined by UK Biobank[29]. Variants were filtered to include those with MAF>0.01%, and INFO>0.8.

Association analyses were carried out using SAIGE (docker, version 1.2.0), with adjustment for sex and the first ten ancestry PCs.

#### MoBa

Genotyping, quality control (QC), phasing, imputation, and post-imputation QC have previously been described in full.[30]

Association analyses were carried out using REGENIE v1.0.6.7, with adjustment for genotyping batch, imputation batch, sex registered at birth, and the first five ancestry PCs.

MoBa is regulated by the Norwegian Health Registry Act. The current study was approved by The Regional Committees for Medical and Health Research Ethics (2016/1702).

#### Generation R

Association analyses were conducted using rvtests (version 20190205), with adjustment for sex, and the first ten ancestry PCs.

#### NCDS1958

Genotyping was performed using multiple platforms: Illumina Human 660-Quad (n=840 samples), Illumina 1.2M (n= 2774), Infinium Human Hap 550K v1.1 (n= 64), Infinium HumanHap 550K v3 (n= 2553), and Affymetrix v6 (n= 179). Quality control procedures were implemented using Plink v1.0b4, R v4.0.3, RStudio v4.1.2, the 1KG_HRC alignment tool (https://www.chg.ox.ac.uk/~wrayner/tools/), and Perl v.5.24.0.

SNP-based QC excluded variants with call rates < 97% and those deviating from Hardy-Weinberg equilibrium ($p < 1 \times 10^{-6}$).

Sample-based QC excluded samples with call rates < 98%. Individuals with discrepant or ambiguous sex data were removed. Ancestry was determined using PCA in Plink with 1000 Genomes reference data, and individuals with non-European ancestries were removed. Heterozygosity outliers beyond ±3 standard deviations from the mean were excluded. Where samples were repeated across arrays, data from the most dense array were retained to maximise coverage. Related samples were identified using a relatedness threshold of 0.1875, with the sample having the most missing data removed from each related pair.

Genotype imputation as described in Bridges et al., (2023)[31] was performed using the Michigan Imputation Server. Phasing was conducted using Eagle v2, followed by imputation with Minimac v4. The HRC r1.1 panel was used as the reference for the European population.

Association analyses were conducted using rvtests (version 20170418), with adjustment for sex, genotyping array, and the first five ancestry PCs.

#### ALSPAC

Genotyping was performed using the Illumina Human Hap550 array. Quality control procedures were implemented using PLINK 1.9 and Eigenstrat 7.2.1.

SNP-based QC excluded variants with call rates < 95%, those deviating from Hardy-Weinberg equilibrium ($p < 1 \times 10^{-6}$), SNPs with MAF < 0.01, and those with > 0.2 difference in MAF compared to the HRC r1.1 reference panel.

Sample-based QC excluded samples with call rates < 97%. Sex mismatches were identified using PLINK's --check-sex option. Ancestry was determined using PCA, after combining ALSPAC genotype data with 1000 Genomes phase 1 data. ALSPAC individuals with PC1 and PC2 values outside the ranges of European 1000G participants were excluded. Heterozygosity outliers beyond 4 standard deviations from the mean were removed.

Genotype imputation was performed using the Michigan Imputation Server. Phasing was conducted using Eagle v2.4, followed by imputation with Minimac4. The HRC r1.1 2016 panel was used as the reference for the European population.

Association analyses were conducted using rvtests (version 20190205), with adjustment for sex, and the first ten ancestry PCs.

#### Lifelines

Genotyping was performed in three batches using three arrays: batch GWAS using the Illumina HumanCytoSNP (n~30,000 samples),[32] batch UGLI1 using the Infinium Global Screening Array (n~38,000) and batch UGLI2 using the FinnGen Thermo Fisher Axiom® custom array (n~28,000).

SNP-based QC for the CytoSNP array excluded variants with call rates < 95%, those deviating from Hardy-Weinberg equilibrium ($p < 0.001$), and SNPs with MAF < 0.01. For the

UGLI1 and UGLI2 batches, variants were excluded if call rates were < 99%, they deviated from Hardy-Weinberg equilibrium ($p < 1 \times 10^{-6}$), had MAF = 0, or showed Mendelian errors.

Sample-based QC excluded samples with call rates < 95% for GWAS and < 99% for UGLI1 and UGLI2. Sex mismatches were identified and removed. Non-European samples were removed based on PCA using 1000 Genomes and Genome of the Netherlands (GoNL) reference data, excluding those with PC1 or PC2 > 2 standard deviations from the mean. Heterozygosity outliers beyond 4 standard deviations from the mean were removed. For the GSA, duplicates and family errors were also excluded.

Genotype imputation was performed using the Sanger Imputation Service. Phasing was conducted using Eagle v2, followed by imputation using the PBWT (Positional Burrows-Wheeler Transform) algorithm (version 3.1). The HRC r1.1 panel was used as the reference for the European population. Details on the genotyping data and imputation can be found at the GWAS and UGLI wiki pages (https://wiki.lifelines.nl/doku.php?id=gwas and https://wiki.lifelines.nl/doku.php?id=ugli)

Association analyses were conducted using rvtests (version 20190205), with adjustment for sex, genotyping array, and the first ten ancestry PCs.

#### GS:SFHS

Genotyping was performed using the Illumina Human OmniExpressExome-8v1.0 BeadChip.

SNP-based QC excluded variants with ≥ 3% missing data, those deviating from Hardy-Weinberg equilibrium ($p \leq 1 \times 10^{-6}$), SNPs with MAF ≤ 1%, and those with minor allele count (MAC) < 50 in the full cohort of > 20,000 samples.

Sample-based QC excluded samples with call rates ≤ 98%. Individuals with discrepant or ambiguous sex data were removed. Ancestry outliers were identified via PCA, with those beyond 6 standard deviations from the mean of the first PC in the full Generation Scotland cohort of > 20,000 samples being removed.

Genotype imputation was performed using the Sanger Imputation Server. Phasing was conducted using Shapeit2 v2r837 with the duohmm option, followed by imputation using the PBWT (Positional Burrows-Wheeler Transform) algorithm. The HRC r1.1 panel was used as the reference for the European population. Chromosome X was imputed separately using the Michigan Imputation Server.

Post-imputation QC excluded variants with an $R^2$ score < 0.8 and monoallelic sites from the wider cohort of 20,000 samples.

Association analyses were conducted using rvtests (version 20170418), with adjustment for sex, kinship, and the first five ancestry PCs.

### Supplementary Figures

**Figure S1: Region plots and forest plots for 24 loci (A-X) meeting $p < 1 \times 10^{-5}$ in the primary GWAS of stuttering.** For each locus, the regional association plot (left) displays $-\log_{10}(p)$ for all variants within the region, with the lead SNP highlighted in purple. Variants are colored according to linkage disequilibrium ($r^2$) with the lead SNP, based on a European population. The forest plot (right) displays study-specific effect estimates (beta) and 95% confidence intervals for the lead SNP.

A. rs34204447 *(CAMTA1)*

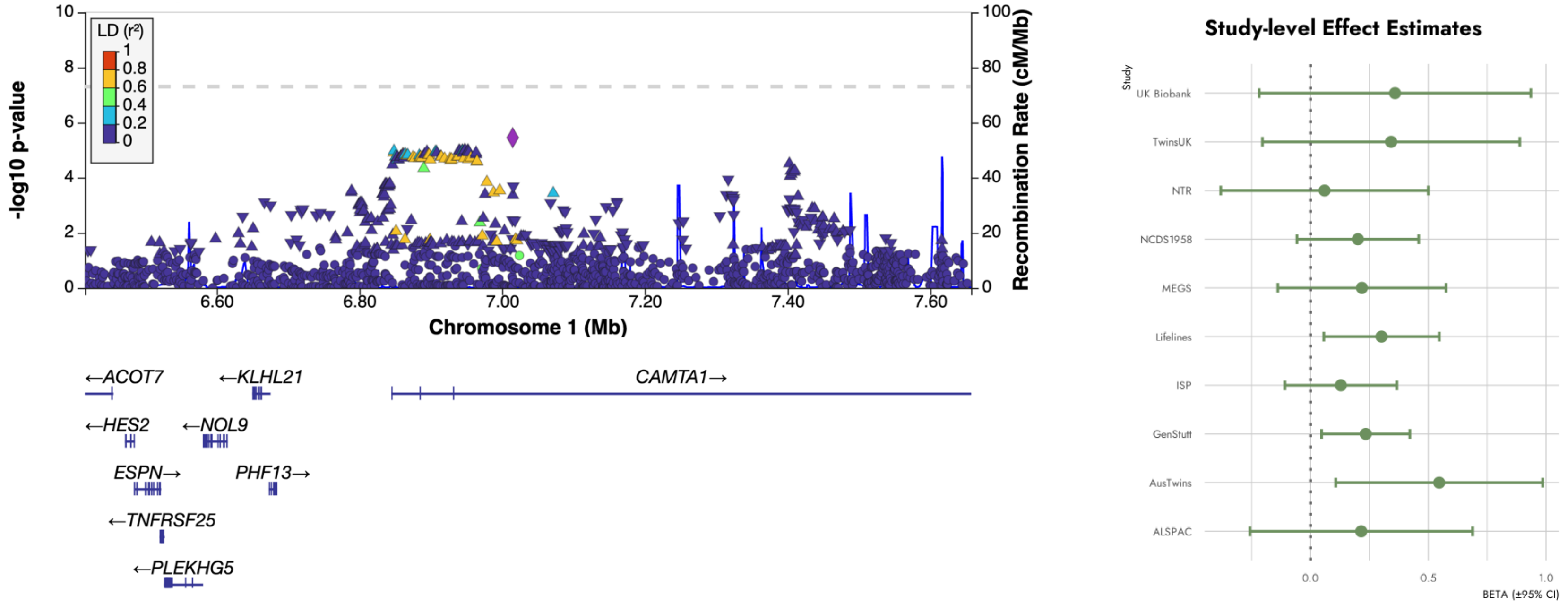

B. rs1359554 *(PTBP2)*

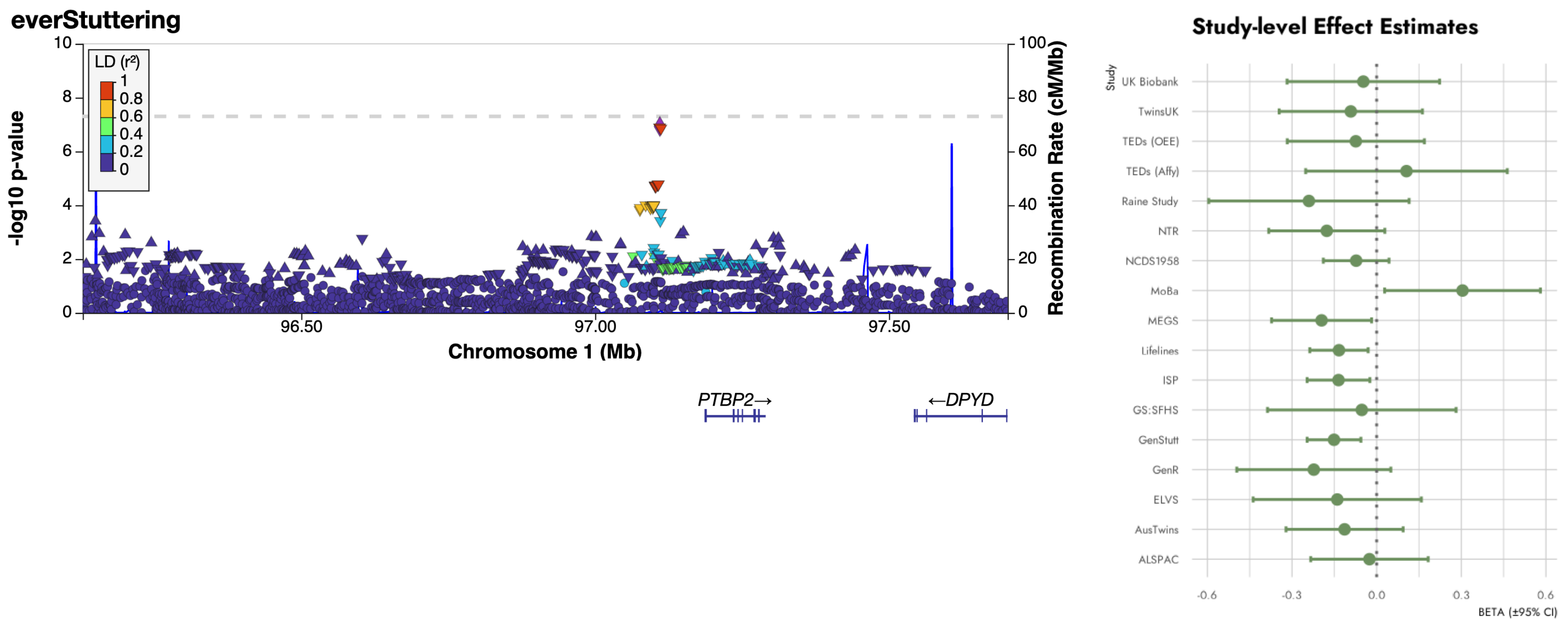

C.  rs3005776 (*KCND3*)

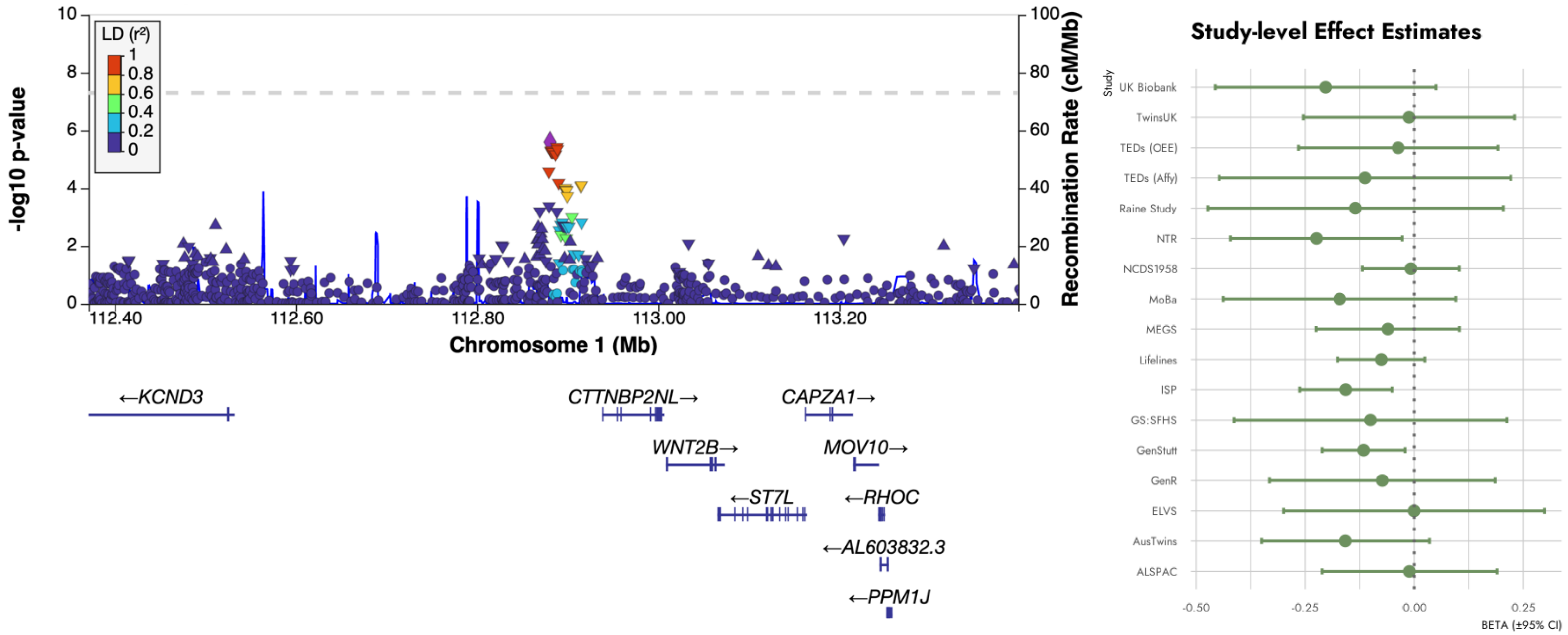

D. rs7582221 (*SUCLG1*)

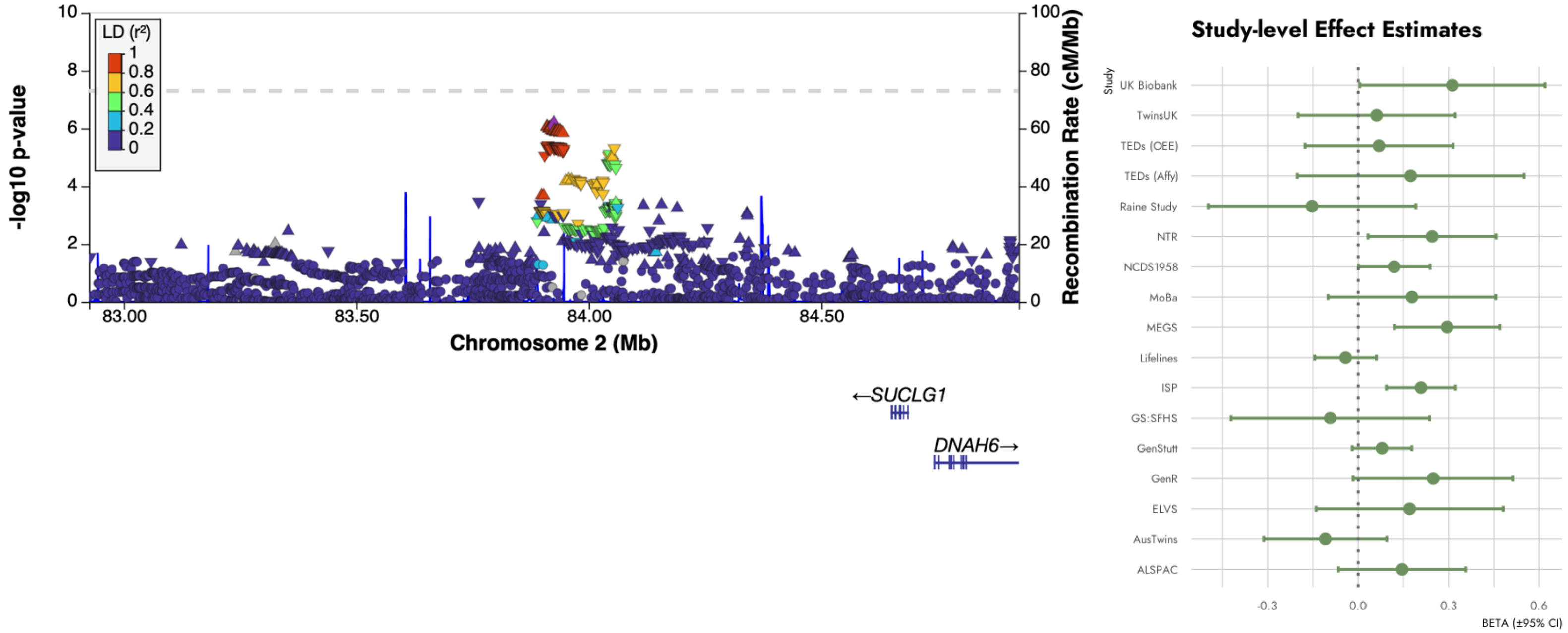

E. rs147710719 (*UBE2E3*)

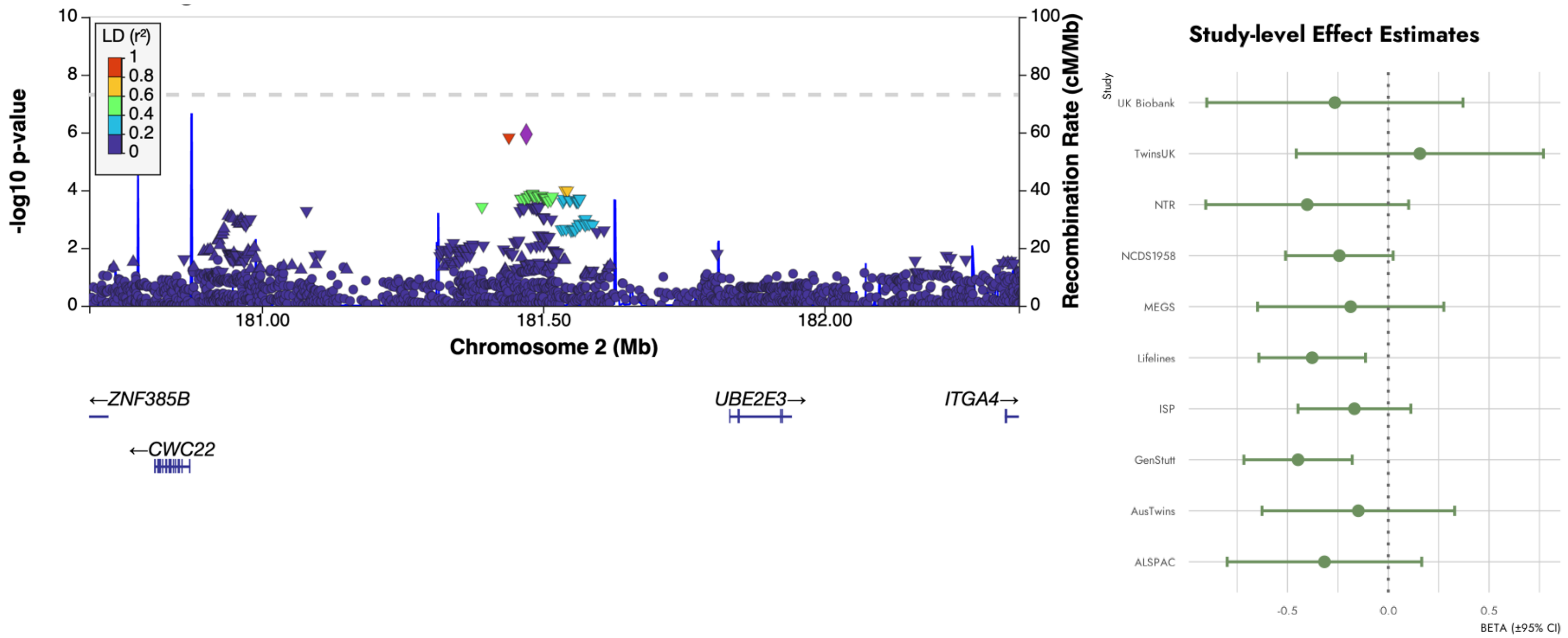

F. rs1806672 (*MIER3*)

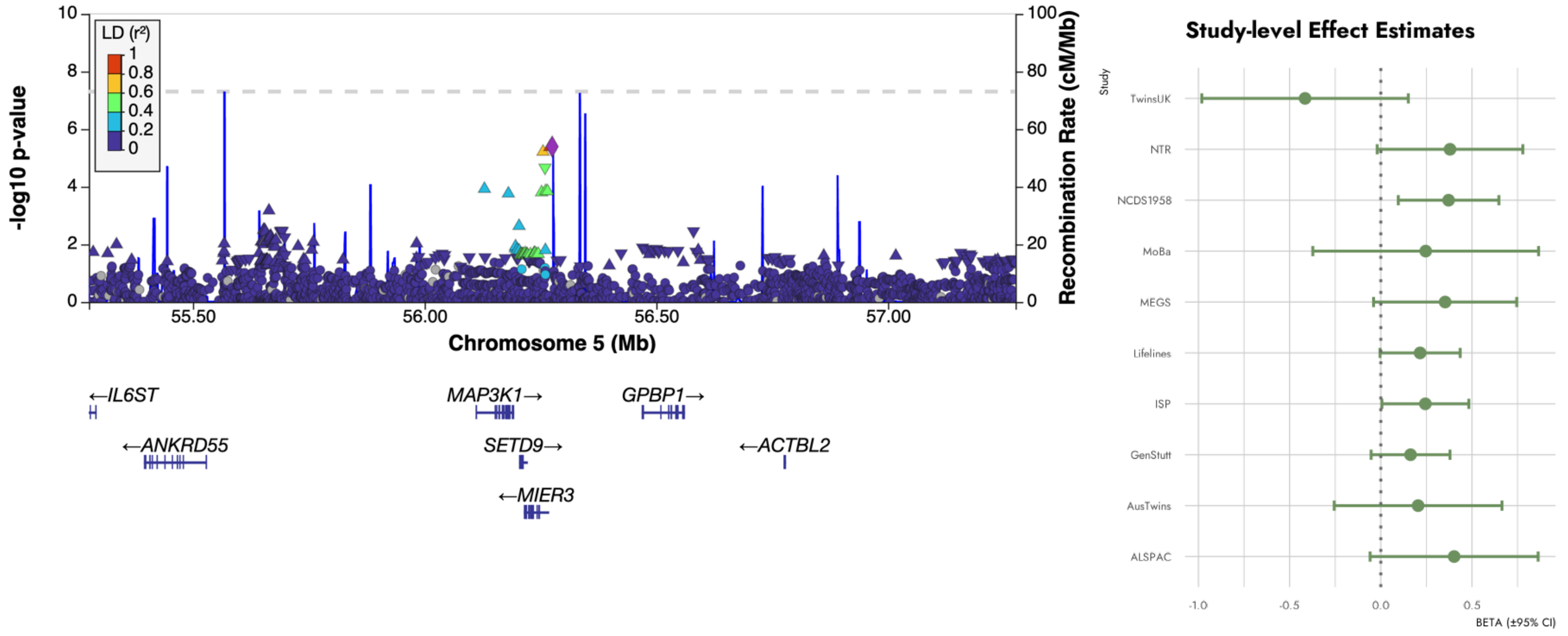

G. rs1793893 (*HCG27*)

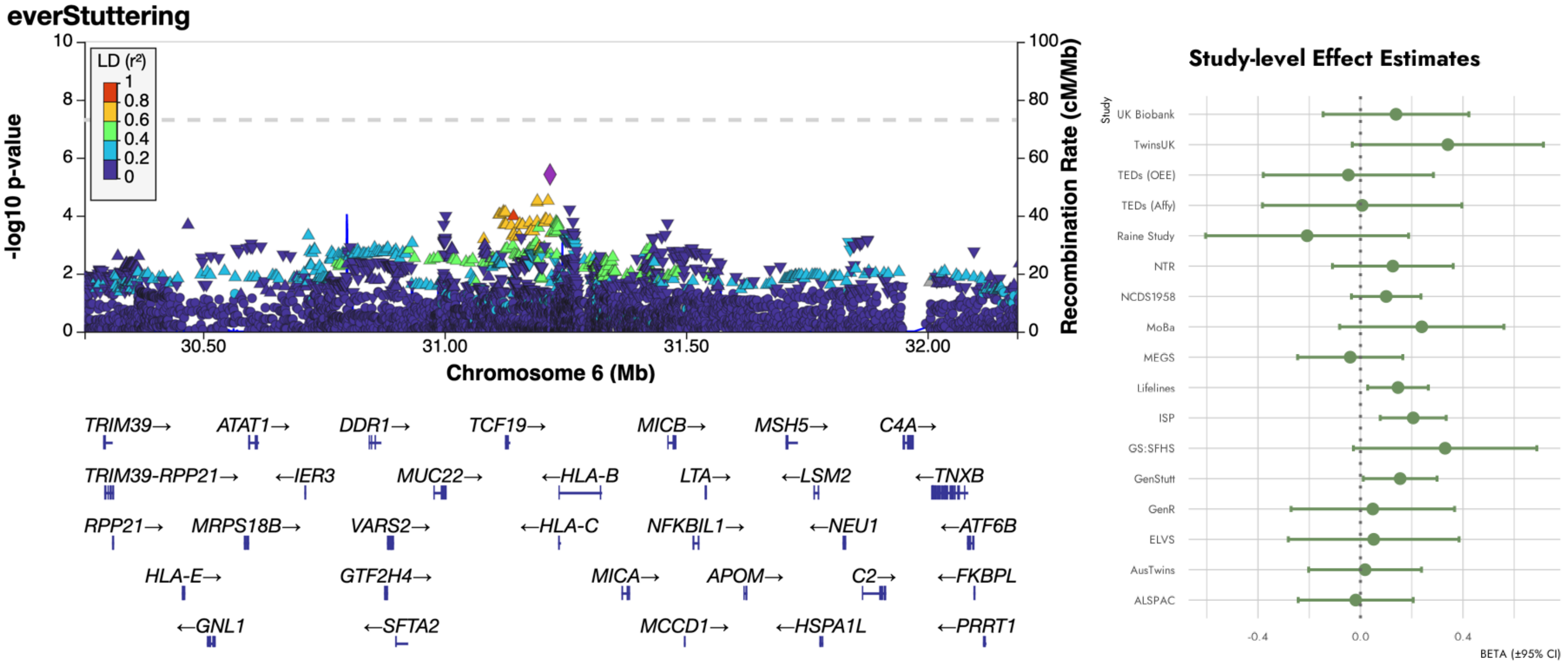

H. rs16884857 (*CDKN1A*)

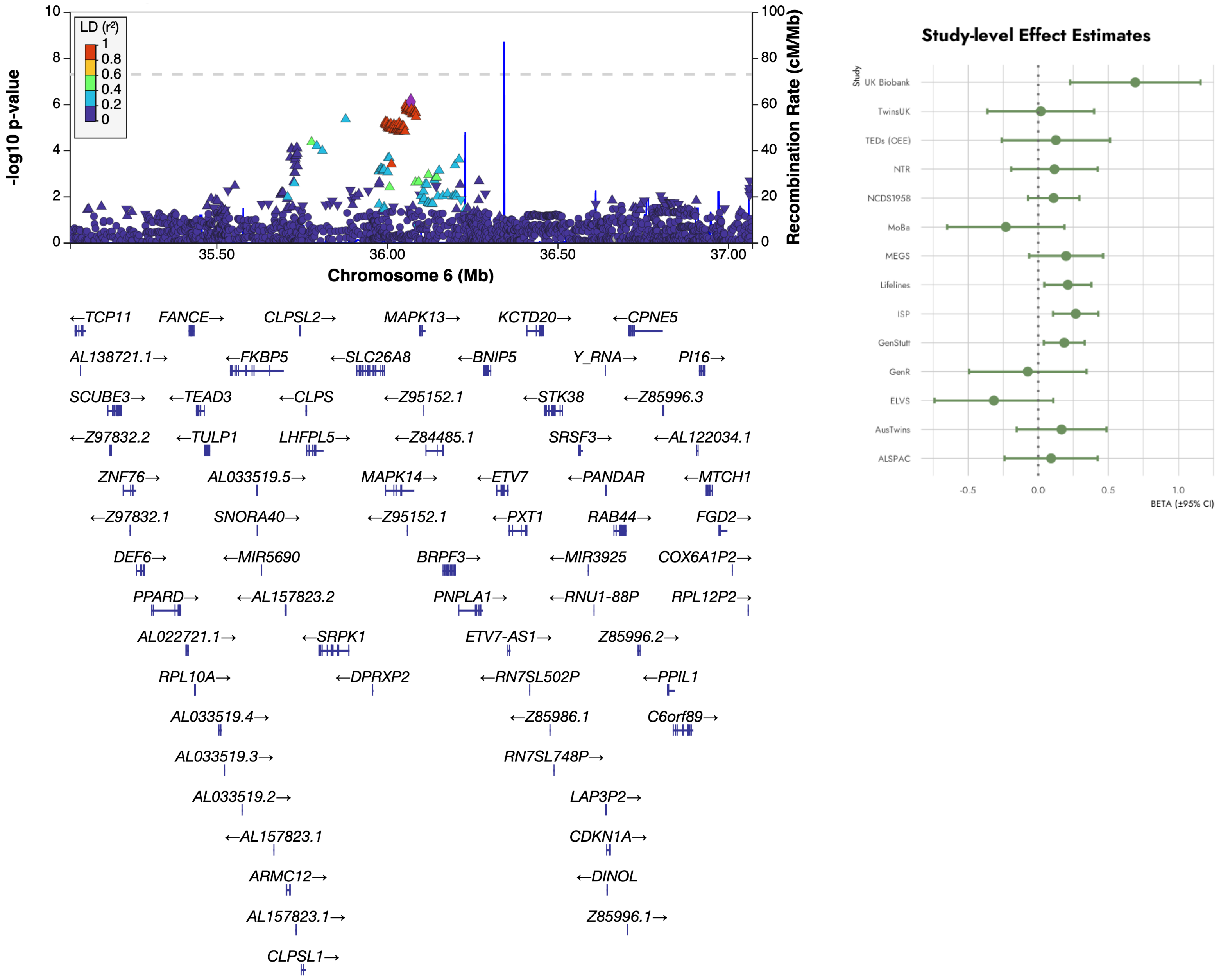

I. rs139878267 (*SLC25A51P1)*

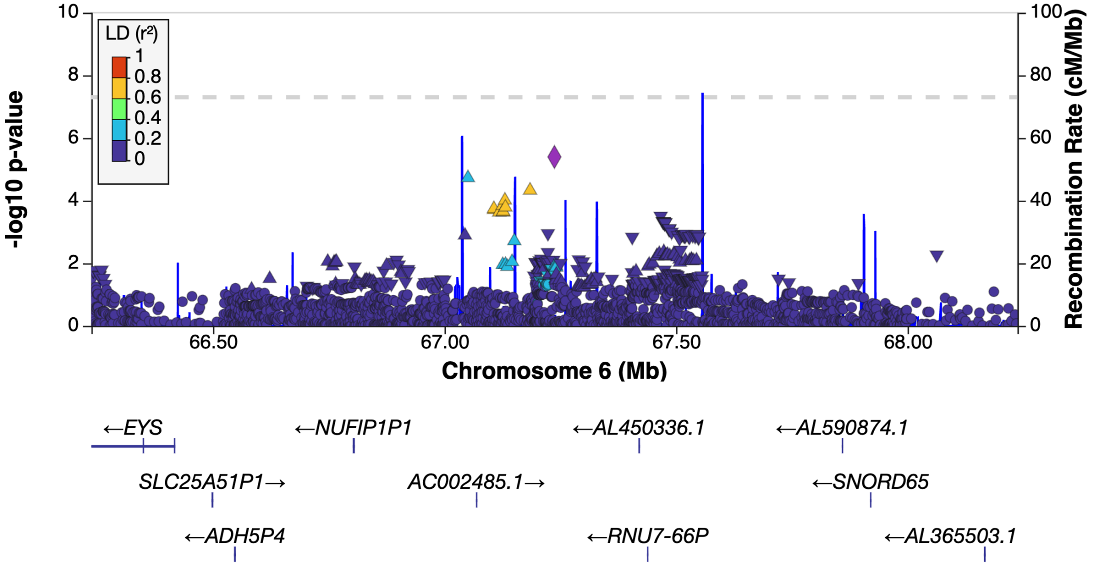


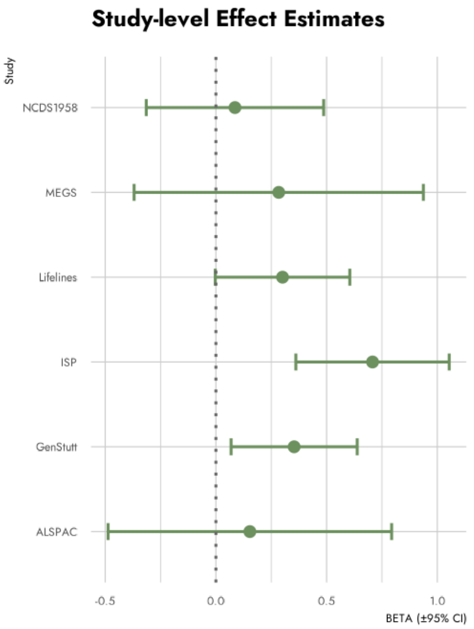

J. rs12555602 (*CAAP1*)

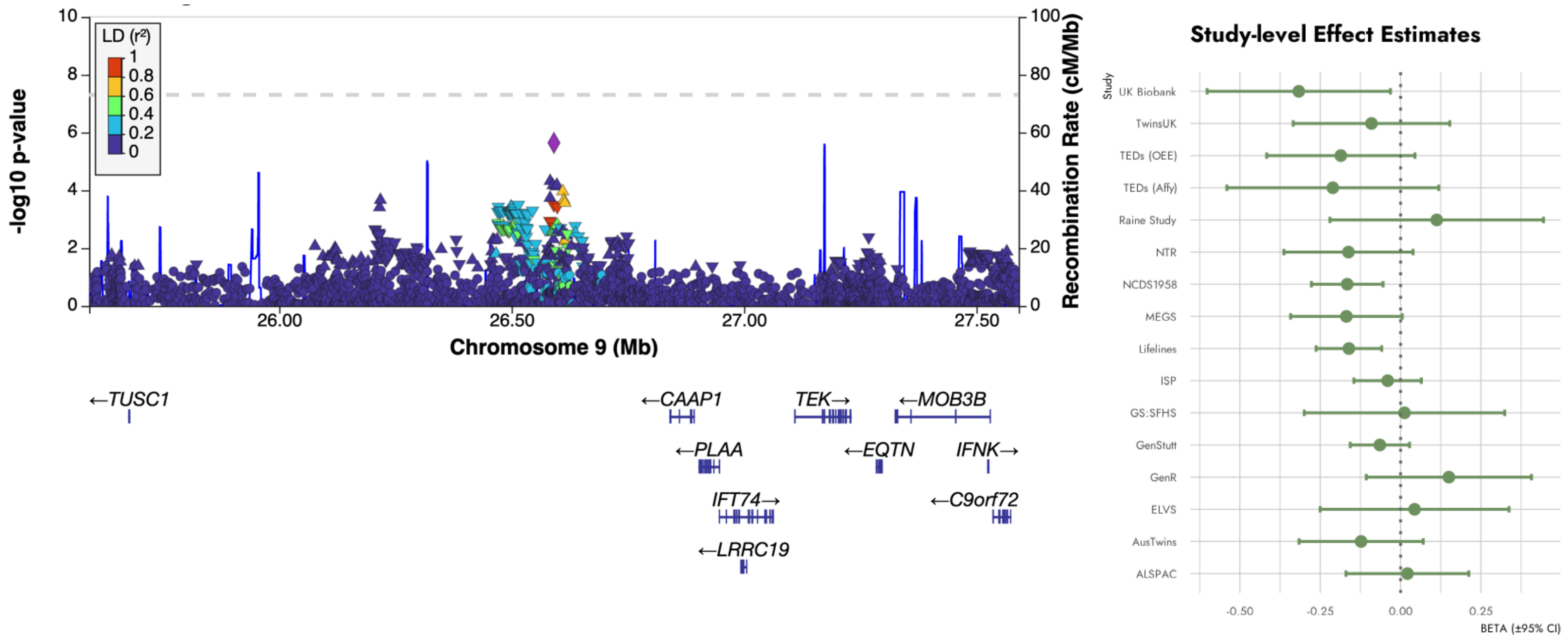

K. rs78493328 (*RORB*)

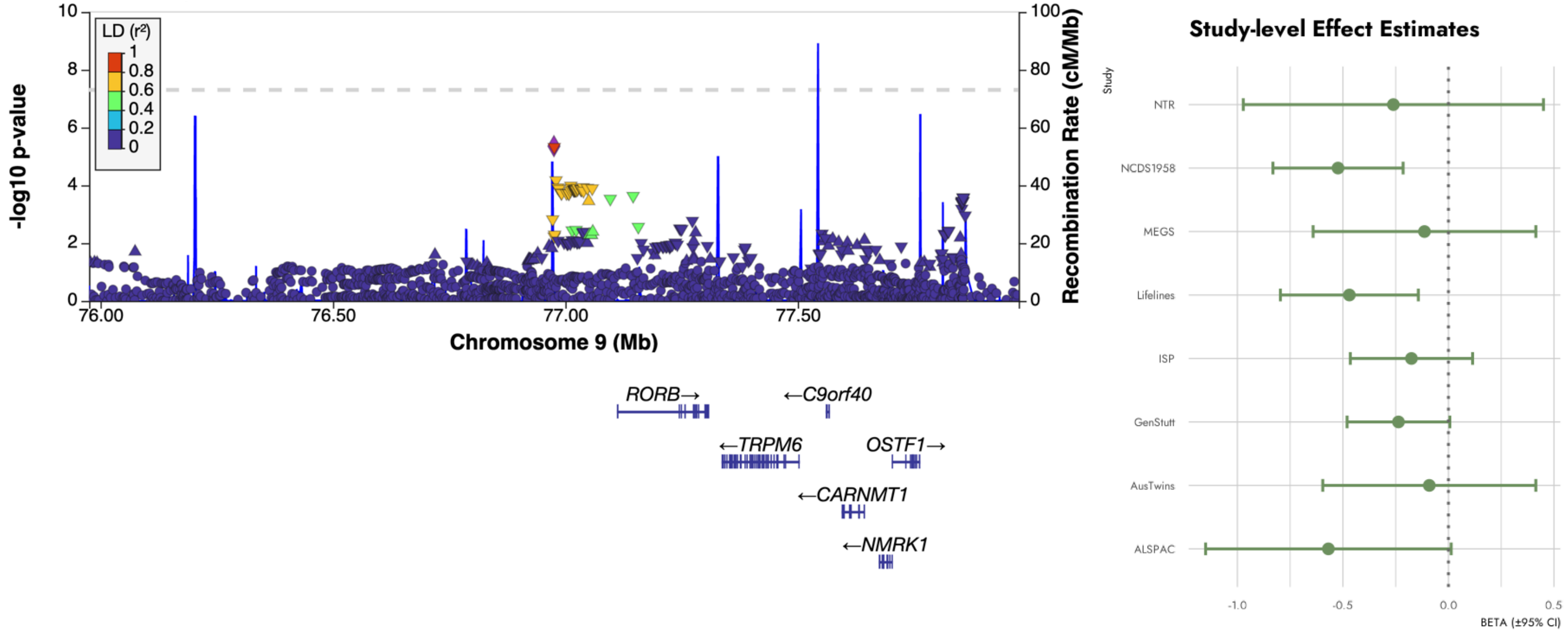

L. rs34330747 (*NELL1*)

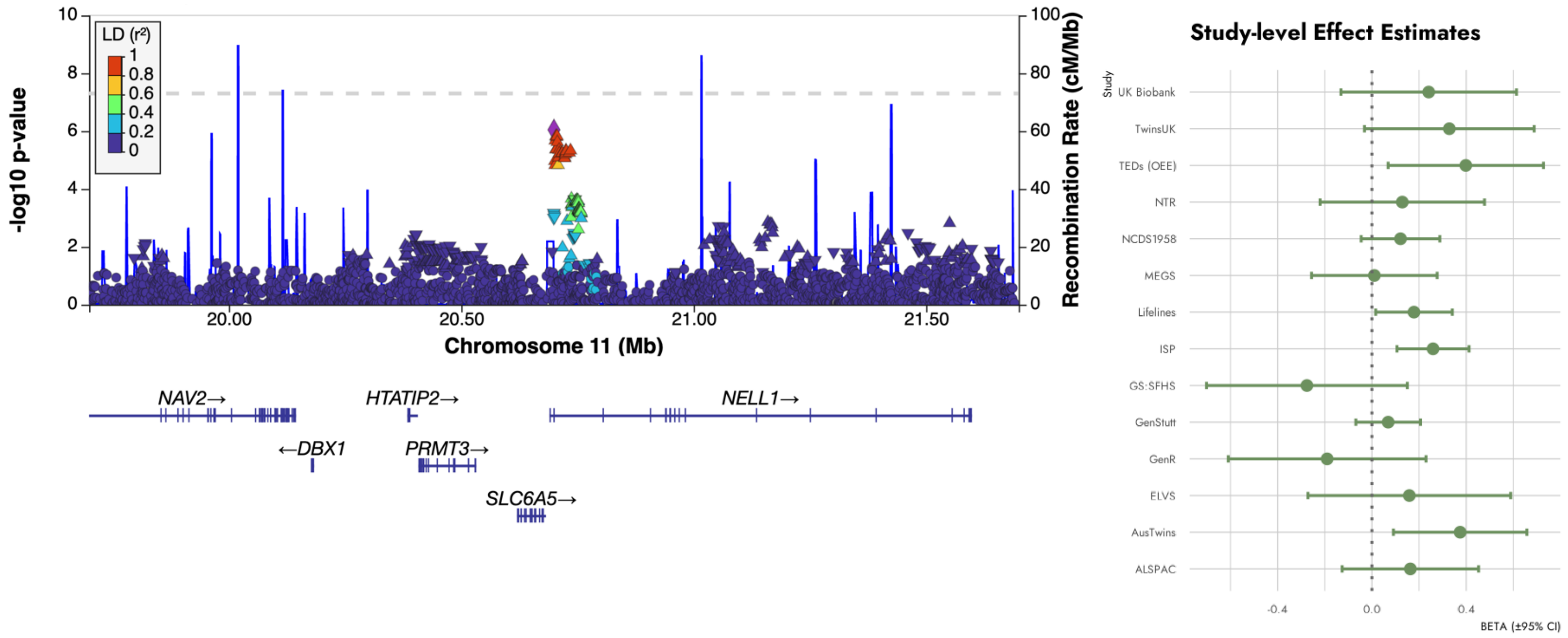

M. rs532395 (*MPPED2*)

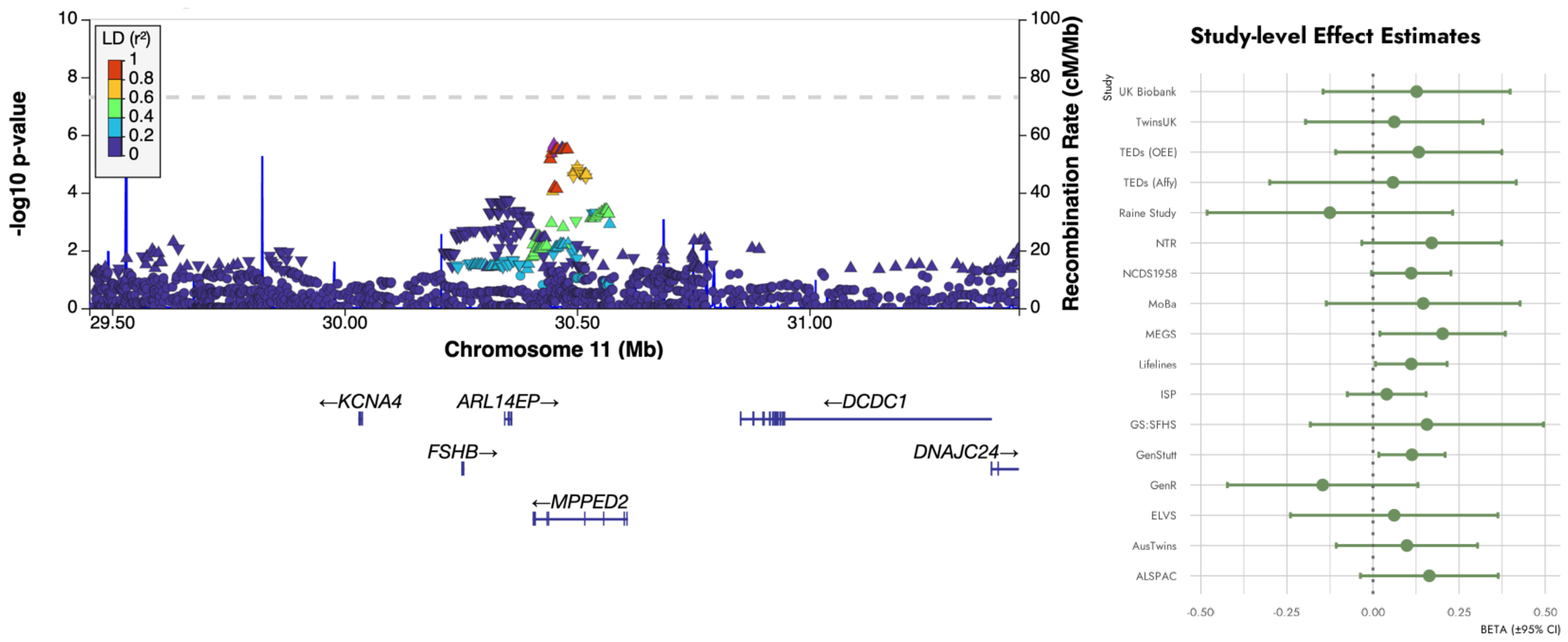

N. rs676846 (*KIRREL3*)

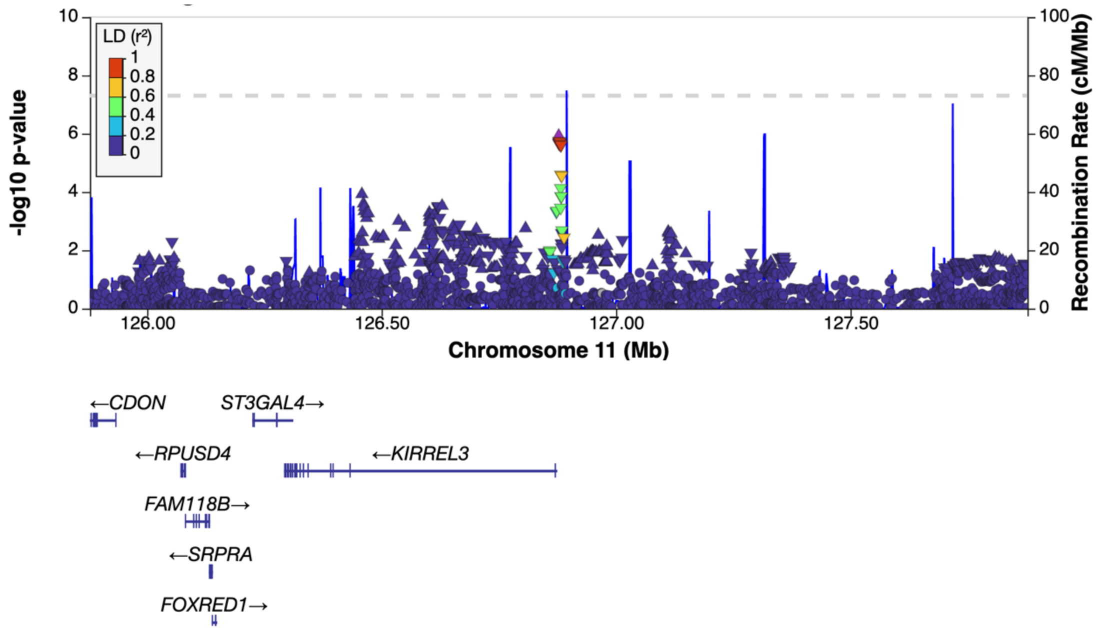


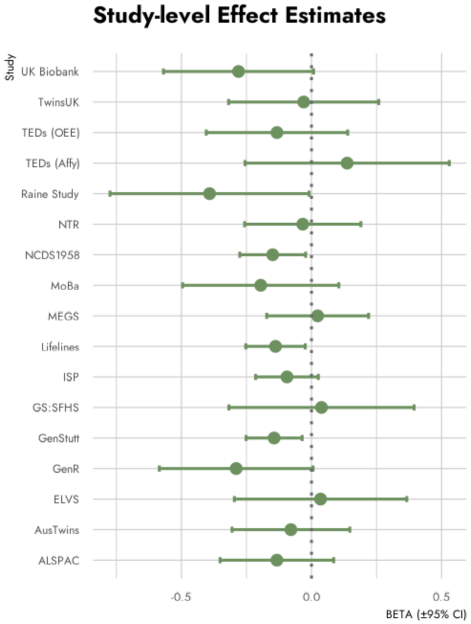

O. rs7981317 (*OLFM4*)

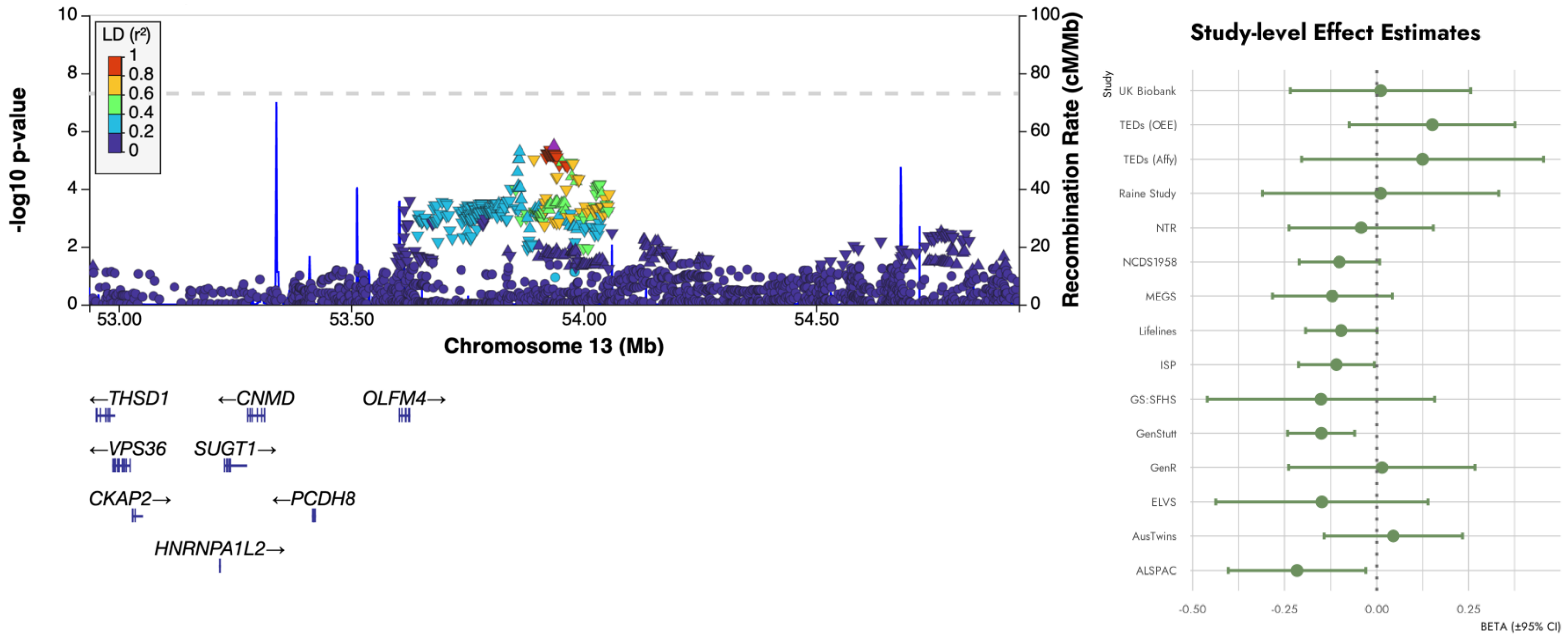

P. rs3117663 (*YY1*)

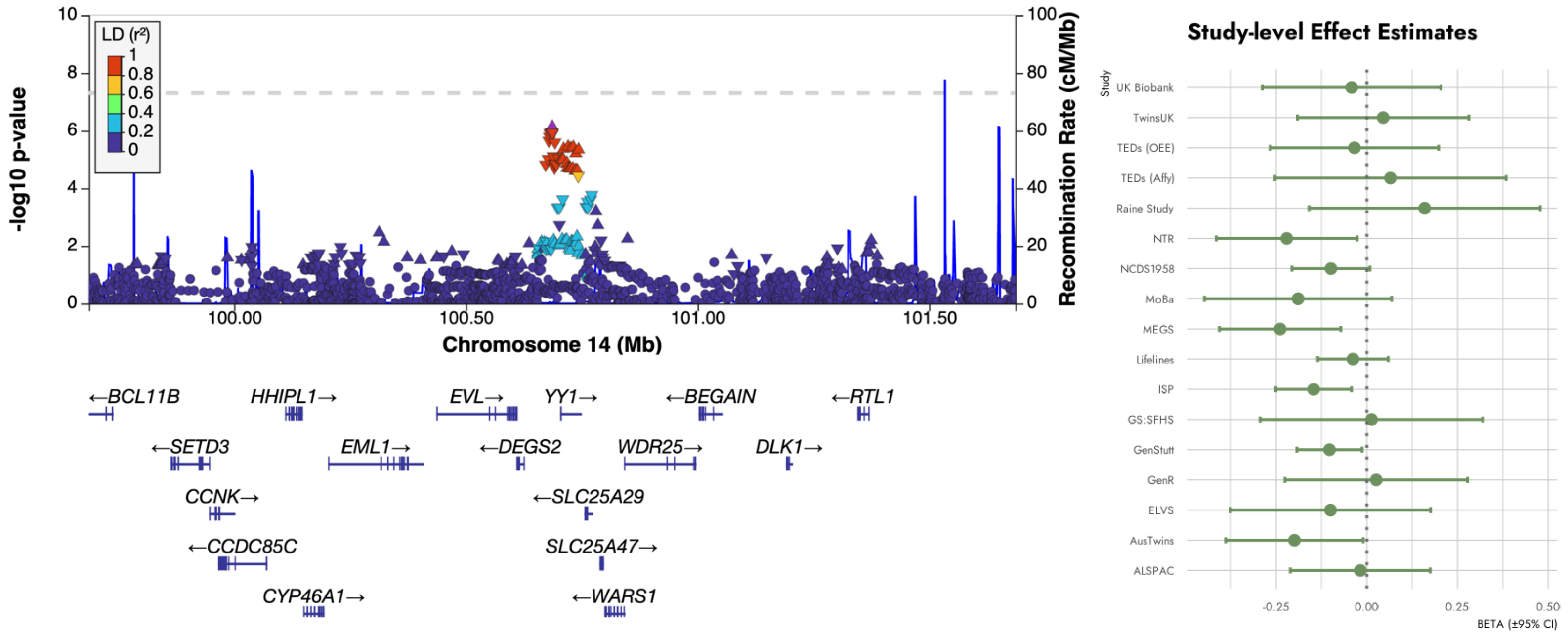

Q. rs148952809 (*DAPK2*)

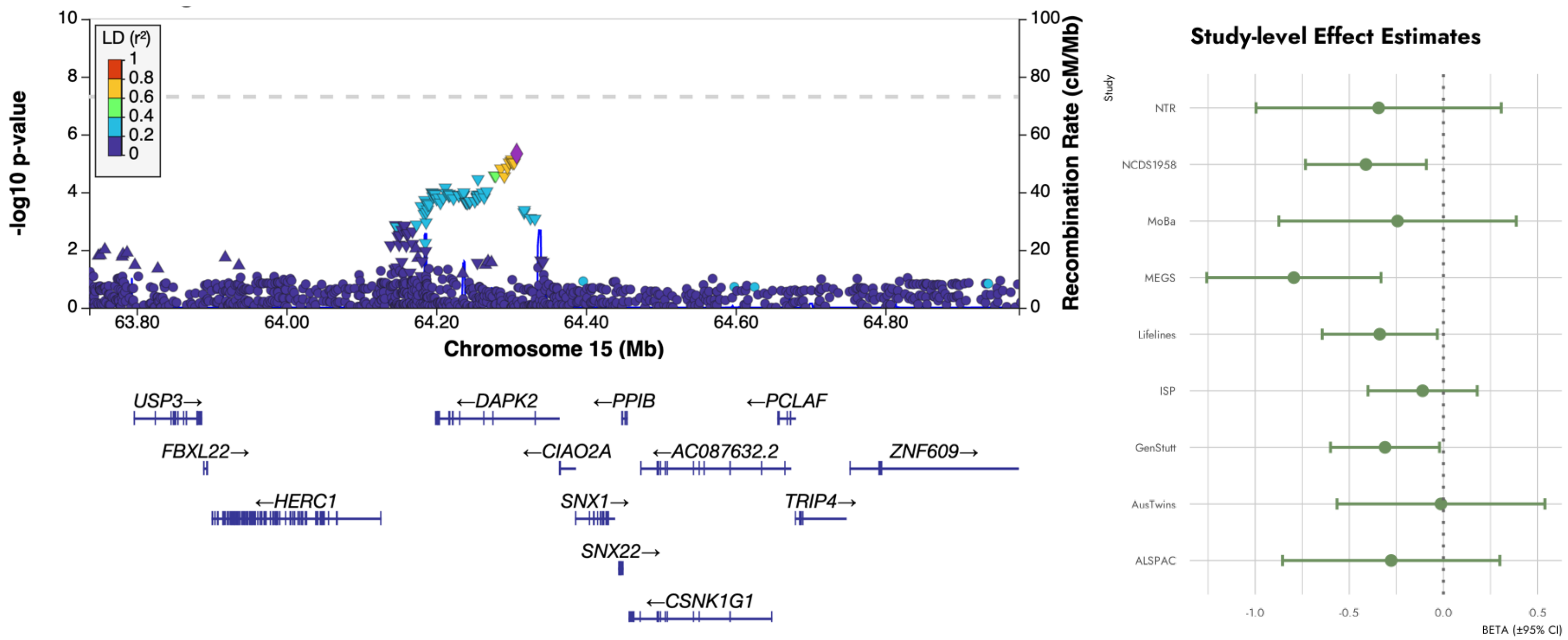

R. rs1946310 (*LINC009424*)

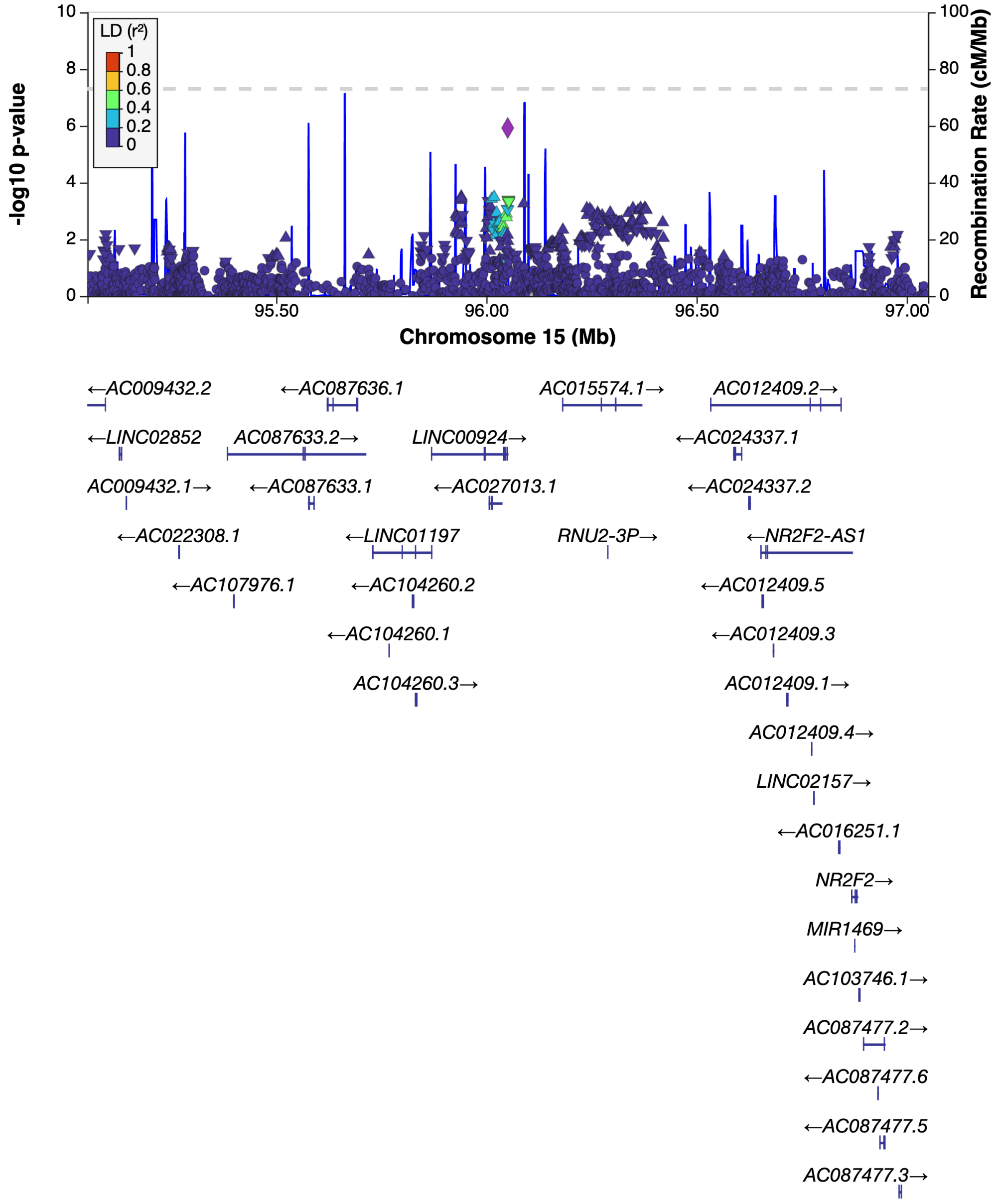


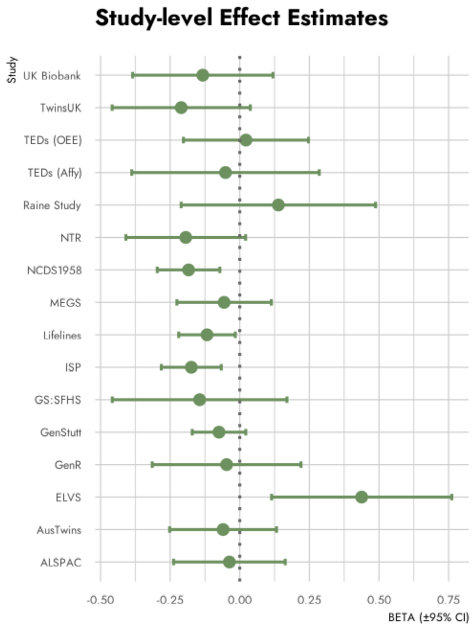

S. rs10852372 (*GRIN2A*)

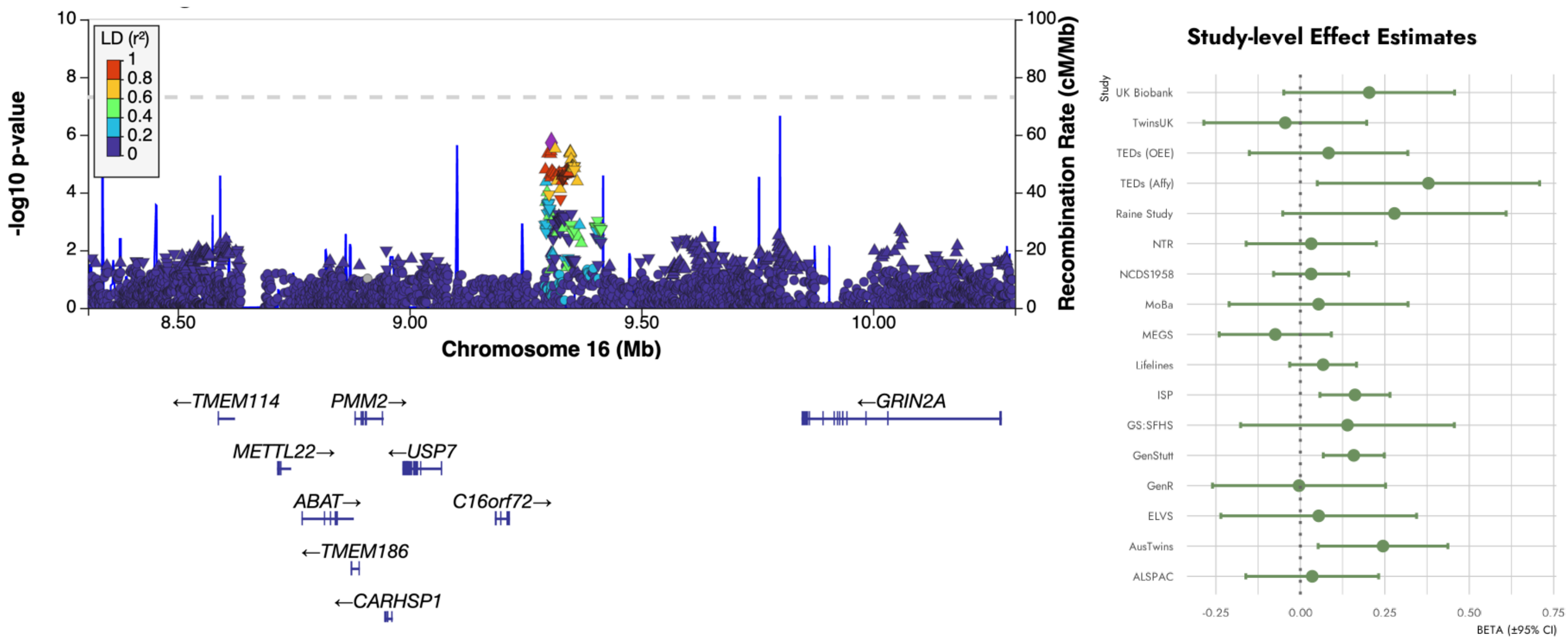

T. rs62046982 (*MON1B*)

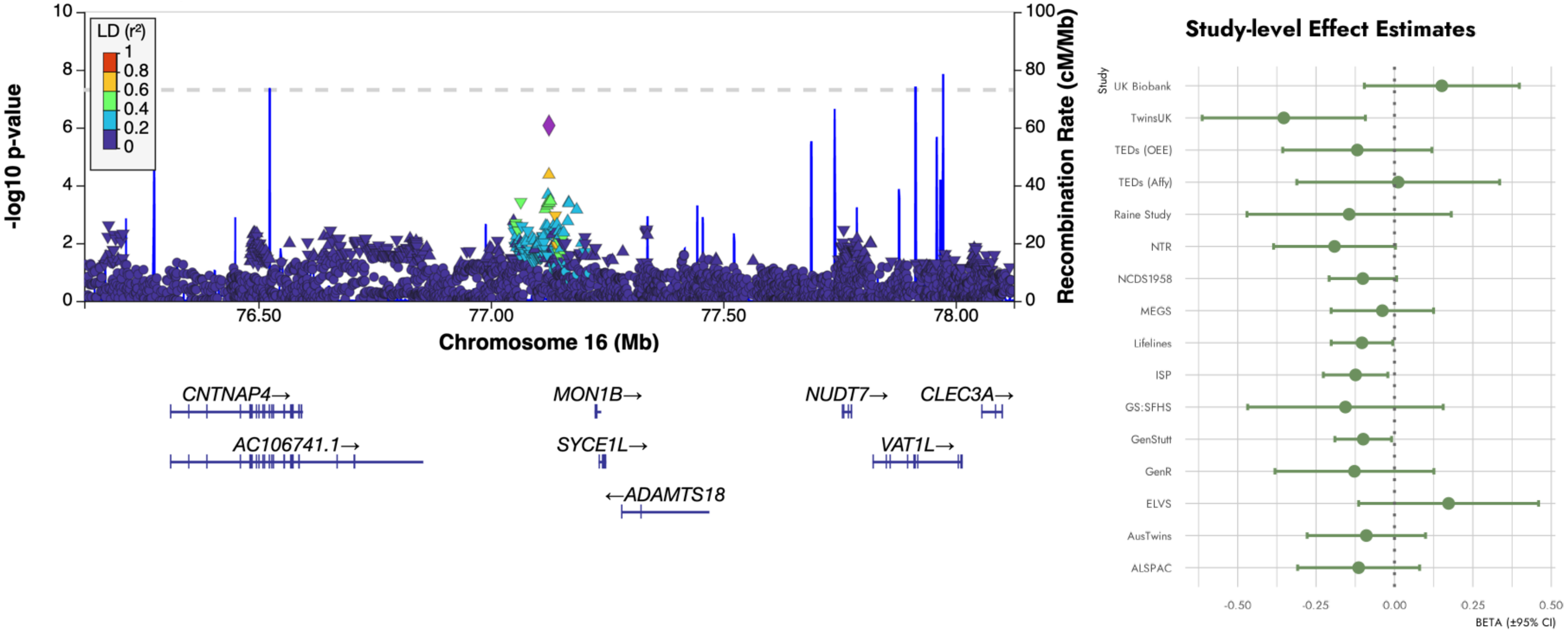

U. rs56243837 (*MAF*)

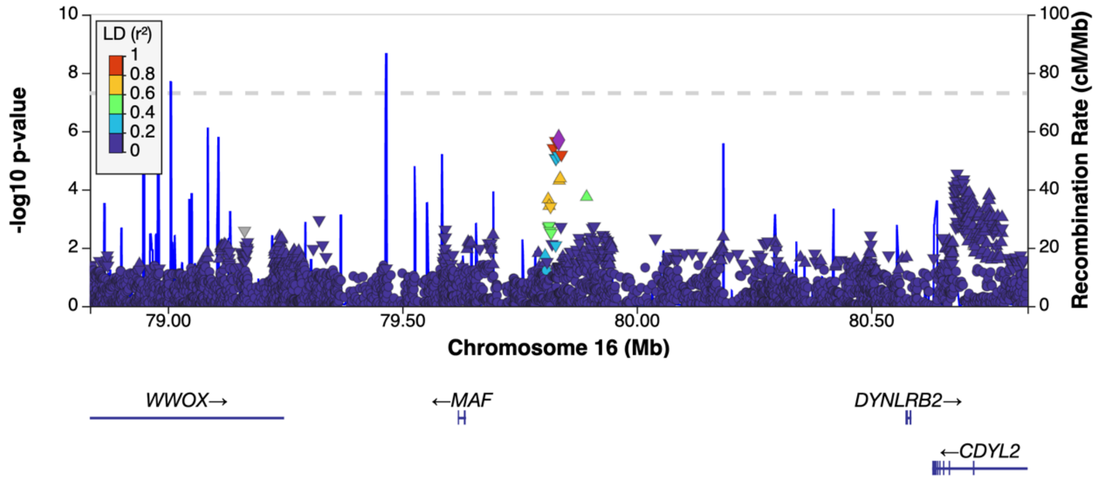


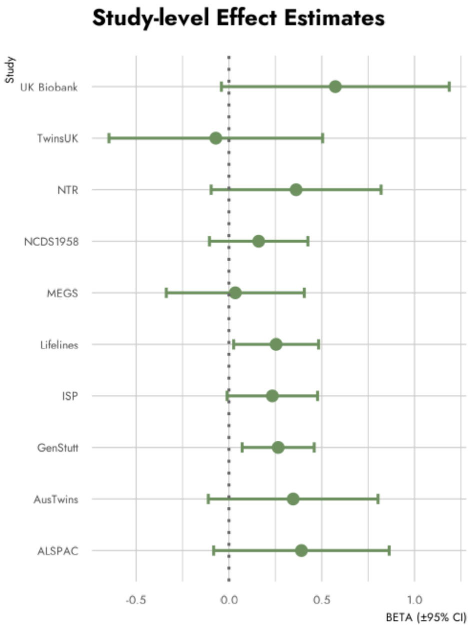

V. rs4383210 (*AKAP1*)

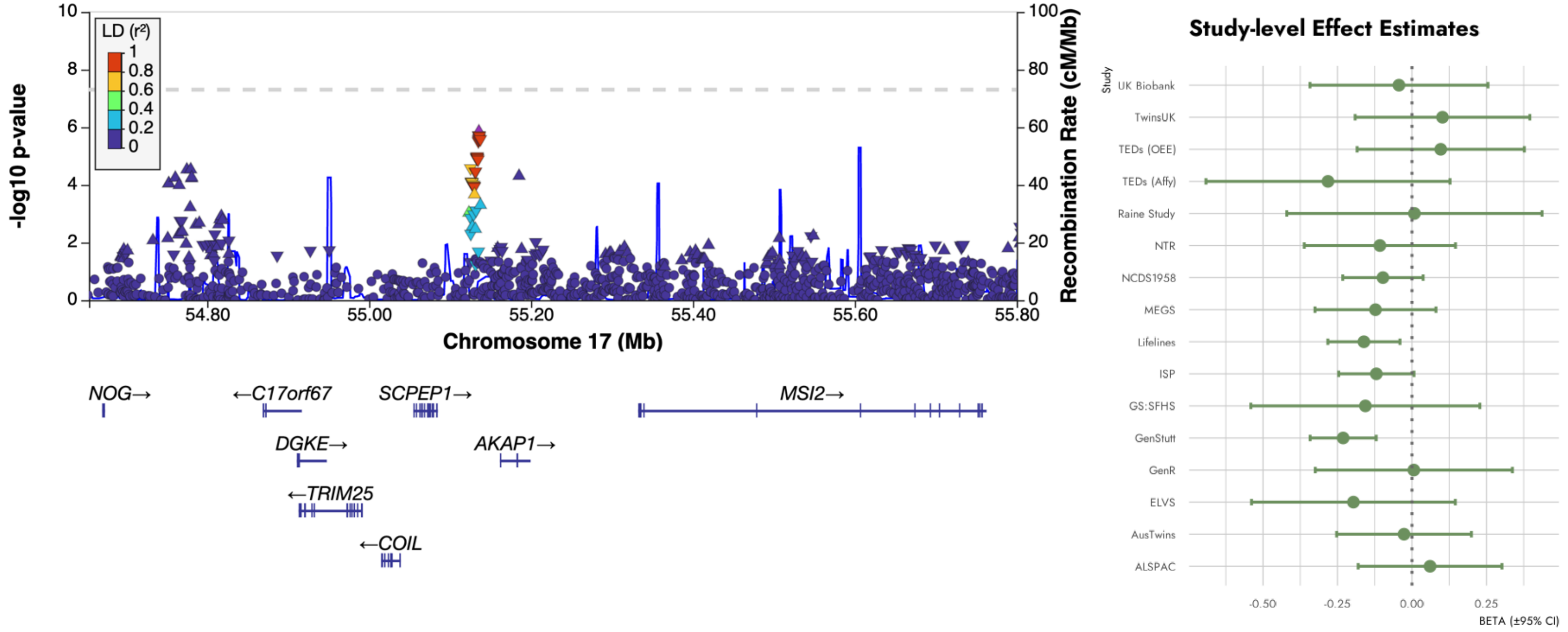

W. rs6507587 (*SETBP1*)

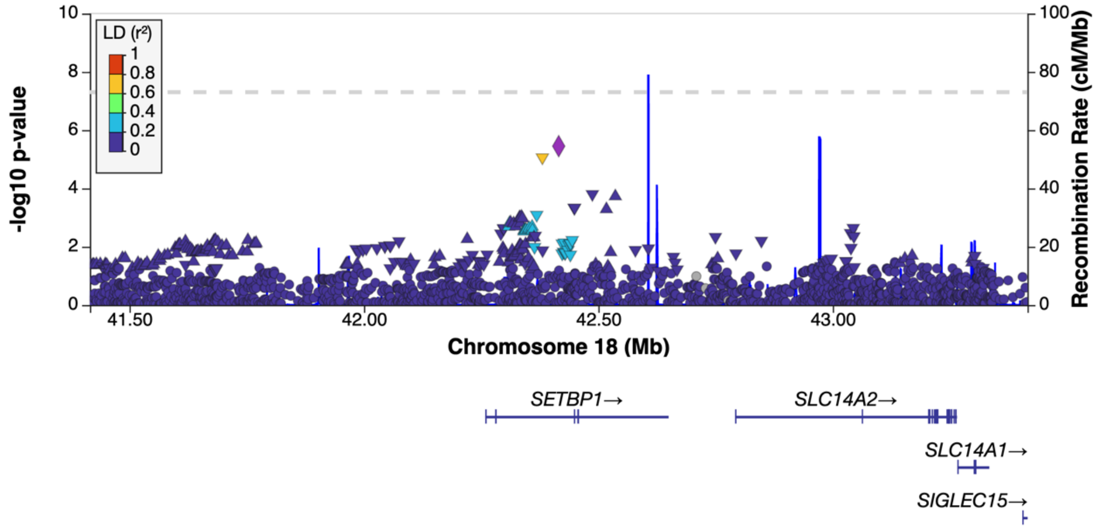


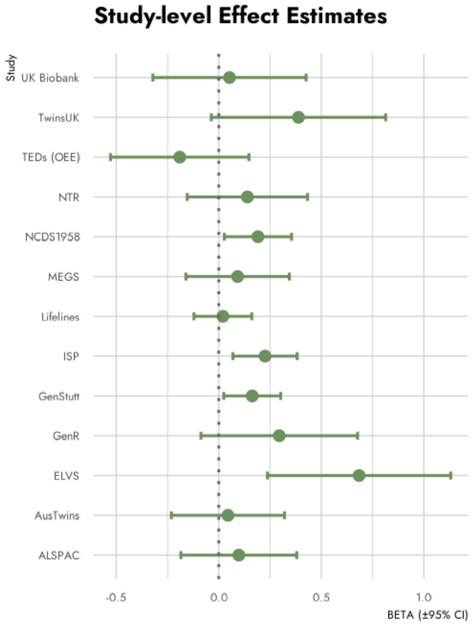

X. rs12972416 (*IRGC*)

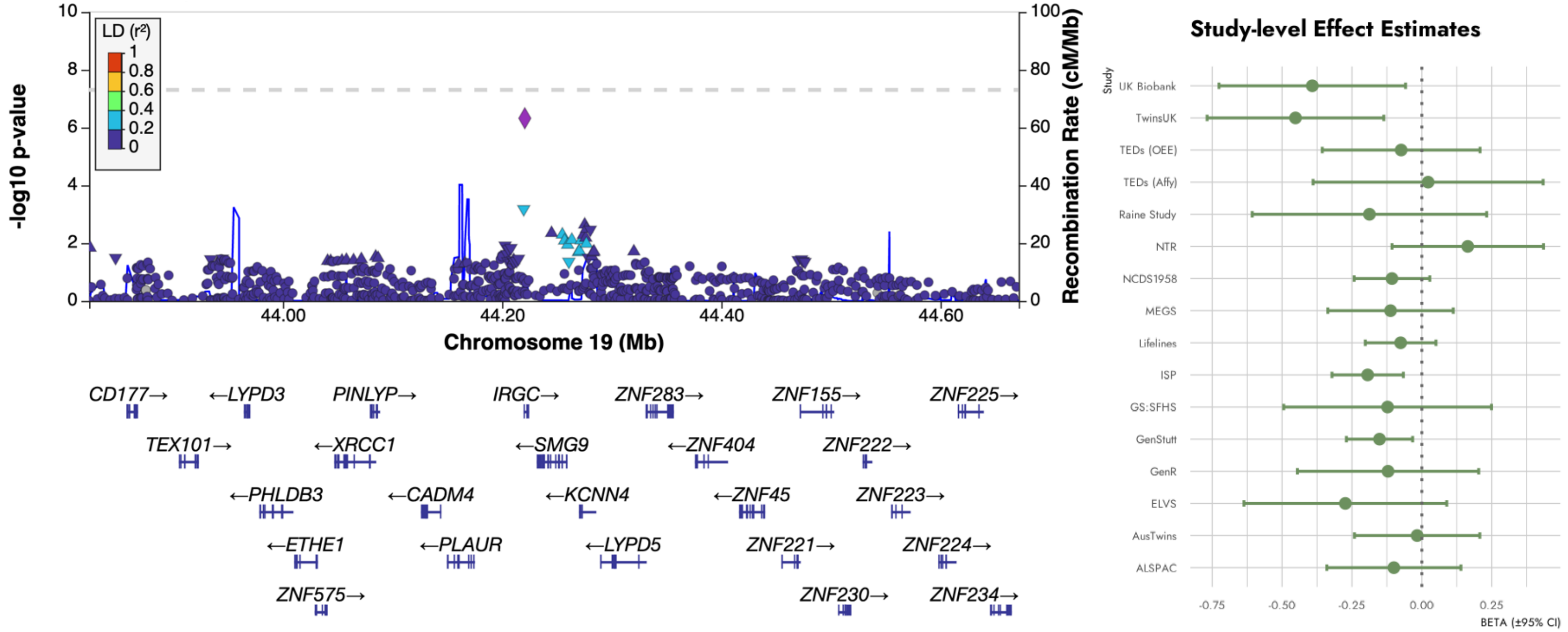

**Figure S2. Effects of 24 suggestive loci from the primary stuttering GWAS, across stuttering phenotypes.** Beta coefficients (±95% confidence intervals) are shown for each lead SNP across four phenotypes: stuttering in all individuals (stutteringAll, green), stuttering in males (stutteringMales, red), stuttering in females (stutteringFemales, yellow), and persistent stuttering in all individuals (stutteringPersistent, blue). The horizontal dashed line indicates a null effect (β = 0). SNPs are ordered by chromosomal position (x-axis).

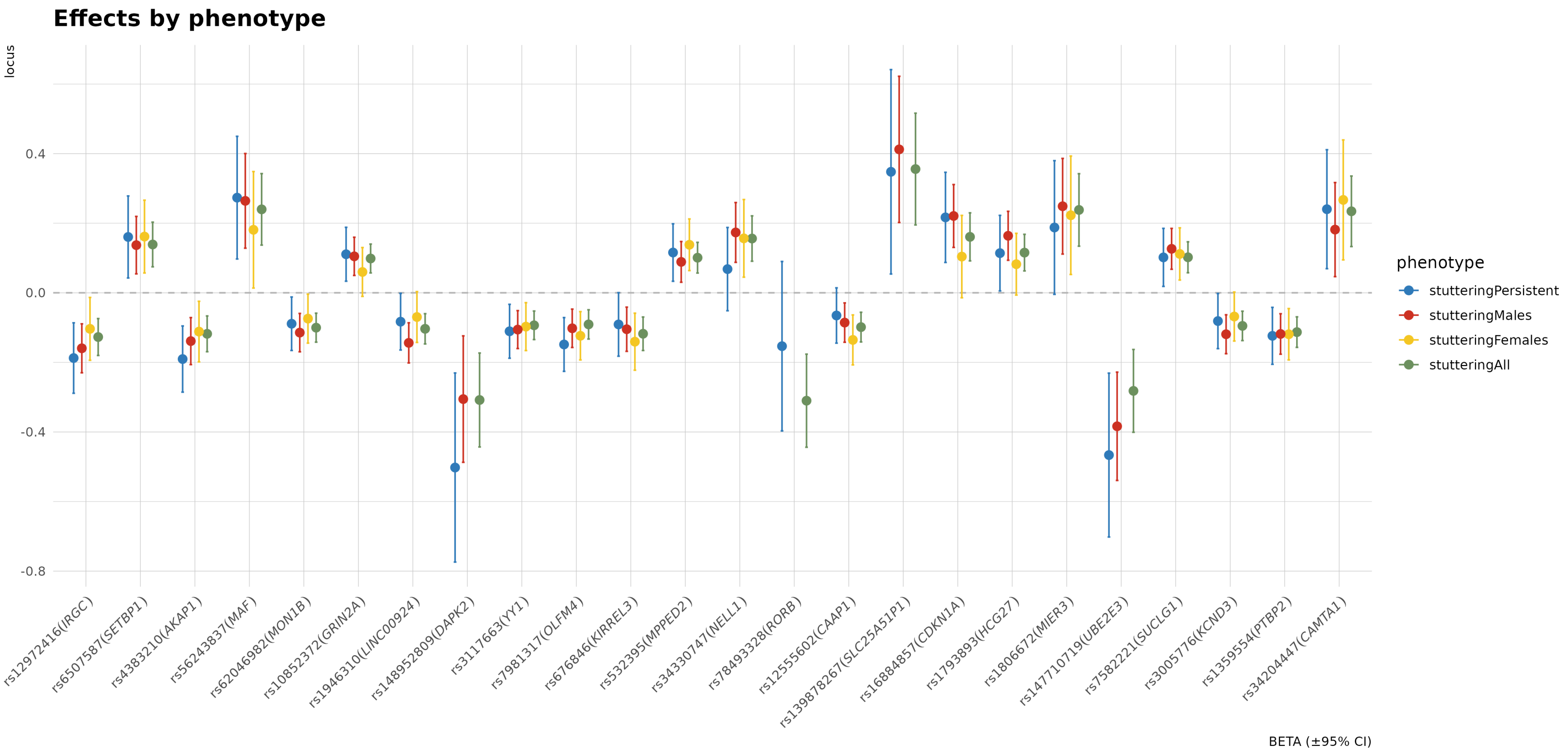

**Figure S3. A line graph of heritability estimates and corresponding population prevalences.** Heritability on the total liability scale was estimated across a range of population prevalences in increments of 0.01 using LD score regression.[33]

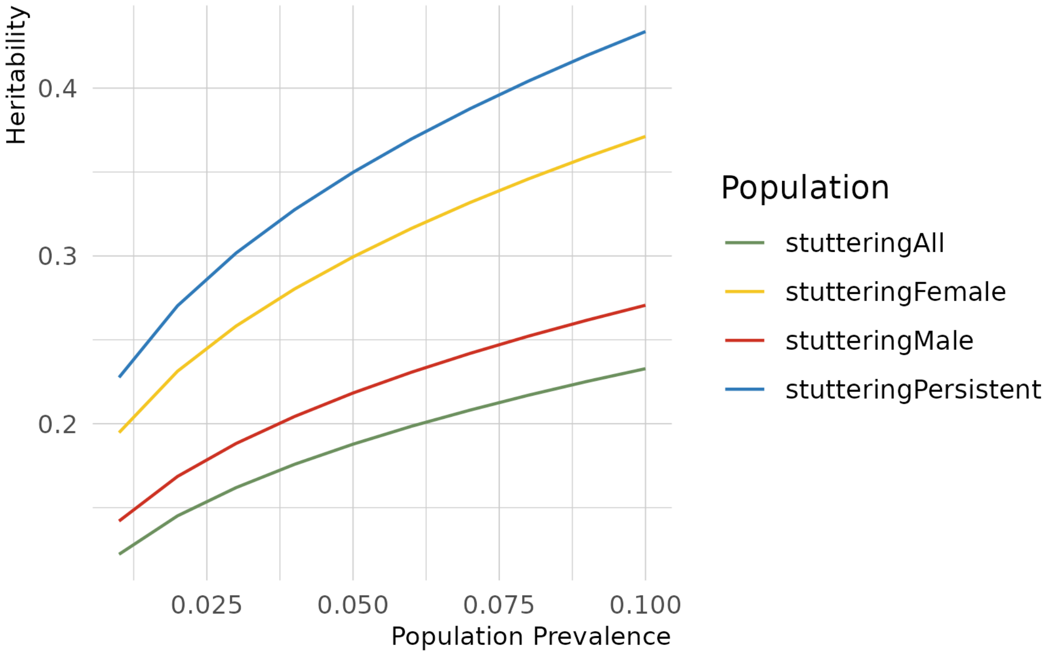

**Figure S4. Partitioned heritability enrichment of functional annotations for stuttering.** Enrichment for binary annotations from the baselineLD model (version 2.2)[34,35] are shown for **(A)** stuttering and **(B)** persistent stuttering. Error bars indicate the standard error of the enrichment. P-values from a one-sided test that the coefficient is greater than zero.

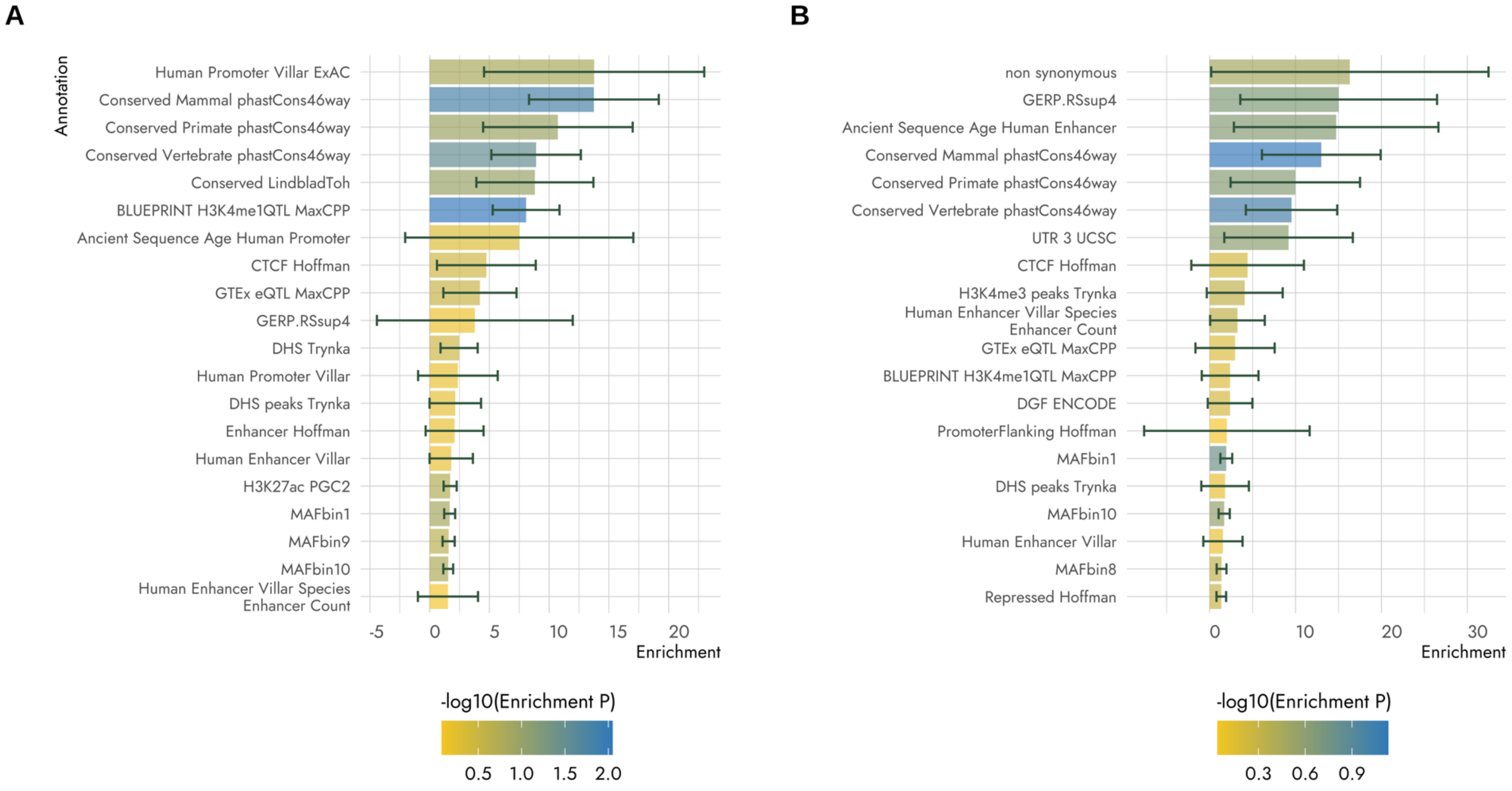

**Figure S5: Manhattan plot of genome-wide association results for stuttering in A) males and B) females.** Each point represents a single nucleotide polymorphism (SNP), plotted by chromosomal position (x-axis) and $-\log_{10}$ transformed *p*-value (y-axis). The dark dashed line indicates the genome-wide significance threshold ($p < 5 \times 10^{-8}$) and the light dashed line indicates the suggestive significance threshold ($p < 5 \times 10^{-5}$).

**A.**

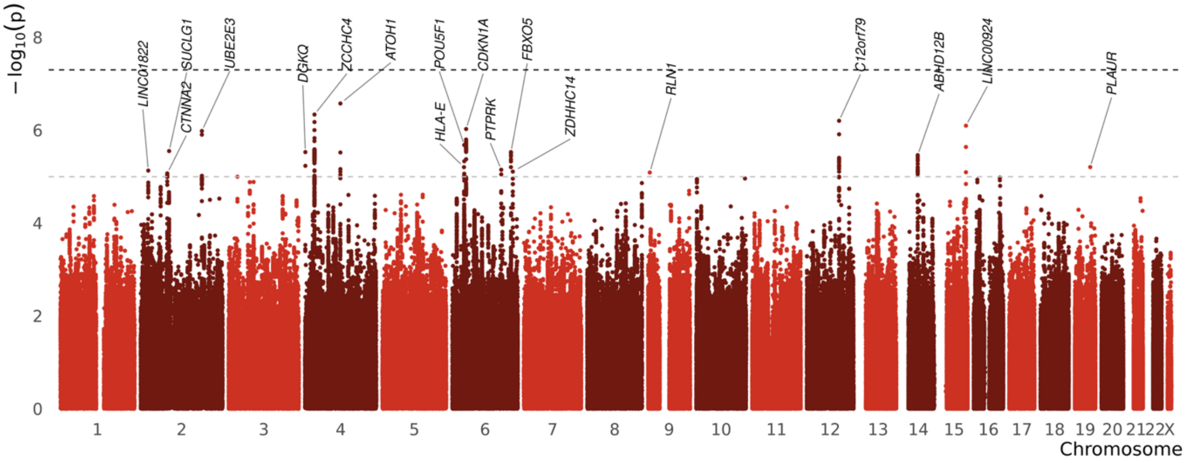


**B.**

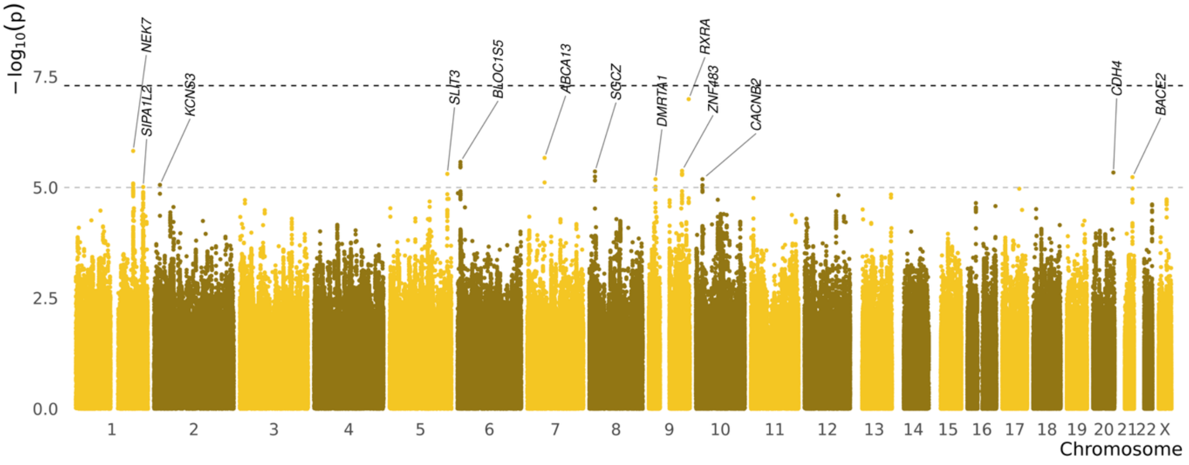

**Figure S6: Comparison of effect sizes for top SNPs in ascertained versus population-based cohorts.** Points indicate effect estimates (betas) with error bars indicating 95% confidence intervals.

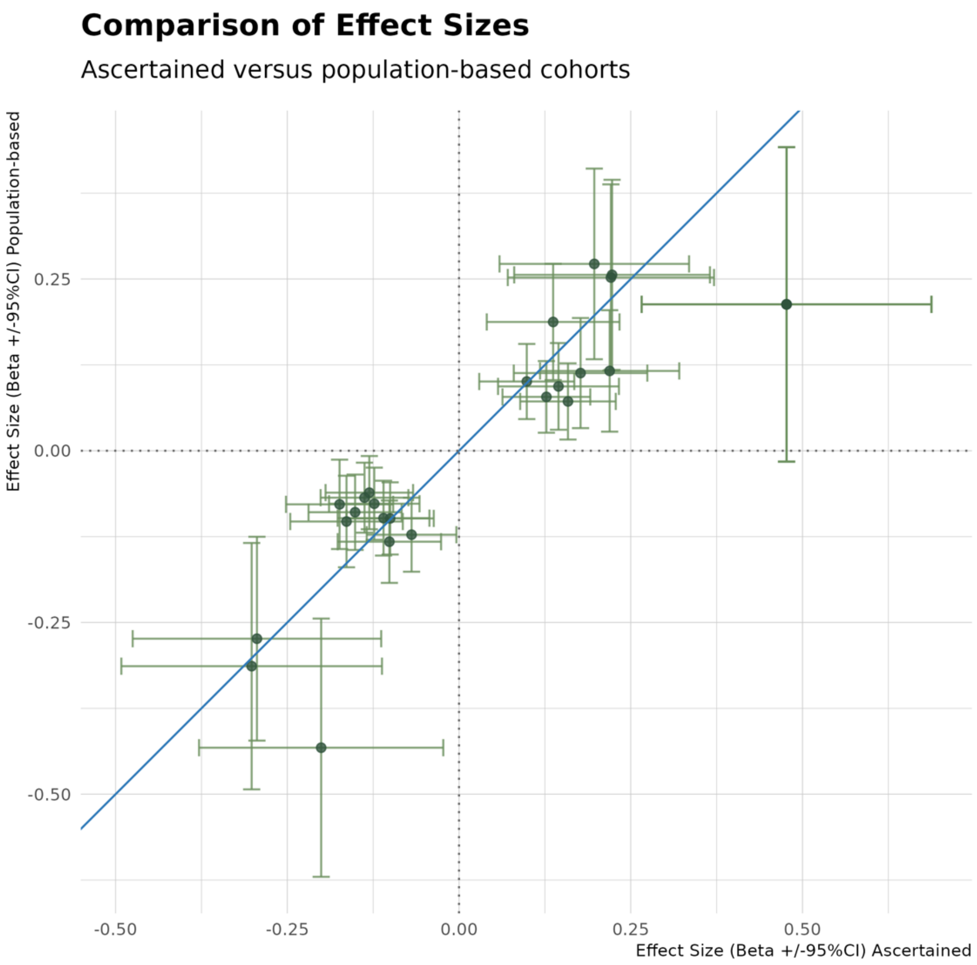

**Figure S7: Manhattan plot of genome-wide association results for persistent stuttering.** Each point represents a single nucleotide polymorphism (SNP), plotted by chromosomal position (x-axis) and $-\log_{10}$ transformed *p*-value (y-axis). The dark dashed line indicates the genome-wide significance threshold ($p < 5 \times 10^{-8}$) and the light dashed line indicates the suggestive significance threshold ($p < 5 \times 10^{-5}$).

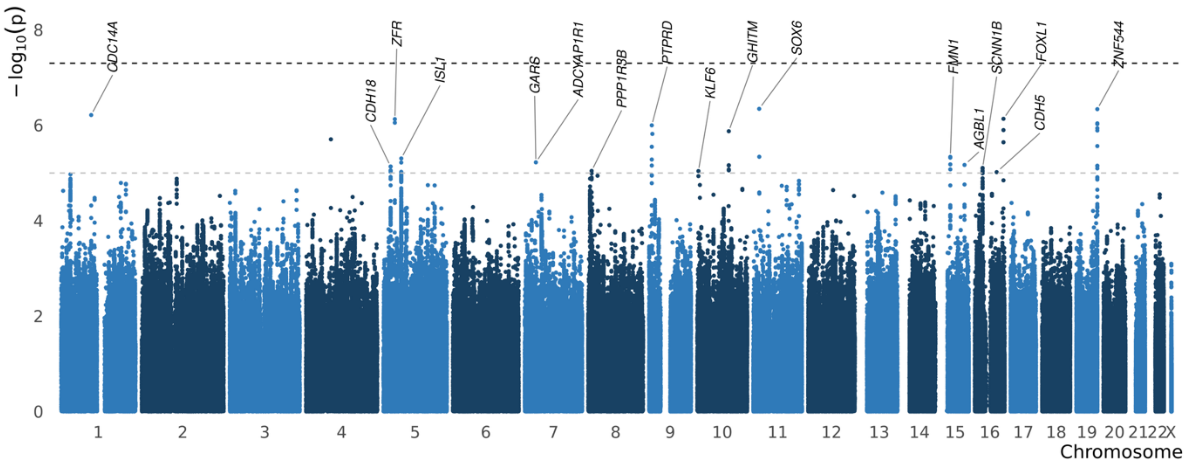

**Figure S8: GWAS-by-subtraction, genomicSEM model**

In the genome-wide model without individual SNP Effects, standardized loadings were estimated as follows: $\lambda_1$= 1.00 (SE = 0.06, $p$ = 1.79 × $10^{-59}$); $\lambda_2$ = 0.97 (SE = 0.08, $p$ = 7.21 × $10^{-36}$); $\lambda_3$ = 0.24 (SE = 0.26, $p$ = 0.35).

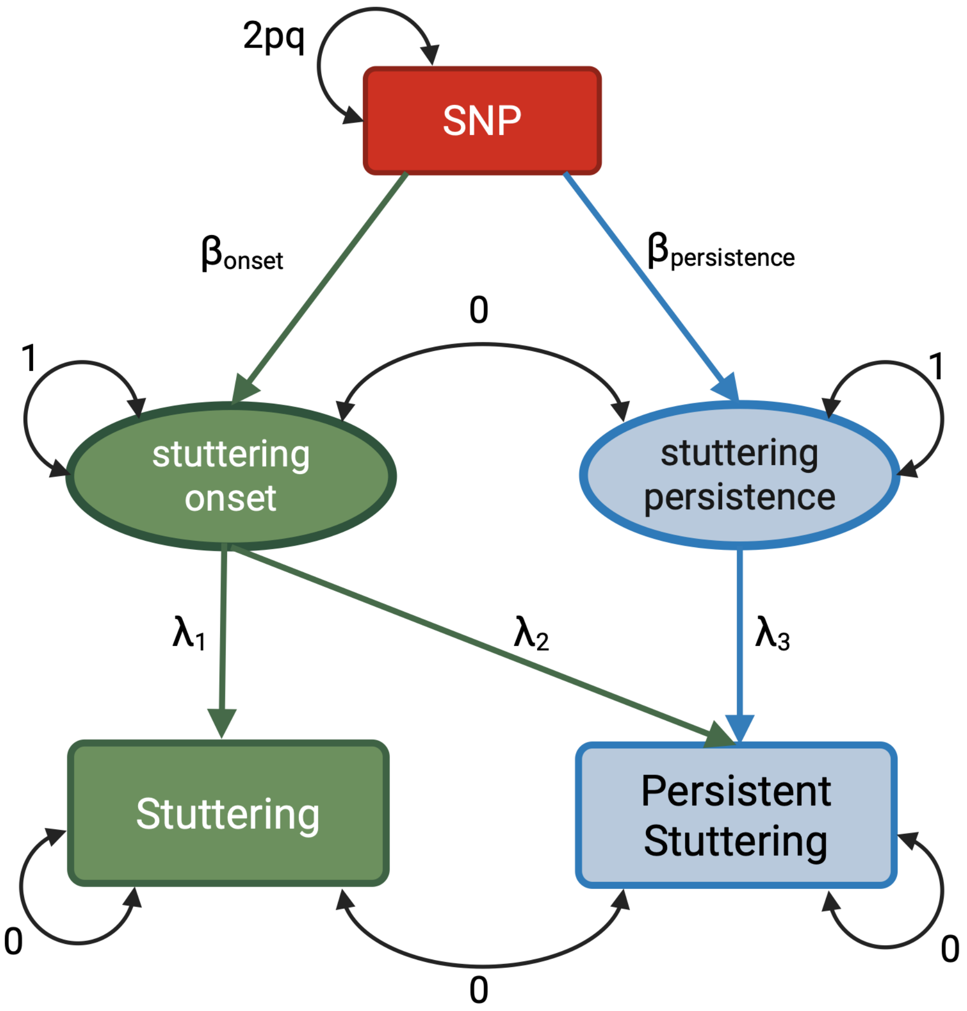

***Figure S9: Associations of 24 suggestive loci from the primary stuttering GWAS, with stuttering frequency and severity.*** *Beta coefficients (±95% confidence intervals) are shown for each lead SNP, with effects harmonised to the allele associated with increased risk of stuttering. Left panel: stuttering frequency (rated 1–5) analysed as an ordinal trait using a Proportional Odds Logistic Mixed Model. Right panel: stuttering severity (rated 1–10) analysed as a continuous trait. The vertical dashed line indicates a null effect (β = 0). * = significant at Bonferroni-corrected significance threshold of* $p < 2.08 \times 10^{-3}$ *(0.05/24 loci).*

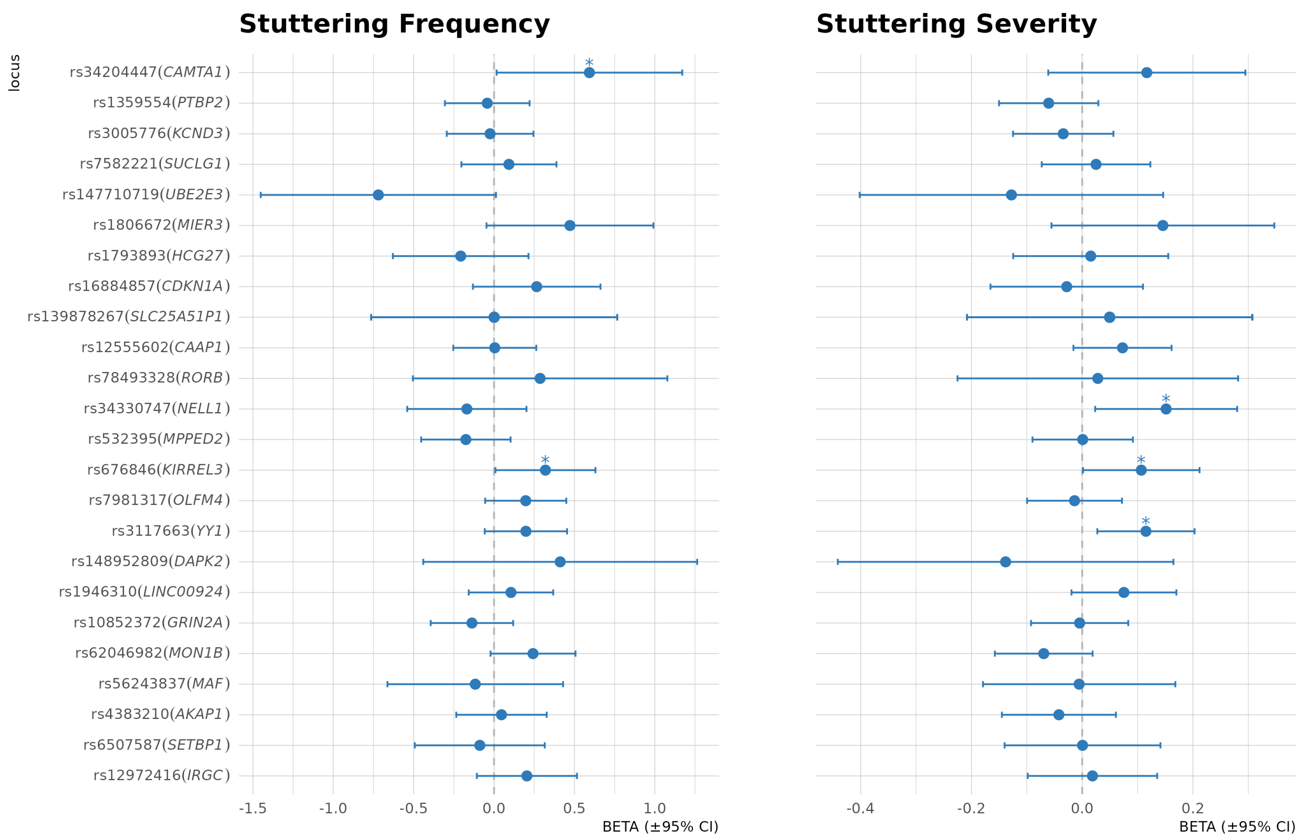

**Figure S10: Genetic correlations between stuttering and select other traits.** Genetic correlations between stuttering and selected traits across four categories: migraine, epilepsies, neurodevelopmental disorders, and language and reading. Point estimates (±95% CI) were estimated using linkage disequilibrium score regression (LDSC).

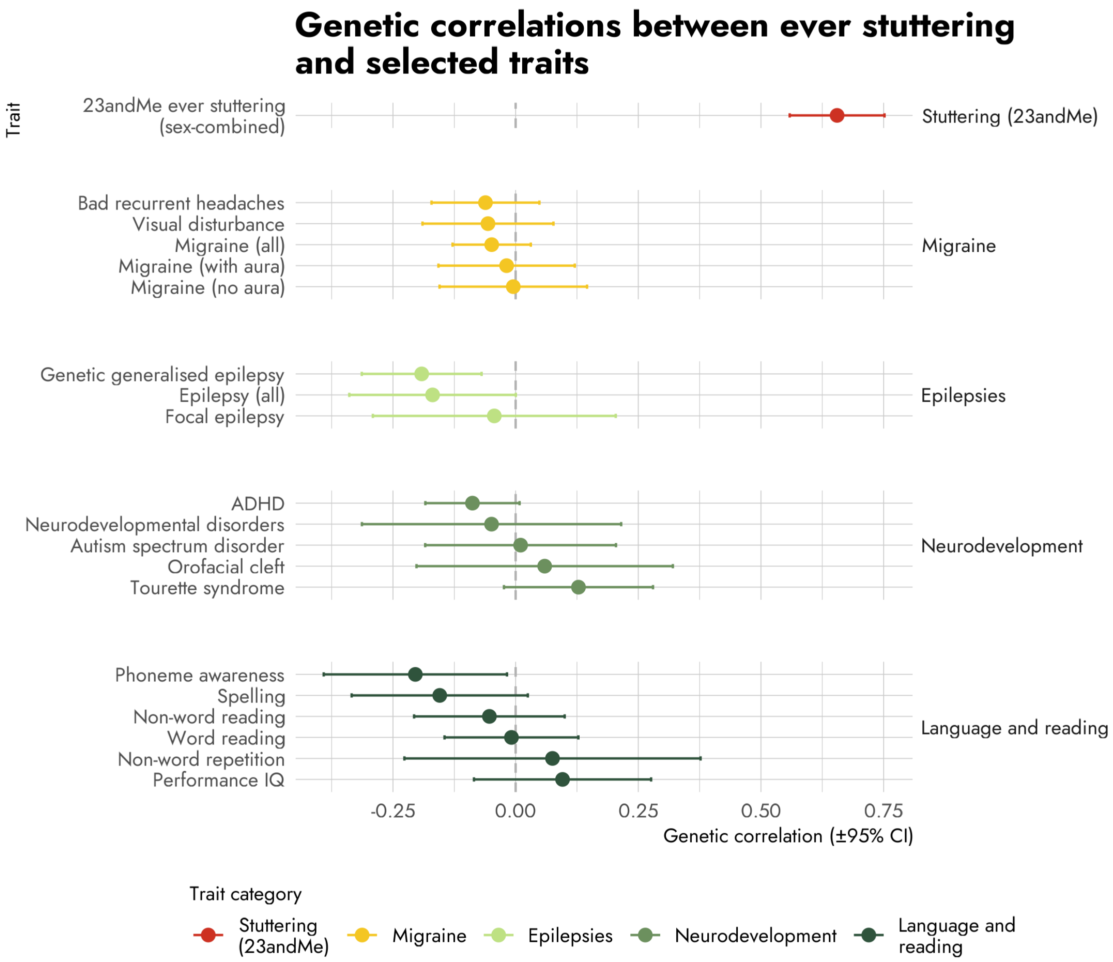

**Figure S11. Partitioned heritability analyses for stuttering using S-LDSC and gene expression datasets.**[36] Regression coefficients for each cell/ tissue type from **A)** Cahoy, **B)** GTEx and **C)** Multi-tissue gene expression gene-sets are displayed. Error bars indicate the standard error of the coefficient. P-values from a one-sided test that the coefficient is greater than zero.

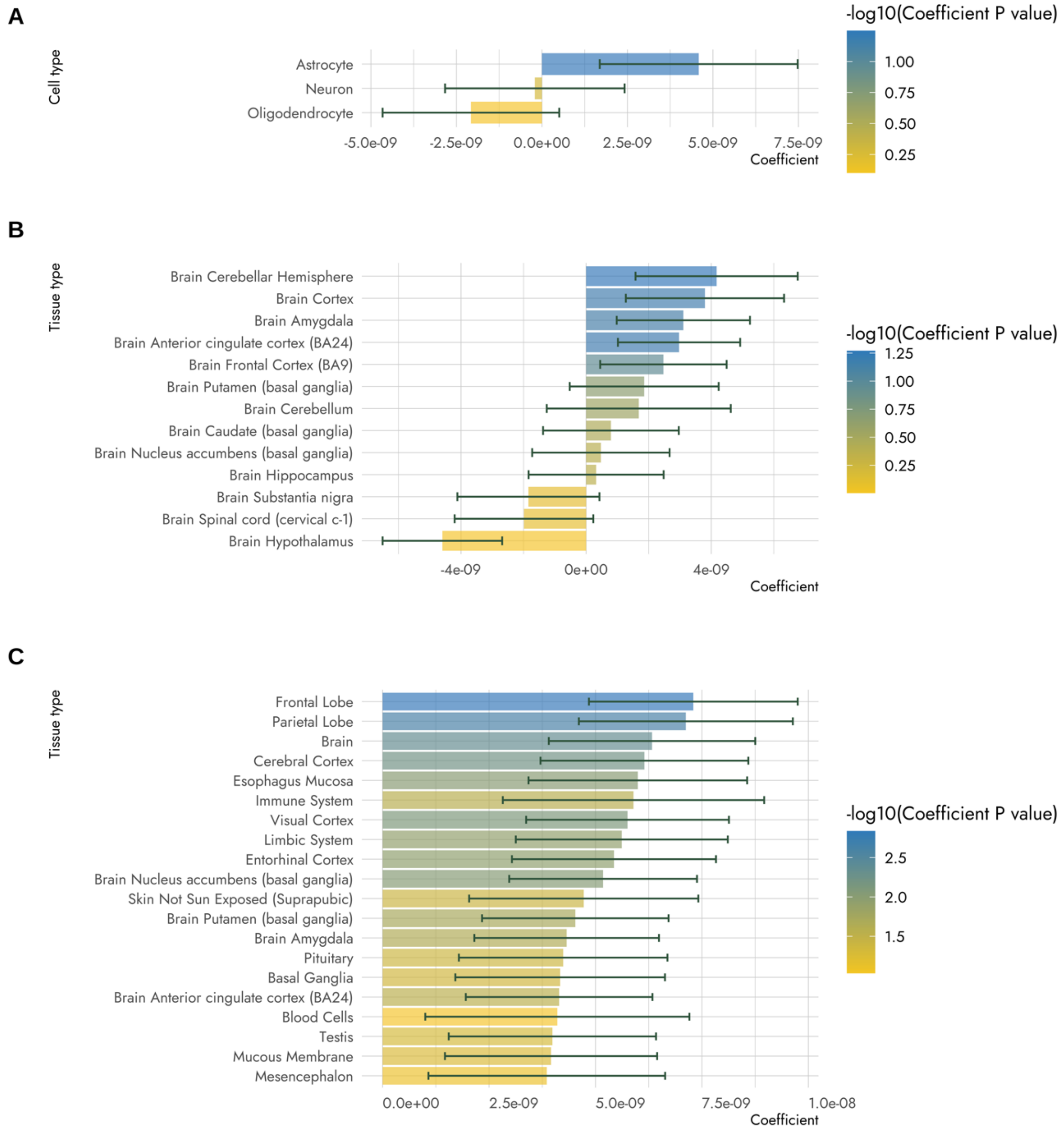

**Figure S12. Partitioned heritability analyses for stuttering using S-LDSC epigenetic data.**[36] Coefficients for tissue-specific chromatin-based annotations (Roadmap) for 10 brain regions are displayed. Error bars indicate the standard error of the coefficient. P-values from a one-sided test that the coefficient is greater than zero.

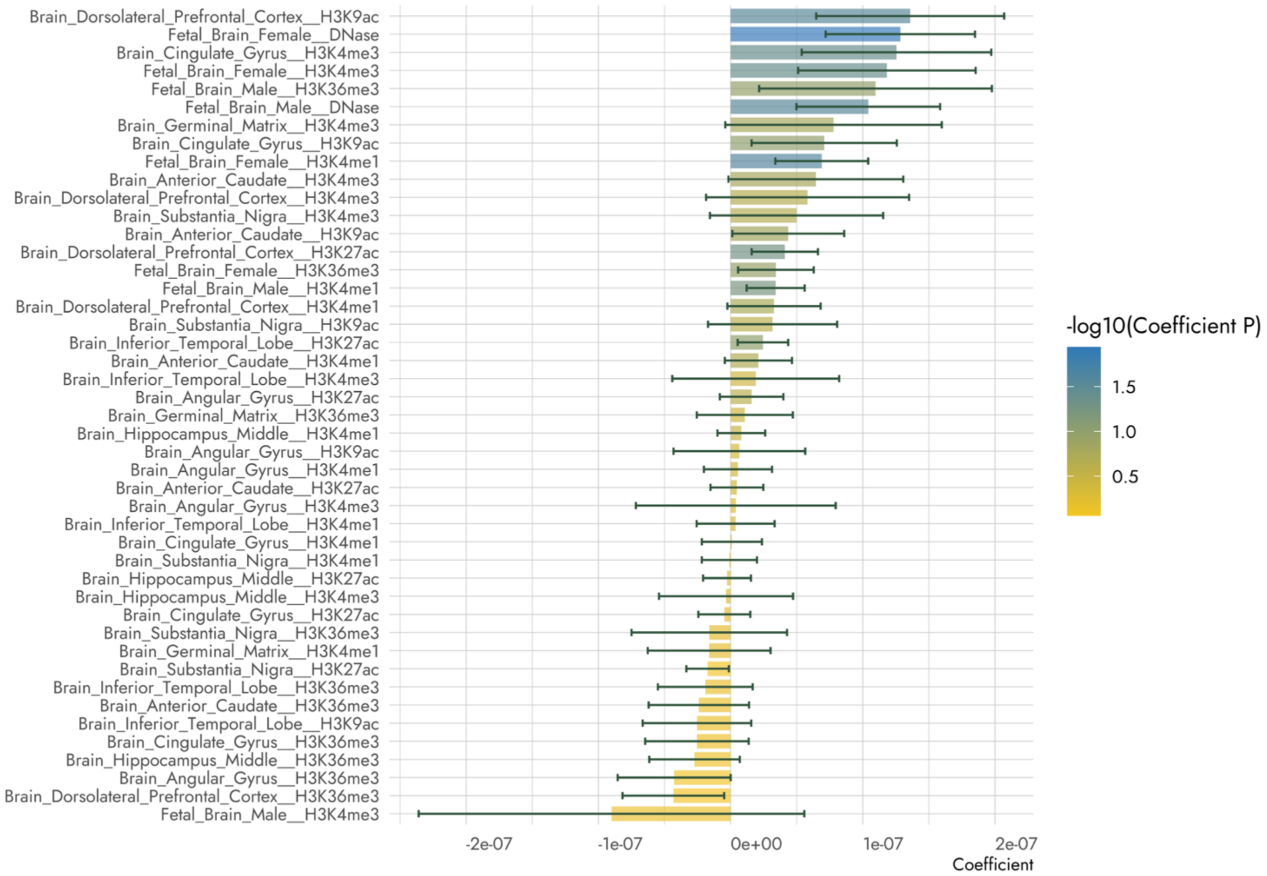

**Figure S13. S-BrainXcan association with Structural (T1) MRI features in stuttering displayed by brain region**. Structural MRI features displayed include **(A)** subcortical grey matter volume, (B) subcortical total volume, **(C)** cortical grey matter volume and **(D)** cerebellum grey matter volume. Brain regions with z-score > 2 or < -2 are labelled.

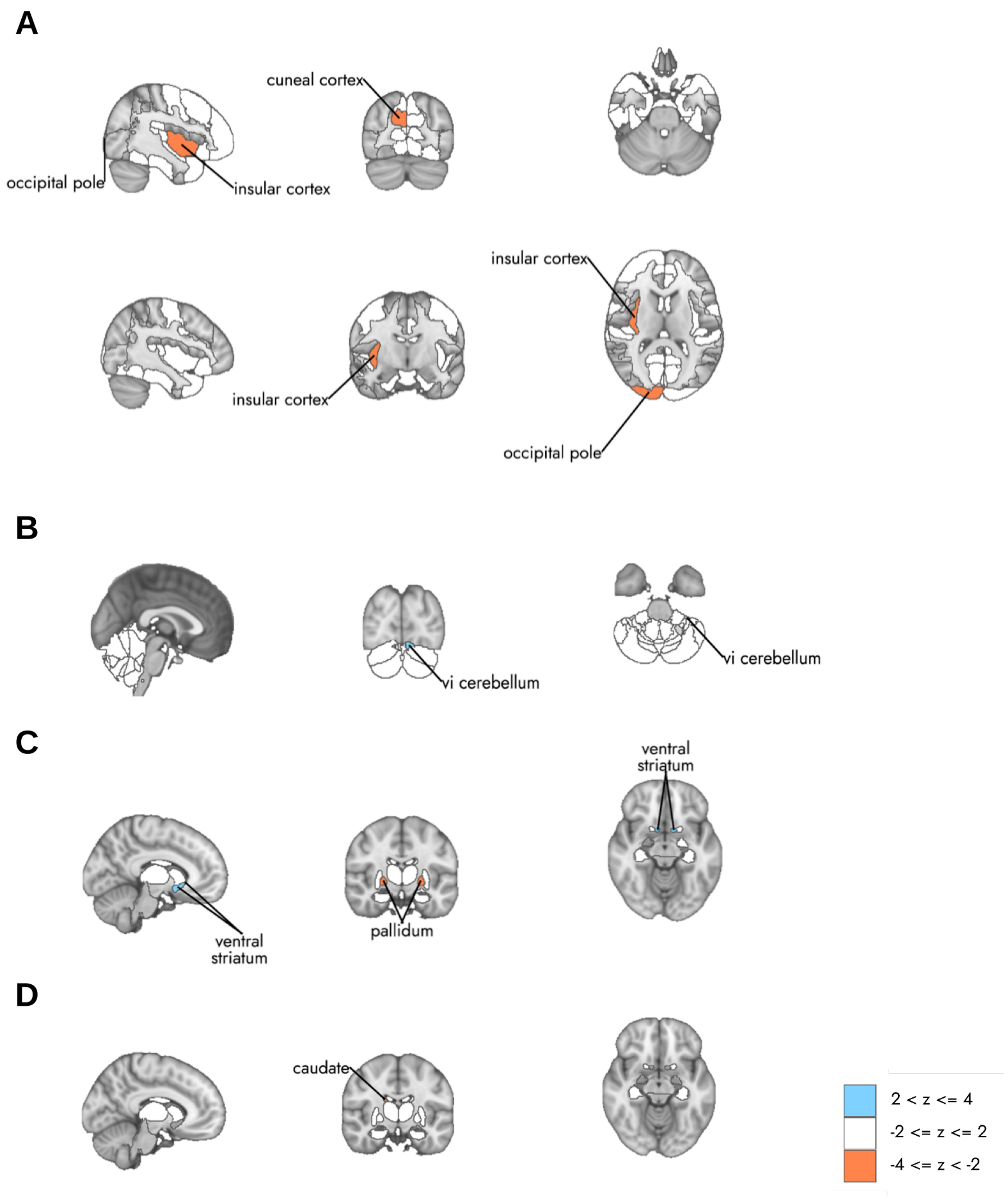

**Figure S14. S-BrainXcan association with diffusion MRI features in stuttering displayed by brain region**. Diffusion MRI features displayed are (A) fractional anisotropy, (B) intracellular volume fraction, (C) orientation dispersion index and (D) isotropic or free water volume fraction, generated from tract based spatial statistics (TBSS). Brain regions with z-score > 2 or < -2 are labelled.

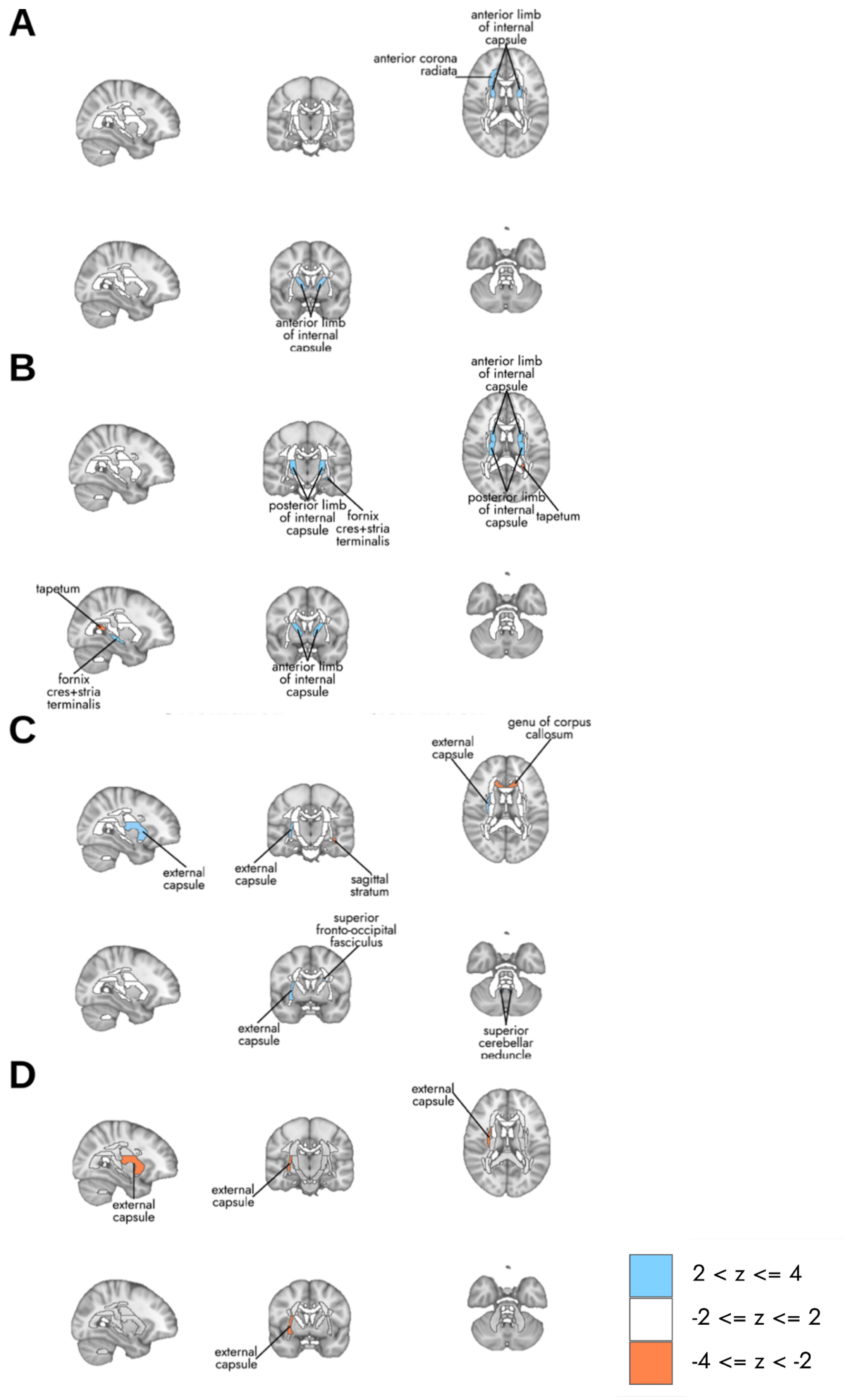
